# A Two-Region SEIR COVID-19 Epidemic Model for the Island of Ireland

**DOI:** 10.1101/2020.10.31.20223727

**Authors:** James J. Grannell, James R. Grannell

## Abstract

The island of Ireland consists of two countries, Ireland and Northern Ireland, which are separated by a land border. We develop a model for the COVID-19 epidemic which consists of two SEIR models, one for each country, coupled through border interaction terms. The model incorporates symptomatic and presymptomatic infectives, but not asymptomatic infectives, together with a simple isolation/quarantine model. The objective of the work is to explore how the two-region epidemic could evolve by examining selected regions of parameter space. In this context we examine the effect of the border status on evolution of the epidemic. We found that, even though the border interaction parameters are relatively small, the open border could significantly affect the course of the epidemic in some of the scenarios studied. We also looked for and found examples of sensitive dependence on several parameters.

## Introduction

Could different government policies and societal behavior in Northern Ireland affect the course of the COVID-19 epidemic in Ireland and vice versa? The two governments have been tackling the epidemic on two fronts: contact/transmission reduction and reduction of duration of infectiousness. Transmission reduction measures include social distancing, mask wearing, avoidance of face touching and enhanced hygiene. Contact reduction measures include closure of schools and businesses, working from home, reducing social contacts and cocooning. Infectiousness duration reduction measures include encouragement of early reportiong of symptoms to general practitioners coupled with self-quarantining or self-isolation and tracing of close contacts of known infectives followed by quarantining or isolation. The Northern Ireland government broadly followed British government policy which has been different to Irish government policy. Northern Ireland policy is also constrained by funding from Westminster. The British government stopped testing/tracing on 12th March, but reinstated it several months later. The Irish government continued to carry out testing/tracing since the beginning of the epidemic. However, the number of tests per capita in the UK climbed above that in Ireland on 18th May and remained above it ever since [2]. The Irish government introduced contact/transmission reduction by closing schools and third-level colleges on 12th March - a week before Northern Ireland. Both governments subsequently closed businesses and restricted travel and instituted cocooning/shielding for the over-70s. Easing of restrictions on a phased basis was subsequently implemented in Ireland and Northern Ireland. Several phases of increased restrictions were again introduced in both Ireland and Northern Ireland, between August and October 2020. If the timing and nature of measures and societal responses are different in the two countries, could progress in one region be impacted by measures taken in the other region in the presence of an open border?

We develop an SEIR model for each region and connect them by introducing border interaction terms. The resulting two-region model is crude in that it only incorporates one class of infectives in each region and has limited capacity to model quarantine/isolation efforts. We did not seek to model the history of mitigation efforts in both countries up to a chosen time and make predictions moving forward from there. Rather, our objective was to explore possibilities for how the all-island epidemic could evolve in each of the countries and to examine how the border status, together with different policies/societal responses in the two countries, could affect the evolution in each country. A second objective was to see if the model could exhibit sensitive dependence on any parameters, in the sense that a small change in a parameter could cause a large change in the evolution of the epidemic. Such findings would be of value in exploring more advanced models incorporating asymptomatic infectives and more realistic testing and contact tracing. Behaviour found in a simple model may persist in a more advanced model. Such explorations are also valuable for policy makers in making them aware of what might be possible in a new situation.

We start by simply modelling the evolution of the epidemic in the presence of a single contact/transmission reduction event in each region. We then model a single contact/transmission event followed a single restriction alleviation event. Several restriction alleviation scenarios are considered: e.g. a single alleviation event in one region prior to alleviation in the other or a simultaneous alleviation event in both countries. We incorporate self isolation/quarantining, but not government testing/tracing/isolation/quarantining. Even in the context of our simple model, many scenarios could be considered. We decided to focus on some very simple scenarios in order to investigate the effect of border coupling. We look at what the model predicts if

1. the border had been closed on the first day of the epidemic
2. the border was left open and different policies were pursued

We find that the border status has little effect in some scenarios, but that it can can have large effects in other scenarios. Especially important is the finding that the two-region model can predict that different policy/societal response in one country can have a profound effect in the other country when the border is open, even though a single country model for the affected country implies the epidemic is well controlled. Given that closing the border may not be possible politically, we consider model implications for a coordinated all-island policy. We also look for sensitivity of the model to small changes in parameters and find that such phenomena can occur.

We first present the one-region SEIR model equations which would apply if the border were closed. We then present the two-region coupled SEIR model, incorporating interactions at the border. We discuss the choice of parameters in the model and present the results of simulations for various scenarios, together with conclusions and recommendations.

## The-One-Region SEIR Model

### Model equations

The one-region SEIR model, in the context of this study, can be applied to either Ireland or Northern Ireland as a single region which does not interact with the other. The SEIR one-region model equations, ignoring births, non-COVID-19 deaths and migration, are [3], [4], [5]:

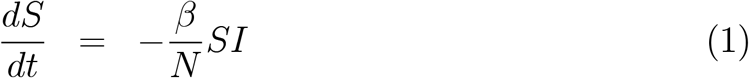

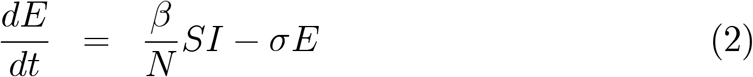

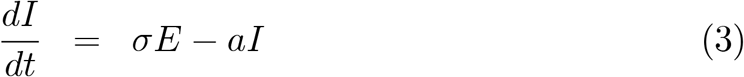

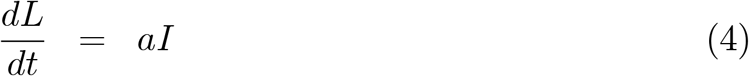

for *t* > 0, with initial conditions:

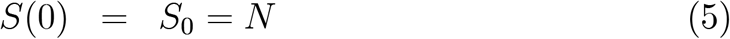

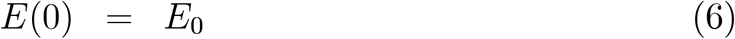

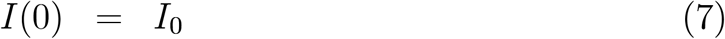

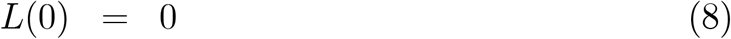

where *S, E, I* and *L* are the susceptible, exposed, infective and removed sub-populations, respectively, and *N* is the total population. The contact/transmission parameter is *β*. The specific loss rates of exposed and infected people are *σ* and *a*, respectively (by specific rate we mean rate per head). It is assumed that all people are initially susceptible to the COVID-19 virus and that there are no recovered people initially. We use *L* rather than *R* for the removed to avoid confusion with the effective reproductive ratio *R*(t), which evolves with time. The basic reproduction ratio is denoted by *R*_0_ and equals *R*(0). The letter *L* indicates those who have left the infective group.

The incubation period is that period following contraction of the disease before symptoms appear. It has been discovered that people in the later part of the incubation period can infect others before they develop symptoms [1], [6], [7]. We define the exposed group to consist of people who have been infected, are not showing symptoms and are non-infectious (also called the latent group). In this model, we group two kinds of infectives together: presymptomatic infectives and symptomatic infectives. The former are infecting before the end of the incubation period and the latter start infecting at the end of the incubation period. There is another (possibly large) group of people who are always asymptomatic, but less infective than the presymptomatic and symptomatic infectives [8], [9], [10], [11]. This group is not included in the model.

Both governments implemented transmission/contact reduction policies at different times [12], [13]. The effects of such interventions are incorporated in the model by allowing *β* to vary with time. In addition, the Irish government carried out testing, identification, tracing and isolation of infectives from the beginning of the epidemic in Ireland. Testing was carried out initially in the Northern Ireland, but suspended on March 12th and then reinstated several months later. In addition to government contact/transmission reduction and tracing/isolation/quarantine interventions, people in both countries, as they gradually realised the seriousness of this new disease, probably self-isolated/quarantined following development of symptoms, in conjunction with seeking and acting on advice from their GPs, as is thought to have happened in Wuhan [41], [35]. Self-isolation/quarantining is included in the model by simply choosing a value of the parameter a. This effectively assumes that the self-isolation/quarantining rate of infectives is proportional to the number of infectives. This is probably the case in the context of self-isolation/quarantining of people in conjunction with GP advice. Testing and isolation following testing was variable in both countries, but was much smaller in Northern Ireland than in Ireland. However, after resumption of testing in the UK, the number of tests per capita in the UK as a whole has consistently exceeded that in Ireland since 18th May 2020. If the number of infectives removed per day throgh testing/tracing were proportional to the number of infectives on that day, removal of infectives through testing/tracing could be incorporated into the model by simply modifying the value of the parameter *a*. The number of people tested each day and the fraction of those testing positive and being isolated is known [38], [12]. Similarly, the number of contacts of those tested who were quarantined/isolated is also known, but not publicly available. However, the number of infectives (presymptomatic/symptomatic) each day is not known. So, at best, an assumption might be made, e.g., that the known number of presymptomatic/symptomatic infectives is proportional to the observed number of presymptomatic/symptomatic infectives. If the removal rate of infectives through testing/tracting was not simply proportional to the number of infectives, incorporating the removal of infectives would involve additional parameters and differential equations in the model. We decided to only incorporate testing/tracing for which the removal rate is proportional to the number of infectives to keep the model as simple as possible in this first exploration of possible model epidemic behaviour.

We assume that the parameter *σ* is a biological parameter that cannot be influenced by government policy or social behaviour and that it is constant throughout the duration of the epidemic.

The equations were scaled and non-dimensionalised as follows:

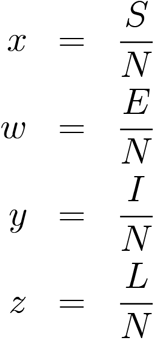

and

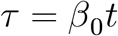

where

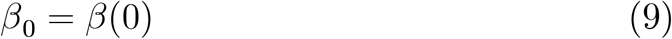

giving:

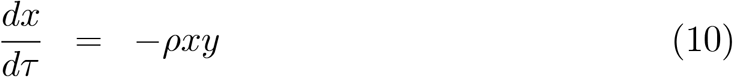

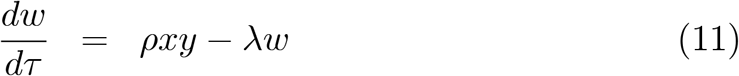

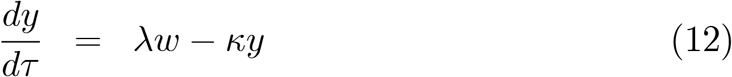

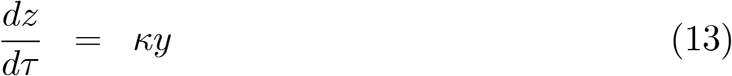

where

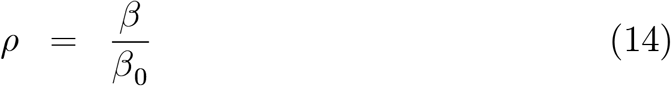

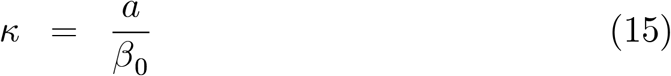

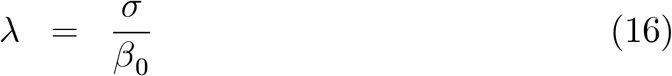

subject to

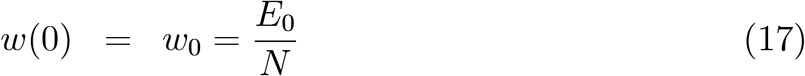

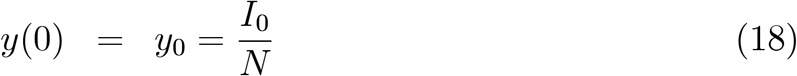

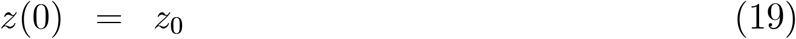

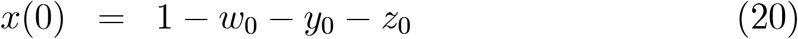

So, *ρ* and *k* are functions of *t*. If we apply the model with *t* = 0 corresponding to the beginning of the epidemic, then

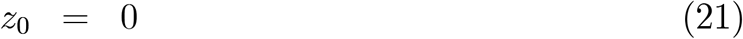

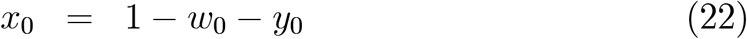

as there are no recovered at the beginning. Naturally occurring timescales are 1/*β*_0_, 1/*a* and 1/*σ*. We chose the 1/*β*_0_ time scale because it turns out to be the fastest in our application.

## Some features of the one-region SEIR model

### Effective reproduction ratio and basic reproduction ratio

The effective reproduction ratio *R*(*t*) and, it’s initial value, the basic reproduction ratio *R*_0_, are key parameters in the model. It can be shown (see Appendix A) that

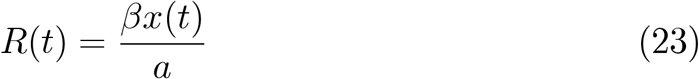

and, a fortiori,

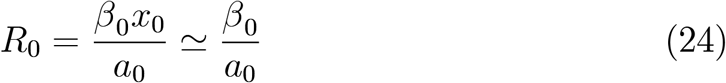

where

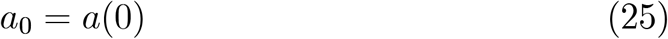

### Determination of the transmission/contact coefficient from other parameters

The early-time behaviour of the model can be studied by linearizing the system of differential equations (10) - (13) about the non-infective equilibrium state (*x, w, y, z*) = (1, 0, 0, 0). It follows that, initially, the number of infectives is a linear combination of two exponentials, one growing and the other decaying. After the decaying exponential has died out, exponential growth is possible for a further period during which the linearized equations are accurate. The substantial statistical variation in the model parameters, and, in particular, superspreading, also imply that this exponential growth does not occur immediately, but requires some initial spreading to take place [14]. These considerations suggest that after an initial transient and prior to government intervention, early exponential growth can occur. The formulae for the doubling time (*T*_1_) of the growing component and the half life (*T*_2_) of the decaying component are given by

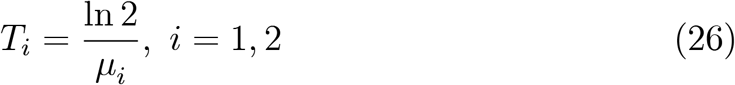

where *μ*_1_ and *μ*_2_ are the roots of the eigenvalue equation [4]:

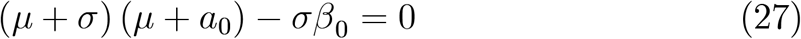

and are given by

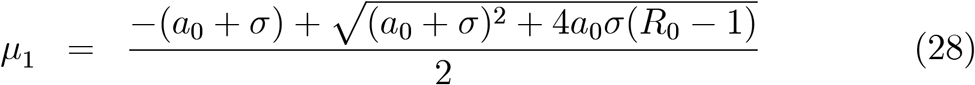

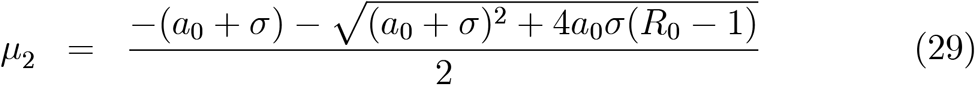

where *a*_0_, *R*_0_ and *β*_0_ are given by equations (25), (24) and (9), respectively. In order for the epidemic to grow, one of these eigenvalues must be positive. The condition for this is

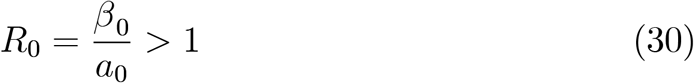

In this case

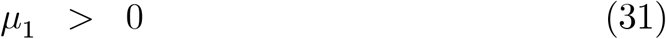

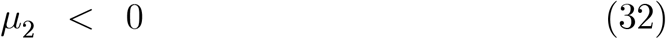

It follows from equations (27), (28) and (29) that

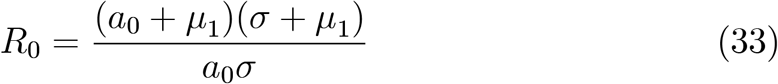

and

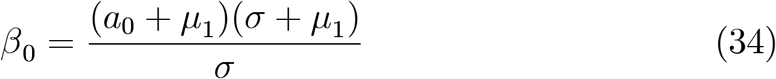

So, the parameters *σ, μ*_1_, *a*_0_ and *R*_0_ are related by equation (33) and cannot be chosen independently. In particular, if *σ, μ*_1_ and *a*_0_ are known, then *β*_0_ and, consequently, *R*_0_ are determined. We will estimate the values of *σ, μ*_1_ and *a*_0_ and then estimate *R*_0_ and *β*_0_ using equations (33) and (34).

## The Two-Region SEIR All-Island Model

### Model Equations

We model the interaction between the Northern Ireland and Ireland populations as being localized in a narrow strip of land straddling the border, the southern part being in Ireland and the northern part being in Northern Ireland, as illustrated in Figure 1. We number the regions as follows: Ireland is region 1 and Northern Ireland is region 2. The susceptible, exposed, infective, removed and total populations in region i are denoted by *S*_*i*_, *E*_*i*_, *I*_*i*_, *L*_*i*_ and *N*_*i*_, respectively, for *i* = 1, 2. For region *i*, the contact/transmission parameter is *β*_*i*_, the specific loss rate of exposed is *σ*_*i*_ and the specific removal rate of infected is *a*_*i*_, *i* = 1, 2. In specifying the values of parameters, we use the same notation as for the single region case, but add a subscript to indicate the region number. In particular, the single-region symbols *a, β*_0_ become *a*_*i*_, *β*_*i*0_, respectively, for region *i, i* = 1, 2. However, in a change of notation, we denote by *μ*_2_ the positive eigenvalue for region 2, corresponding to the positive eigenvalue *μ*_1_ for region 1 defined in equation (28).

**FIGURE 1.**
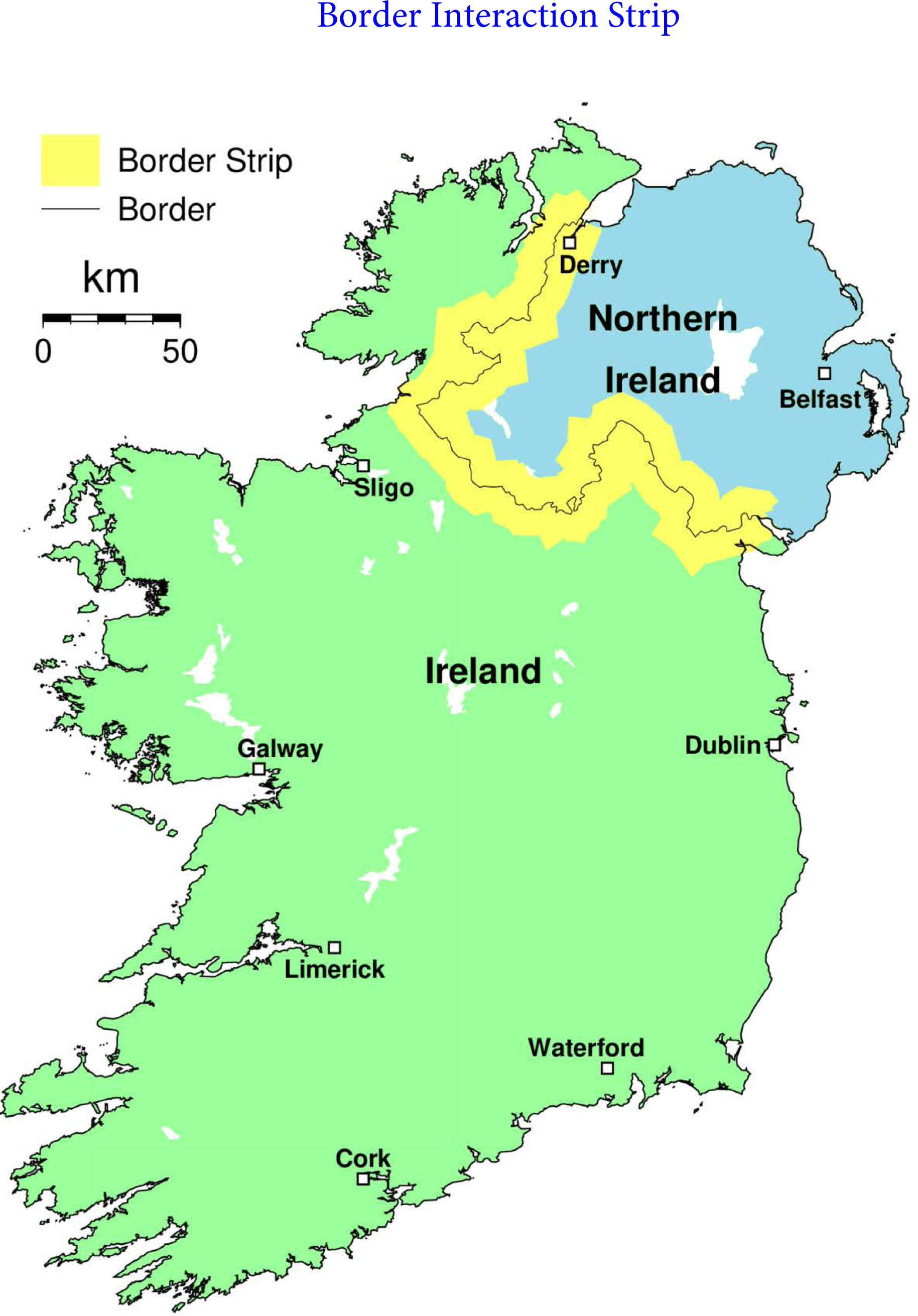

The population of that part of the border strip in region i is expressed as a fraction *a*_*i*_ of the population *N*_*i*_ in region *i*, for *i* = 1, 2. We did not incorporate interactions between people in Ireland and Northern Ireland due to travel between Belfast and Dublin via the M1 motorway and the towns easily accessible from the M1. Neither did we include travel between Northern Ireland and Ireland as a whole. These features could also possibly be included by increasing the *α* _*i*_, *i* = 1, 2 coefficients, and we did examine this (see Case Studies 8 and 9).

Susceptibles in either of the border strips can encounter infectives from both border strips. The interactions are indicated in Figure 2. The notation *S*_*i*_ − *I*_*j*_ in Figure 2 indicates an encounter between a susceptible from the border strip in region *i* with an infective from the border strip in region *j*. We assume that all interactions south of the border are in accordance with Irish government policies and laws and that interactions north of the border are in accordance with Northern Irish government policies and laws. The colours of the susceptible and infective symbols are intended to aid visualization of the regions from which interacting people come. The colours of the dashes indicate the region policies/laws according to which the interaction takes place. To keep notation simple in the derivation, we denote

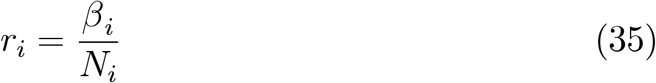

**FIGURE 2.**
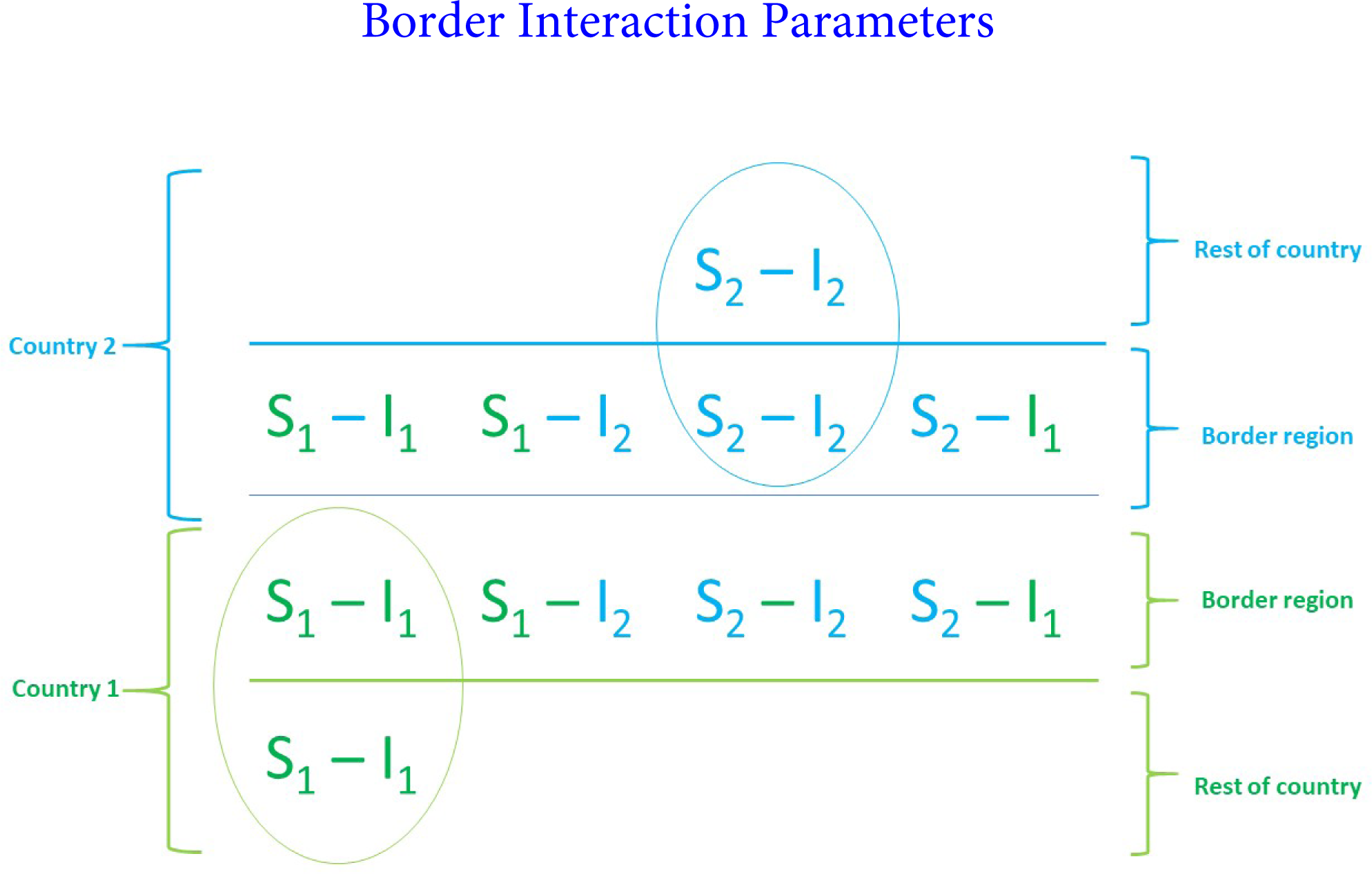

Susceptibles from region 1 can become exposed in four ways:

1. through encounters in region 1 with an infective from region 1. This results in a loss of *r*_1_*S*_1_*I*_1_ susceptibles per unit time.
2. through encounters in the border neighbourhood of region 1 with an infective from region 2. This results in a loss of *r*_1_ (*α* _1_*S*_1_) (*α* _2_*I*_2_) susceptibles per unit time.
3. through encounters in the border neighbourhood of region 2 with an infective from region 1. This results in a loss of *r*_2_ (*α*_1_*S*_1_) (*α*_1_*I*_1_) susceptibles per unit time.
4. through encounters in the border neighbourhood of region 2 with an infective from region 2. This results in a loss of r_2_ (*α*_1_*S*_1_) (*α*_2_*I*_2_) susceptibles per unit time.

Hence, adding the four loss rates

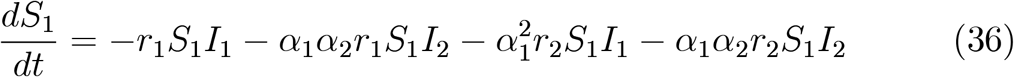

The first loss rate r_1_*S*_1_*I*_1_ term has been written down directly on the basis that all interactions between susceptibles in region 1 are homogenized. It can also be constructed by modelling that loss rate term as consisting of a sum of four loss rates involving interactions between susceptibles and infectives in the region 1 border region and the rest of region 1, viz:

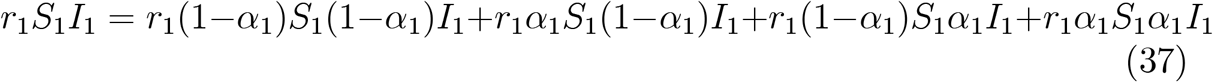

Similar considerations apply to the exposed from region 1 and the susceptibles and exposed from region 2. The resulting coupled SEIR model equations are as follows:

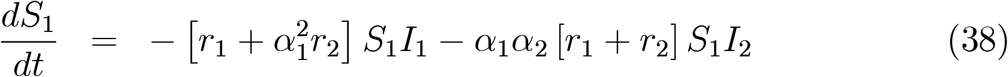

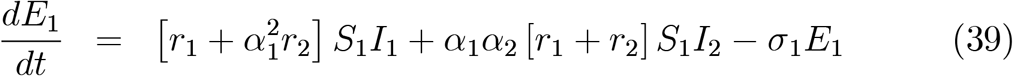

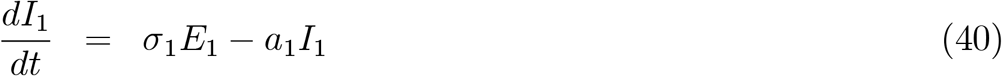

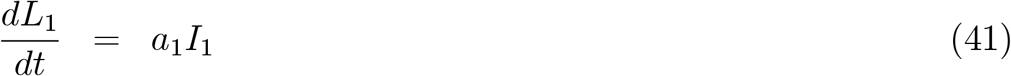

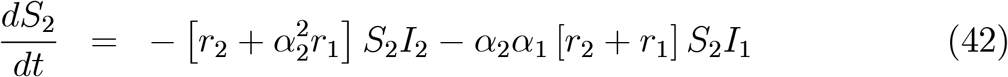

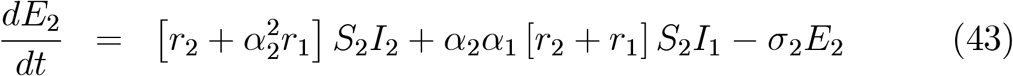

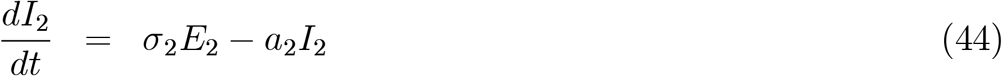

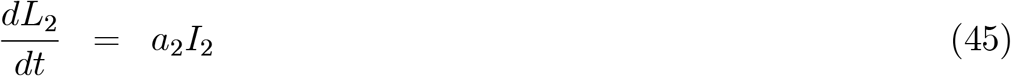

subject to the initial conditions:

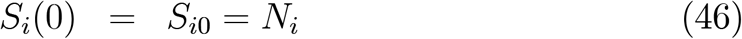

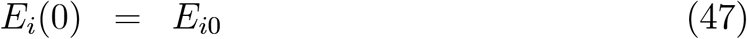

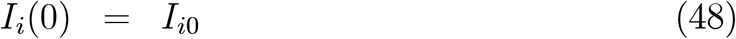

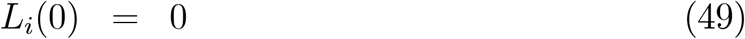

for *i* = 1, 2. It is assumed that the entire population of each region is initially susceptible, with the exception of a small number of exposed and/or infectives. It is assumed that there are no removed people in either region initially and that there will be small number(s) of exposed or infectives or both in each region. Note that *β*_*i*_ and *a*_*i*_ may be functions of *t* for *i* = 1, 2, following the comments on time-variation of government policies in the discussion on one-region models.

We scale and non-dimensionalise the equations as follows:

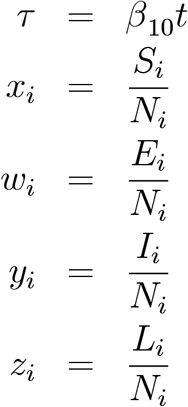

for *i* = 1, 2, where

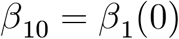

There are other natural times occurring in the problem, but we expect 1/*β*_10_ or 1/*β*_20_ to be comparable and substantially larger than 1/*σ*_*i*_ and 1/*a*_*i*_ in order for the epidemic to develop in the context under study. The scaled equations are given by:

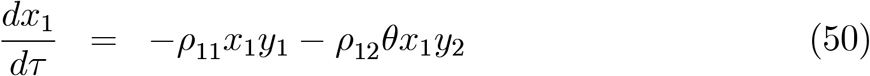

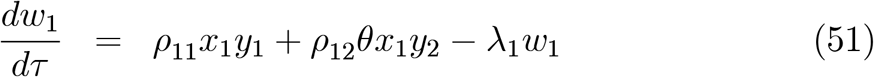

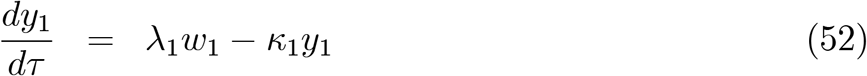

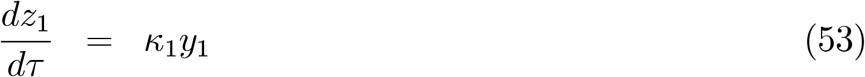

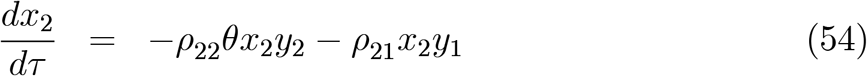

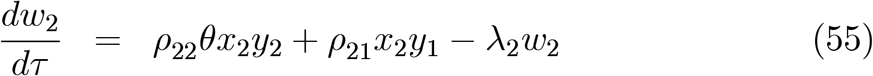

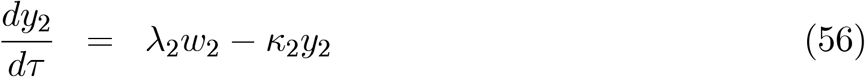

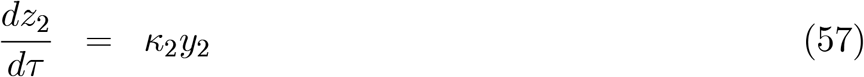

subject to initial conditions:

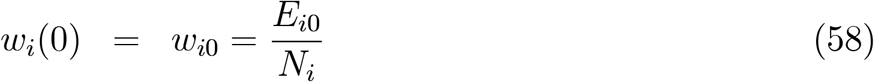

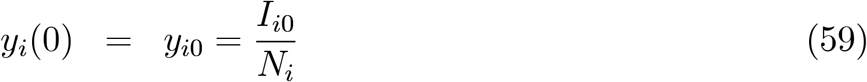

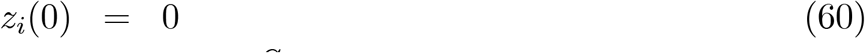

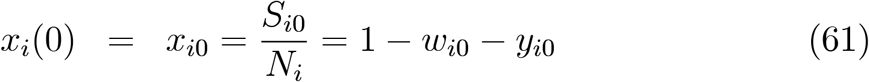

for *i* = 1, 2, and where:

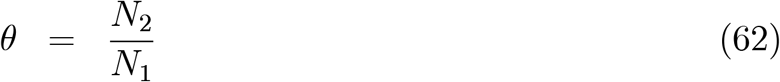

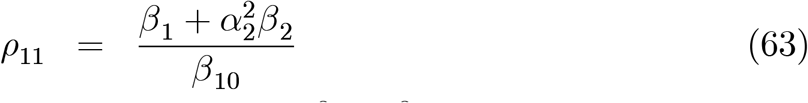

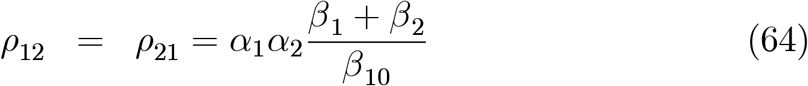

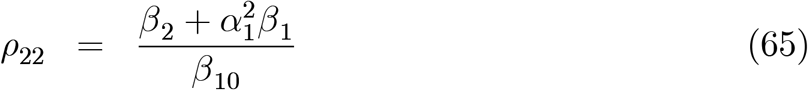

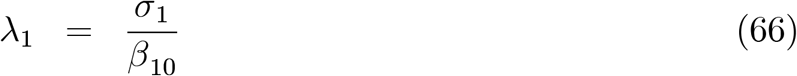

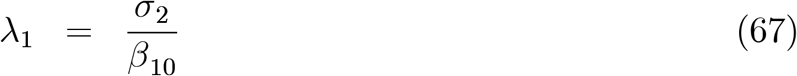

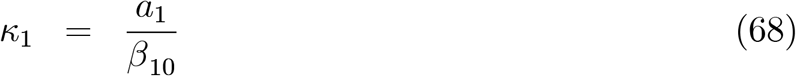

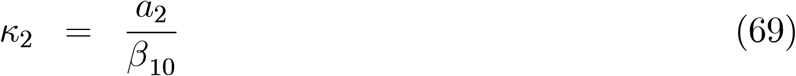

## Some Features of the two-region SEIR model

### Effective reproduction ratio and basic reproduction ratio

The effective reproduction ratio and its initial value, the basic reproduction ratio, are key parameters in the model. It can be shown (see Appendix A) that the effective reproduction ratios for infectives in region 1 and region 2 are given by

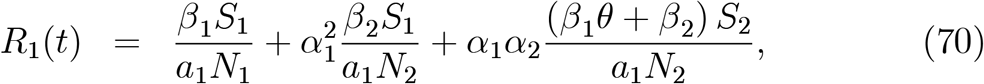

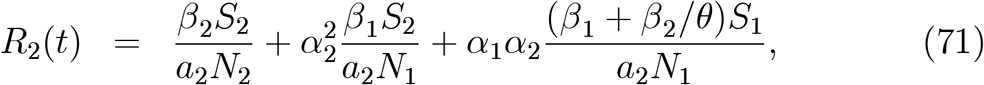

respectively and, hence, that the corresponding basic reproduction ratios are given by:

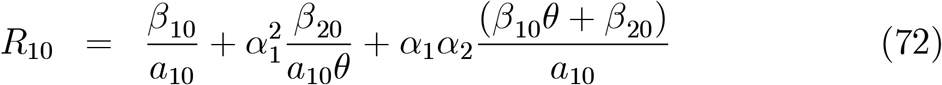

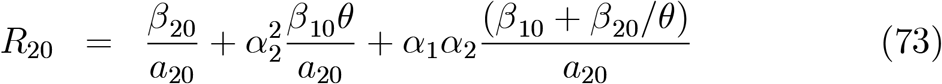

where

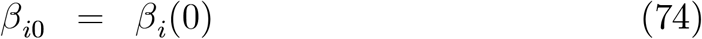

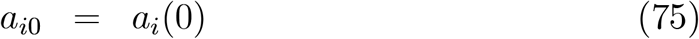

for *i* = 1, 2. If *α*_*i*_ ≪ 1, *i* = 1, 2, the effective and basic reproduction ratios are close to the one-region values - cf. equations (23) and (24). Note that *R*_*i*_ is defined to be the number of infectives on the island of Ireland produced by an infective from region *i, i* = 1, 2 and is different to its one-region counterpart (by a small amount if 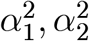 and *α*_1_ *α*_2_ are small in comparison with unity).

### Determination of the transmission/contact coefficients from other parameters

The initial growth in the two-region case is of interest because it enables estimation of *β*_*i*0_, *i* = 1, 2, and, hence, of *R*_*i*0_, *i* = 1, 2 in terms of other model parameters. We first linearize the system (38)-(45) about its disease free equilibrium:

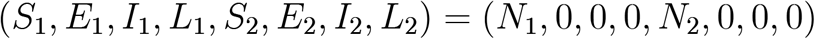

Let

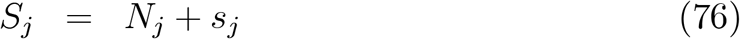

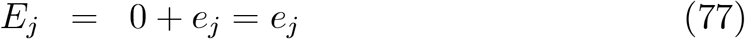

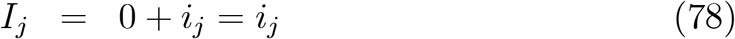

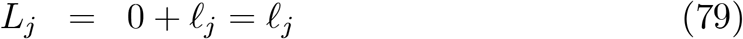

- the deviations of the variables from their disease-free equilibrium values. Note that *E*_*j*_, *I*_*j*_ and *L*_*j*_, *j* = 1, 2 are equal to their deviations *e*_*j*_, *i*_*j*_ and *l*_*j*_, respectively, from equilibrium. Taking account of the identities in equations (77)-(79), the linearized system is given by:

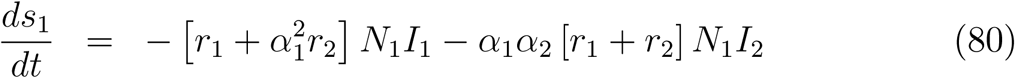

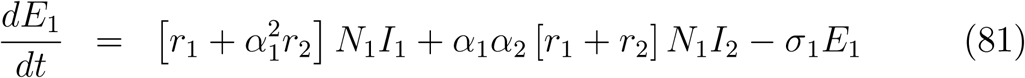

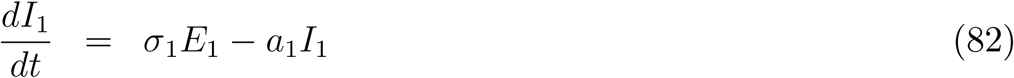

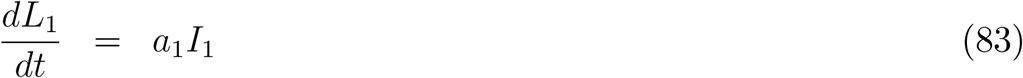

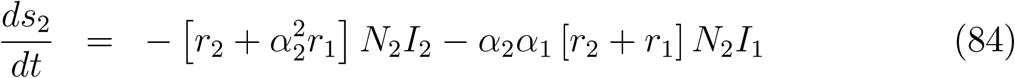

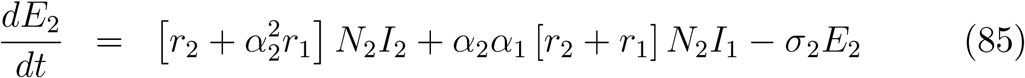

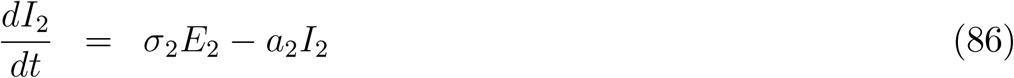

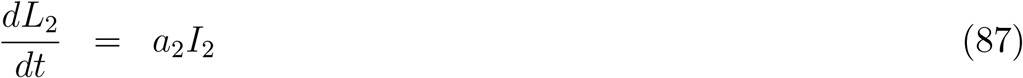

Equations (81), (82), (85) and (86) can be solved independently. They can be written in matrix-vector form:

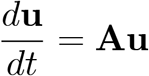

where

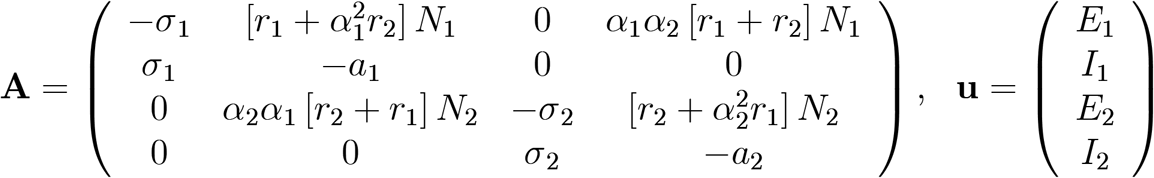

The eigenvalues of the matrix *A*, satisfy

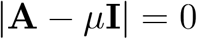

where *I* is the 4 × 4 identity matrix. This fourth order algebraic equation has the form:

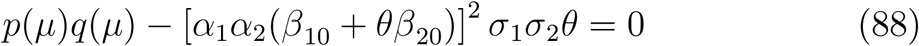

where

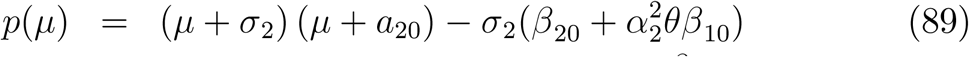

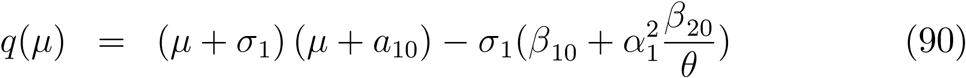

In the limit *α*_*i*_ → 0, *i* = 1, 2, the eigenvalue equation becomes:

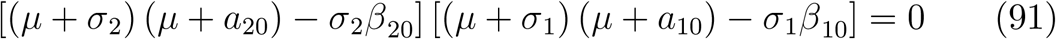

which is simply the product of the two one-region model eigenvalue equations (cf. equation (27)). Hence, it has two positive and two negative eigenvalues, provided

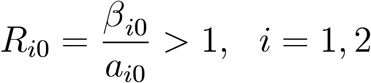

So, for *α*_*i*_, *i* = 1, 2 sufficiently small, and after an initial interval in which the negative exponentials decay to become negligible, a period of exponential growth should occur in each region and should be close to that predicted by the one-region model provided the eigenvalue equation (88) is not sensitive to small changes in the equation coefficients. On this basis, we would also expect the initial growth in each region to be dominated by a single exponential term (out of the two possible positive exponentials).

Let *μ*_1_ and *μ*_2_ be the positive eigenvalues of equation (88) where *μ*_*i*_ is the infective specific growth rate parameter for region *i, i* = 1, 2. Then *β*_10_ and *β*_20_ must satisfy the equations

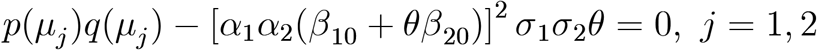

This system of equations has the form:

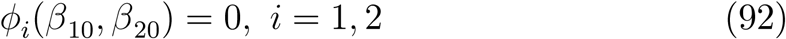

where

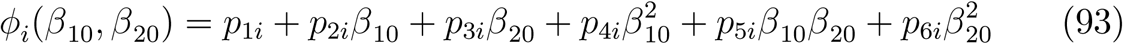

We find the solutions of equation (92) by first finding the curve *C* on which the surfaces

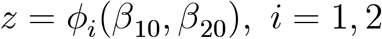

intersect. This curve can be parametrized in terms of *β*_10_ and is given by:

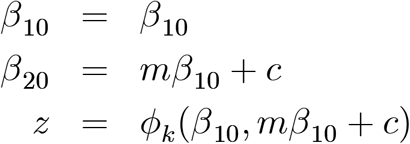

where *k* is either 1 or 2. The result is independent of *k* since *ϕ* _1_ = *ϕ*_2_ on the intersection curve *C*. The solutions of equations (92) then correspond to the points at which the curve C intersects the *z* = 0 plane. This, in turn, reduces to the solution of the quadratic equation

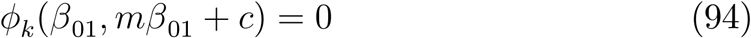

where

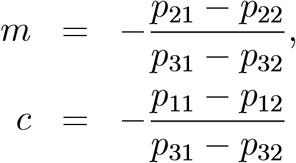

Thus, equations (92) do not uniquely determine (*β*_10_, *β*_20_) given the values of *σ, μ*_*i*_ and *a*_*i*_, *i* = 1, 2, unlike the result of a similar approach to the one-region case. Denote the two solutions of equation (94) by *β*_10*s*_, *s* = 1, 2 and let

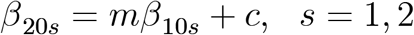

Both solution pairs (*β*_10*s*_, *β*_20*s*_) are functions of (*α*_1_, *α*_2_). As (*α*_1_, *α*_2_) → (0, 0), only one of the pairs tends to a pair corresponding to the one-region values given by equation (28) applied to each region. This is how we identified the appropriate solution pair (*β*_10*s*_, *β*_20*s*_).

So, we first estimated values of the parameters *σ, μ*_*i*_, *a*_*i*_ and *α*_*i*_, *i* = 1, 2 and then estimated the transmission/contact coefficients *β*_*i*0_, *i* = 1, 2 using the above procedure. Both the effective and basic reproduction ratios can then be estimated using equations (70) - (74), since values of *β*_*i*_(*t*) for *t* > 0 will be defined as piecewise constant multiples of *β*_*i*0_, *i* = 1, 2.

## Model Parameter Values, Government Interventions and Societal Responses

### Border interaction parameters

The border could have been closed at the beginning of the epidemic to enable greater control for both governments over their own regions. The border could also have been closed at any time during the epidemic. However, the Irish Government would be very reluctant to close the border [15], and some limited efforts have been made to enable cross-border cooperation [17], [16], [18]. For every case studied, we considered two border scenarios:

1. Border closed from the beginning of the epidemic
2. Border open throughout the epidemic

Another scenario worthy of study would be to consider the effect of closing the border at an intermediate time.

The border strip we envisage straddles the border and is about 20km wide, (see Figure 1 for an illustration). The Central Statistics Office of Ireland (CSO) carried out a study on the 10*km* wide strip on the Irish side of the border in 2016 [19]. They estimated the population in this strip to be 128, 000. We take it to be 130, 000 in our model. Based on looking at large towns in a strip 10*km* wide on the Northern Ireland side of the border, we estimated the population in this strip to be also about 130, 000. Within this 20*km* strip, some people on one side of the border work, carry out business, do shopping and socialize on the other. According to studies carried out by the Centre for Cross-Border Studies and the UK government on cross-border travel, over 30,000 people from one side cross the border daily to work in the other side [20], [21], [30] and there are are over 110,000,000 person border crossings annually, mainly by private vehicle, and nearly one million each of coach and train crossings annually [22]. These crossings would include M1 traffic and general Northern Ireland - Ireland travel, but we have not generated a specific estimate for these in terms of our model parameters.

We used the value *N*_1_ = 4, 900, 000 (following [23]). A recent more accurate value is available, viz. 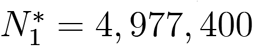, from the Irish Central Statistics Office [24]. We used the estimated value *N*_2_ = 1, 900, 000 for the Northern Ireland population. The most recent population value available for Northern Ireland is the mid-year 2019 value, which is 1, 893, 700 [25]. The 2020 population would exceed this if the population continued to grow at the 2018-19 historic rate [25].

We use the border strip populations to estimate the coefficients *α*_*i*_, *i* = 1, 2:

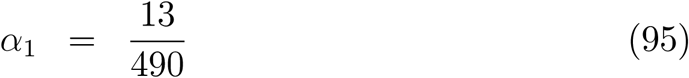

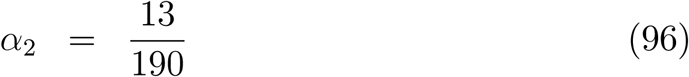

These values would have been very slightly smaller if we had used more accurate recent population estimates for Ireland and Northern Ireland. The average population density in Northern Ireland is significantly higher (2.3 times) than that in Ireland, since the land areas of Ireland and Northern Ireland are 70, 273 *km*^2^ [26], [27] and 14, 130 *km*^2^ [28], respectively. Also, the fraction of the Northern Ireland population in the northern border strip is higher than the fraction of the Ireland population in the southern border strip. The parameters *α*_*i*_, *i* = 1, 2 only enter into the model equations through their products. Note that

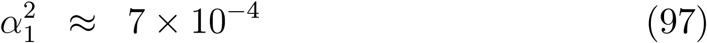

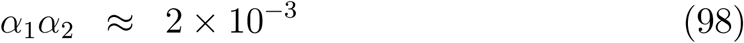

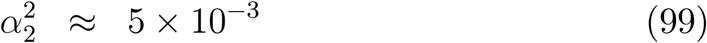

So, the border interaction parameters are all small in comparison with unity. We note that if 30,000 people cross the border daily to go to work, that would account for about 30, 000 × 5 × 50 = 15, 000, 000 border crossing per annum, which is much smaller than the 110, 000, 000 crossings estimated by the British government in a Brexit-related study of cross-border traffic [22]. We did one study in which we increased the values of the products *α* _*i*_*α*_*j*_ by a factor of 10, corresponding to increasing each *α*_*i*_ by a factor of 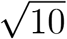 in an attempt to include all three possible cross-border interactions mentioned above. This enabled some assessment of the sensitivity to the values of the *α*_*i*_, *i* = 1, 2. It would be worthwhile to gather and study data on cross-border interaction more thoroughly.

## Exposed/Latent period parameters

In our model, we have defined 1/*σ*_*i*_ to be the average length of the latent period during which the exposed in region *i* are asymptomatic and not infective.

We assume that

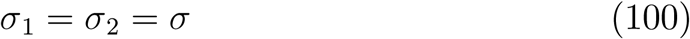

as we consider *σ* to be a biological constant, independent of government interventions or societal behaviour. We follow Li [9] and choose

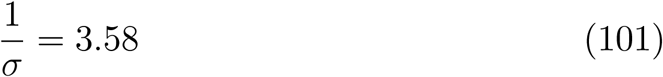

which gives

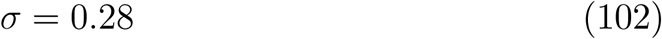

approximately. We note that shorter and longer values have been proposed and considered in models in the research literature [44], [35], [39].

### Infectious period parameters

For the average duration of the incubation period, we follow the WHO [31] and assume it to be about 5 to 6 days, noting that it can vary from between 2 and 14 days. This is not a parameter in our model, but it does provide information relevant to the choice of *α*_*i*_, *i* = 1, 2. Data is limited and variable on the duration of the infectious part of the incubation period [32], [7], [33], [1], [34], [35]. From our choice of incubation period duration and latent period duration it follows that we are assuming the length of the infectious presymptomatic part of the incubation period to be between 5 − 3.58 = 1.42 and 6 − 3.58 = 2.42 days, roughly. This indicates a lower limit on the duration of the infectious period. We chose a middle of range value of 2 days for the duration of the presymptomatic infectious period in our model.

How might people respond when they first exhibit symptoms? There would probably be a spectrum of response times, possibly conditioned on the severity of the symptoms. We took an estimate of an average of 2 days after exhibiting symptoms to self-isolate or self-quarantine, which would result in a roughly 4-day infectious period (including 2 presymptomatic infectious days) before self-isolating. So, we estimate the natural specific removal rate of infectives to be 1/4 = 0.25 per day. A key point is that, according to this model, the rate of removal of infectives through self-isolation or self-quarantine would be proportional to the number of infectives. We employ the CDC definition of isolation and quarantining [37]. This means that both suspected and confirmed cases are not in contact with others.

In addition to the natural social response causing removals via self-isolation, there would be a further removal process as a consequence of testing, tracing, identification and isolation/quarantine. Tracing involves checking all contacts of each infective ientified by a test. Ideally, the number of tests with associated tracing performed per day would be proportional to the number of infectives on that day. If the number of positives found per day in this way was a fixed fraction of the number of tests, then the removal rate of infectives due to testing would be proportional to the number of infectives. If applied in region *i*, this would effectively increase the value of *a*_*i*_ and, hence, shorten the duration of infectiousness in region *i*. However, the number of infectives each day is not known. Only the number of observed or ‘active’ infectives each day is known. The latter could be substantially less than former. The daily number of tests that have been carried out in Ireland since 18th March 2020 has been published [38]. No distinction is made between asymptomatic and presymptomatic/symptomatic infectives in official figures [38]. It would strictly be necessary to distinguish between asymptomatic infectives and presymptomatic/symptomatic infectives to estimate the parameters of the associated loss terms in the governing equations. There is very little data available on asymptomatic infectives [39], [40]. All of these considerations imply that modelling of elimination of both kinds of infectives through contact tracing/identification/isolation is challenging. Various simplifying assumptions could be made (see e.g. [23]). They would involve additional parameters and equations. In this work, we do not include the modelling of infectives reduction through contact tracing. One type of contact tracing reduction can be incorporated into the model: that for which the removal rate due to contact tracing is proportional to the number of infectives. We do not explicitly incorporate such a feature, but it could be investigated by looking at the consequences of increased values of the parameters *a*_*i*_, *i* = 1, 2.

There was little emphasis on testing/tracing/identification/isolation in the UK, which includes Northern Ireland, although it was eventually resumed several months after being terminated, as noted above. We assumed that societal self-isolation and self-quarantining would also be the dominant removal mechanism in Northern Ireland, and considered two cases; one for which the response time was the same as for Ireland and one for which it was a little slower (4.5 day infectious period, giving a specific removal rate of 1/4.5), to reflect a possible lower level of awareness due to the lower UK government emphasis on testing/identification/tracing/isolation/quarantining. We recognise that the values of *a*_*i*_ could be larger or smaller and that a range of values of *a*_*i*_, *i* = 1, 2 could profitably be investigated [39].

### Transmission/Contact parameters - Initial Values

Choosing values for *β*_*i*0_, *i* = 1, 2, reflecting initial social contact behaviour prior to government intervention, is difficult. The values do not seem to be directly measurable or intuitively obvious, as they embody multiple factors blended together into a single parameter: the frequency of contacts (direct and indirect) between susceptibles and infectives and the facility of transmission of the virus during encounters (depending on social distancing, mask wearing, COVID-19 related hygiene, fce touching, e.g.). It is no easier to choose a value for *R*_*i*0_, which depends on the values of both *β*_*i*0_ and *a*_*i*0_. Reported values of *R*_*i*0_ can vary quite a bit and have increased with time, from values little greater than 2 to as high as 7 [35], [36], [41]. We note, also, that values of the basic reproduction ratio do not seem to have been directly measured at all. It should be easiest to do so at the beginning, but that is a time when governments might not be prepared to act quickly in this respect [42]. So, basic reproduction ratio values are generally estimated through fitting models to data. The result depends on the quality of both the model and the data. There has been a tendency for reported values of the basic reproduction ratio to increase substantially in the light of better data on Wuhan, e.g. [35], [41], [32], [43], [44], [45].

We adopted the approach described earlier of estimating *β*_*i*0_, *i* = 1, 2 from the values of the parameters *σ, a*_*i*_, *μ*_*i*_ and *α*_*i*_, *i* = 1, 2. Assuming two positive and two negative eigenvalues when 0 < *α*_*i*_ ≪ 1, as occurs in this case, we sought to approximate the infective populations for the two regions using a linear combination of two growing exponential functions, on the basis that after an initial transient, the negative exponentials would have died out. Looking at the cumulative infectives data [46], [47], we could see that the data for days 9 to 15, inclusive, in Ireland and days 10 to 15, inclusive, in Northern Ireland, where day 1 is 1st March 2020, could each be approximated reasonably well by a single growing exponential function. The infective numbers for the two countries on these days are as follows:

| Number of Infectives each day |  |  |  |  |  |  |  |
| --- | --- | --- | --- | --- | --- | --- | --- |
| Day number | 9 | 10 | 11 | 12 | 13 | 14 | 15 |
| Ireland | 24 | 34 | 43 | 70 | 90 | 129 | 169 |
| Northern Ireland | - | 16 | 18 | 20 | 29 | 34 | 45 |

We also looked to see if a linear combination of two exponential functions (with unknown specific growth rates) could better approximate the data in each region using a direct least-squares fit in each region. We also jointly approximated the data in the two regions using a linear combination of the same two exponential functions with unknown specific growth rates. Examining the different approaches, we felt the best approach was to seek the best single exponential function approximation to the data in each region. Having done this, we now had two positive specific growth rates determined from the data. We denote these by *μ*_*i*_, *i* = 1, 2. The values of *μ*_*i*_, *i* = 1, 2 that we obtained from the least-squares data fits were:

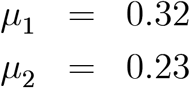

approximately, corresponding to initial doubling periods of

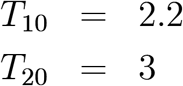

days, approximately, respectively. The daily number of active cases [48] in Ireland differed little from the daily number of cumulative cases in the first few weeks. So, using the daily active rather than the daily cumulative data makes only a minor difference. We only had access to the cumulative daily data for Northern Ireland and assume the differences between cumulative and the active daily infective values were also negligible during the first few weeks in Northern Ireland.

We note that the value for the initial doubling time for Northern Ireland is somewhat anomalous. The initial infective doubling time for Ireland, the UK as a whole, and for Italy, France, Germany and Spain were all close to 2 days [29]. So, in order to err on the site of caution, we also carried out simulations with a doubling time slightly greater than 2 days for Northern Ireland. We used

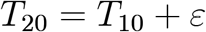

with *ε* = 0.001. We chose slightly different values because our algorithm to find *β*_*i*0_, *i* = 1, 2, requires *μ*_1_ ≠*μ*_2_.

### Transmission/Contact Parameters Intervention values

There were several substantial contact/transmission reduction measures at various times taken in Ireland and Northern Ireland [12], [13]. The first major intervention for both countries was closing schools and third-level colleges. This Northern Ireland measure was implemented a week later than the Ireland measure. Measures such as these and later relaxation measures signify that *β*_*i*_, *i* = 1, 2 are functions of time. We did not seek to model every transmission/contact reduction measure in each region. Rather, for simplicity, we modelled transmission/contact reduction as an instantaneous single reduction of *β*_*i*_ in each region, with the reduction in *β*_2_ timed to be later that for *β*_1_ in most cases. We chose the reductions so that the effective reproduction ratios, *R*_*i*_(*t*), *i* = 1, 2, would eventually become less than unity in both regions. We also considered cases in which the reduction in *β*_2_ was such that *R*_2_ remained a little above 1 while *R*_1_ had dropped below 1, to allow for possible uncertainty in the estimation of *R*_2_(t) in Northern Ireland, in order to err on the side of caution in considering policy implications for the Government of Ireland. We examined one case where a similar approach is taken for policy implications for the Government of Northern Ireland on account of the approximate nature of the data and parameter values relating to Ireland.

We also looked at an additional change (upwards) in *β*_2_, or in both *β*_1_ and *β*_2_, together, to represent easing of transmission/reduction in one or both regions. These were chosen to be instantaneous increases in *β*_*i*_.

So, the functions *β*_*i*_ had the following forms:

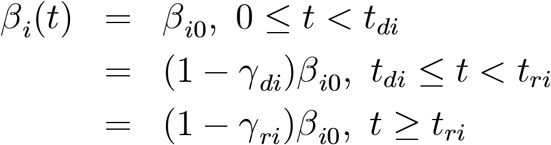

for *i* = 1, 2, where *γ*_*di*_ and *γ*_*ri*_ are fractional changes in transmission/contact coefficients from their initial values and *γ*_*ri*_ < *γ*_*di*_.

### Border Status - Coordination

We also examined a case where as many parameters as possible were equal in both regions to examine the effect of the border on coordinating policies north and south.

### Asymptomatic infectives

We did not include asymptomatic infectives in our model. Such infectives would presumably have a latent period, but would subsequently remain asymptomatic when the asymptomatic person becomes infective. Unfortunately, there is very little data available on asymptomatic transmission [39], [40]. There are varying estimates of the fraction of infectives that are asymptomatic [39]. It is possible that as many as 80% of infectives are asymptomatic [50]. The infectiousness of asymptomatics relative to presymptomatics/symptomatics is not well-understood [39], [40]. They would not self-quarantine on account of not being aware they had contracted the disease. So, they could be expected to be infective for a substantially longer period than the infectives who become symptomatic.

Introduction of asymptomatics into the model equations would add at least three extra parameters (contact/transmission, fraction of exposed who become symptomatic the remainder being asymptomatic, and duration of infectiousness) for a single-region model and six parameters for a two-region model. We did not include asymptomatics in order to keep the number of parameters small to facilitate exploration of parameter space. We reasoned that any important behaviour discovered in our simple model could be investigated for more complex models that would include asymptomatics and contact tracing.

### Case Studies

We carried out 13 case studies to examine the effects of various policies and societial responses in the context of border status (open or closed). They are described in detail in Appendix C.

Case Studies 1, 2 and 4 comprise scenarios involving just one intervention (transmission/contact reduction) for which an open border leads to infective peaks that are higher than those in the corresponding closed border cases. The increase is modest in one region (2%) and larger in the other (5% or 8%). The open and closed border peak widths in both regions are comparable in all three cases. The larger increase can occur in either region.

Case Studies 3, 5, 6 and 7 all involve similar effects with different causes. In all cases, the progress of the epidemic in Northern Ireland causes very substantial changes in the progress of the epidemic in Ireland, but without any obvious cause if looked at from the perspective of an Ireland model not incorporating an interaction with Northern Ireland. In cases 3 and 5, the effective reproduction ratio in Ireland, *R*_1_(t), remains near 0.5 suggesting the epidemic is being well controlled. In cases 6 and 7, *R*_1_(*t*) remains below but close to 1. In cases 3 and 5, an increase to a second infective peak is completely unexpected in the context of a one-region model. In cases 5 and 7, the increase to a second peak is much faster and steeper. If there was not an awareness of these negative effects due to pursuit of different policies or societal responses in the two regions, there could be a delay in trying to come to terms and cope with the effects. In all of cases 3, 5, 6 and 7, the source of the problem is not in the region in which it occurs, but, rather, the other region.

In Case Studies 8 and 9, we look at the effect of the size of the border interaction parameters on the evolution of the epidemic. We base these studies on Case Studies 4 and 6, respectively. We noted above that our chosen values for *α*_*i*_, *i* = 1, 2 were based on the interactions occurring in a narrow strip of land enclosing the border. We noted that the M1 motorway corridor and general Northern Ireland-Ireland travel could be other sources of a similar interaction. These coefficients enter the equations as products. So, we increased the size of each coefficient by a factor of 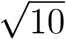 to give an order of magnitude increase in the border interaction coefficients in the equations. In Case Study 8 (based on Case Study 4), the infective peaks increase in magnitude substantially (dramatically in Northern Ireland). In Case Study 9 (based on Case Study 5), the effects observed in Case Study 6 are intensified dramatically. These results suggest it would be worthwhile to try to better quantify the M1 corridor and general Ireland - Northern Ireland travel to get better estimates of the interaction parameters.

We found a number of cases of unstable behaviour in the model. Some examples are considered in Case Study 10. In one case a very small change of the order of 2.5% in *γ*_*di*_, *i* = 1, 2 causes a dramatic (1000%) change in the height of the infective peak in Ireland. In another example, a change of 2.7% in the value of 1/a_2_ causes a 13% increase in the height of the infective peak in Northern Ireland and a 162% increase in the time of occurrence of that peak.

In investigating the possibility of an all-island approach by designing policies to make as many parameters as possible equal in Case Study 11, it turned out that the Northern Ireland infective peak in the scenario considered was about 2.5 times that in Ireland. Possible sources causing this were the values of *α*_*i*_, *N*_*i*_ and *y*_*i*_(0) = 1/*N*_*i*_. It turned out that the source of the disparity was not the values of a_*i*_, but, rather, the values of the initial conditions, which are tiny:

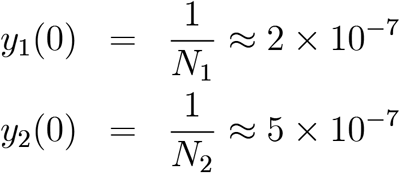

Case Study 12 shows the results obtained for a range of values of y_1_(0), keeping y_2_(0) = 1/*N*_2_. For y_1_(0) = 1/*N*_2_, the infectives curves for the two regions are quite close. The epidemic curves in both regions are then quite close. This highlights two factors: the initial conditions were not under the control of the governments; so, the difference is real; one could explore the model to see if it could be overcome by policy; also, there is a sensitivity to small changes in initial conditions in the model. The latter is a third type of instability. These instabilities are important. If there was not an awareness of them, certain large changes in numbers of infectives in either region could come as a surprise.

Case Study 13 is similar to Case Study 5 with the roles of Ireland and Northern Ireland reversed. In this case 1/*a*_2_ = 4, 1/*a*_1_ = 4.5. The intervention times are the same as for Case Study 5 and the sizes of the interventions in region 2 for Case Study 13 are the same as those for region 1 in Case Study 5 and vice versa. The conclusions are similar to those in Case Study 5 with roles reversed. In this case, a single region model for Northern Ireland would predict a single peak with *R*_2_ substantially less than 1 after the single Northern Ireland contact/transmission reduction intervention, whereas the two-region model predicts a second large peak in Northern Ireland caused entirely by the evolution in Ireland. Thus, the second peak in Northern Ireland would come as a surprise in the context of a single region model, again indicating the wisdom for both Governments of employing a two-region model for the island of Ireland in order to be aware of what can happen.

## Conclusion

In this study, we did not seek to model the historical evolution of the islandwide epidemic, but focused on a simple model to see if it exhibited any pronounced difference between the predictions of one-region models for Ireland and Northern Ireland and a two-region all-island model incorporating border interactions in a variety of simple scenarios. We found that substantial differences between the one- and two-region models could occur. In addition, the negative effects prediced by the two-region model would not be predicted or be explainable by the one region models.

In our exploration of parameter space for our simple two-region model we discovered sensitivities to small changes in values of key parameters, the values of which are determined by choices of policies intended to control the epidemic in the two regions. Large changes corresponding to small changes in certain parameters could occur for certain parameter ranges and not for others. Awareness of such features of the model could be helpful to those formulating policies to contain the epidemic.

We found that the pursuit of different policies in the two parts of Ireland, in the presence of an open border, could result in substantial negative effects on either side of the border, indicating that North-South cooperation on policy would be valuable in avoiding possible unnecessary and unanticipated high levels of infectives in the two regions.

Our study of the effects of the size of the border interaction parameters suggests better quantification of the interactions including more general travel-related North-South interactions would be valuable. Our interaction model could also possibly be employed to assess travel interactions between Ireland and other countries.

We did not include asymptomatic infectives in the models. Nor did we include government testing/tracing. The various substantial interactions between the two regions, due to policy/societal response differences, and sensitivities to small changes in policy/societal response suggest looking to see if such phenomena also occur in two-region models incorporating asymptomatics and testing/tracing.

## Data Availability

All data referenced are contained in the references quoted in the manuscript. No other data was referenced.

## Acknowledgements

The authors would like to express their appreciation to Professor Chris Bean, Director, School of Cosmic Physics, Dublin Institute for Advanced Studies for releasing James R. Grannell from duties at DIAS to work on this project and for making DIAS computing facilities and software available to facilitate the work, to Dr Claus Koestler, School of Mathematical Sciences, UCC, for suggesting the idea of looking at the pandemic on an all-island basis, to Professor Gerard Killeen, School of Biological Sciences, UCC for sharing his extensive experience of pandemics in Africa, for assuaging our concerns about finding larger values for basic reproduction ratios in our model and for his encouragement of this work, to Professor Denjoe O’Connor, Director, School of Theoretical Physics, Dublin Institute for Advanced Studies for providing valuable information on epidemiological approaches to modelling and making helpful suggestions in relation to the focus of the work, to Dr. Phillipp Hoevel, School of Mathematical Sciences, UCC, for helpful discussions and providing insights on epidemiological thinking, to Professor Patricia Kearney, School of Public Health, UCC for connecting us to important current research on COVID-19, and to the UCC COVID-19 Research Group/UCC Data Analysis and Modelling Support Group for helpful discussions and support.

This work was undertaken to support the Irish Epidemiologal Modelling Advisory Group (IEMAG) in its role of providing mathematical modelling support and advice to the Chief Medical Officer of Ireland and the National Public Health Emergency Team (NPHET). This is an expanded version of a report which was submitted to IEMAG on 10 June 2020. The authors are grateful for the sharing of knowledge, encouragement and feedback provided by members of IEMAG.

## Appendix A Effective and Basic Reproduction Ratios

### SEIR one-region model

The model differential equations are

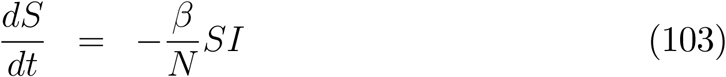

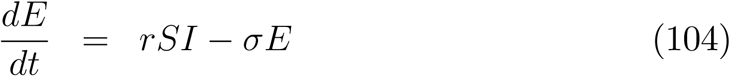

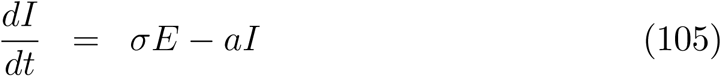

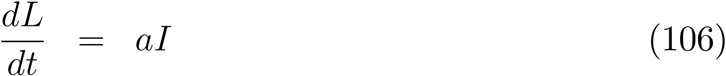

There are two classes of people who have contracted the disease: exposed and infectives. let us call them the infected. Infectives create only exposed. So, the question is: how many exposed does each infective create? The number of new exposed per unit time is *rSI*. So, the number of new exposed per unit time created per infective is *rS*. An infective is infectious for 1/*a* time units. Hence, the total number of new exposed created by each infective is

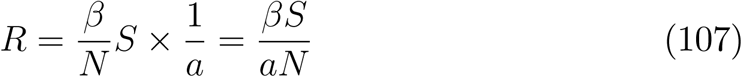

This is the effective reproduction ratio and varies with time. If *R* > 1 the epidemic will get worse. This can also be deduced directly from the differential equations - add equations (104) and (105) to get

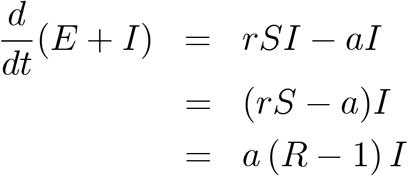

So, the number of infected will increase if *R* > 1 and decrease if *R* < 1.

Finally, the initial value of the effective reproduction ratio is the basic reproduction ratio:

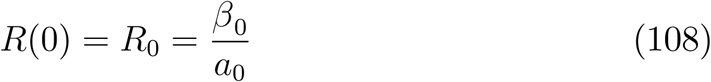

since *S*_0_ = *N*. The basic reproductive ratio can also be computed using the next generation matrix approach [49], [3]. The *F* and *V* matrices, in this case, are:

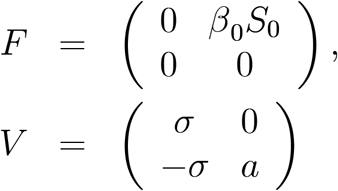

and *R*_0_ is the spectral radius of *FV* ^−l^, which gives the same result as that in equation (108).

### SEIR two-region model

The model differential equations are (38) - (45) and (35), repeated here for convenience:

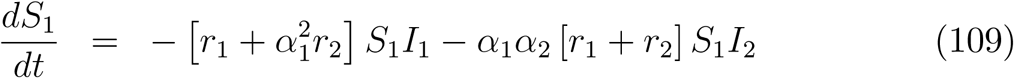

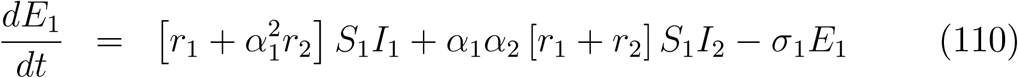

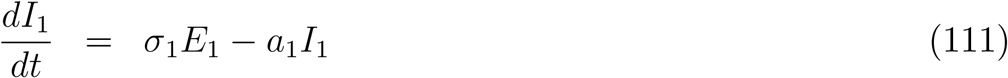

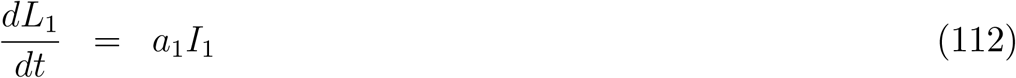

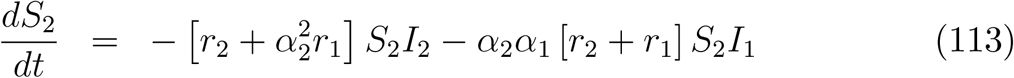

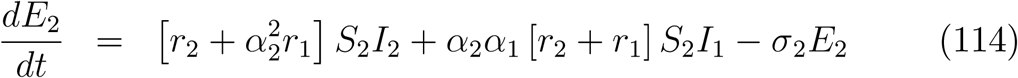

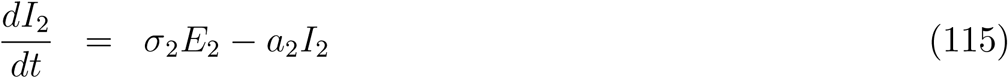

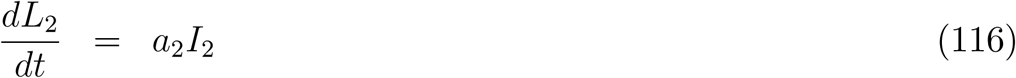

and

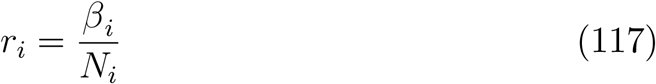

Consider infectives in region 1. They create new exposed in both countries. The total number of exposed created per unit time by region 1 infectives is:

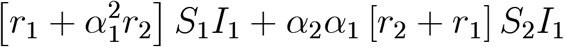

The exposed are the newly created infected. Hence, the number of infected created per region 1 infective per unit time is

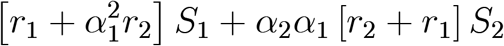

The region 1 infectives are infectious for 1/*a*_1_ time units. Hence, the number of new exposed created by a region 1 infective is

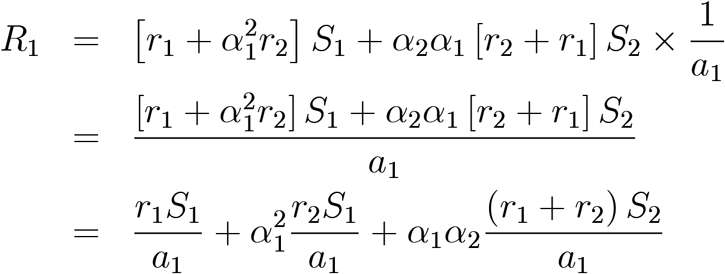

*R*_1_ is the effective reproduction ratio for region 1 infectives. Similar considerations apply to region 2 infectives infecting susceptibles in both countries to create newly infected (exposed) and the effective reproduction ratio for region 2 infectives is

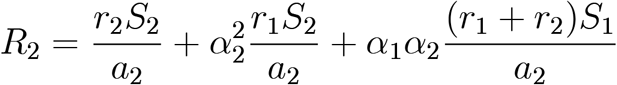

*R*_1_ and *R*_2_ can be expressed in terms of 6_*i*_ instead of r_*i*_, using the relationship in equation (117) to give:

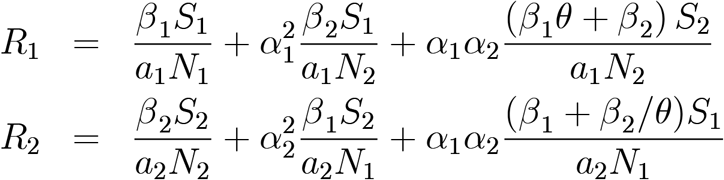

where *θ* is given in equation (62). So, the effective reproductive ratios for the two-region model differ only a small amount from the corresponding values for the respective single region models. The corresponding basic reproduction ratios are the initial values of the effective reproduction ratios:

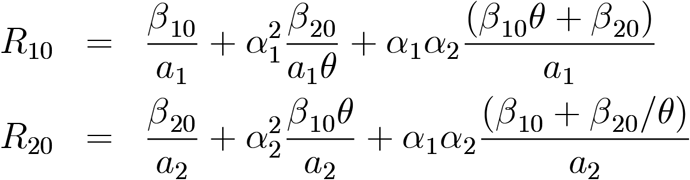

## Appendix B Case Studies

Each case study was focused on determining if the status of the border (open or closed) in the model could significantly affect the evolution of the epidemic in either or both regions. We also examined if the value of the initial doubling time in Northern Ireland significantly affected the predicted evolution in either or both countries. We considered two values: that obtained from the data for days 10 to 15 (viz.: 3 days approximately) or a doubling time the almost the same as in Ireland (about 2.2 days). We examined a range of possible transmission/contact reduction times for both countries and settled on two pairs. The chosen intervention needed to be substantial in order to reflect the substantial lockdowns in both regions and in order to get the effective reproduction ratios below unity. This meant that the interventions could not be too early or too late. Late interventions could lead to huge peaks and early interventions could lead to very small peaks. We choose two pairs of transmission/contact reduction intervention times:

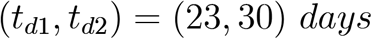

and

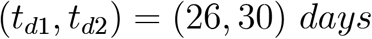

The fractional reductions were about 0.9 for Ireland and between 0.75 and 0.8 for Northern Ireland to reflect a lesser emphasis on transmission/contact reduction in the UK as a whole. The latter choice was made for the Irish Government to err on the side of caution. We also chose different values for the infectious period duration in Ireland and Northern Ireland: 4 and 4.5 days, respectively, again reflecting lower emphasis on testing/identification/tracing in Northern Ireland, again to err on the side of caution. We did consider some cases with similar parameters for both regions. We also considered one case with parameters for the two regions interchanged (Case Studies 5 and 13).

For each case study we list the values of the model parameters in the following way:

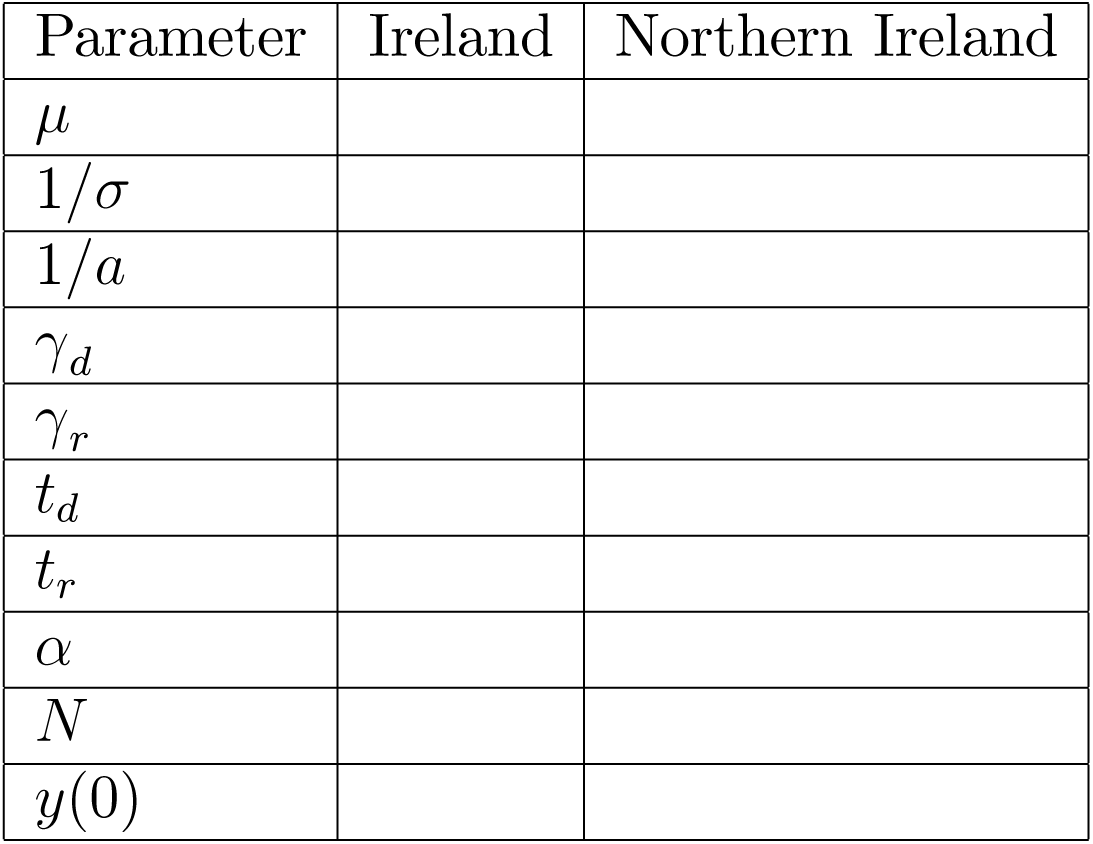

We omit the region subscripts 1 or 2 from the parameter names for simplicity of presentation. All of the above parameter values are independent of the border status open or closed, with the exception of *α*_*i*_, *i* = 1, 2. For most of the case studies, the same set of values of *α*_*i*_ and *y*_*i*_(0) are used, except for those case studies (8, 9 and 12) that explore the influence of the values of these parameters on the outcome. The values of *α*_*i*_ are listed explicitly only in the parameter tables for which variation of these parasmeters is studied. Parameters not arising in a particular study are not listed by default.

The derived parameter values *β*_0_ and *R*_0_ do depend on the border status and are determined from other parameters in the above parameter table, as explained earlier. The values of these parameters are also displayed in a separate table:

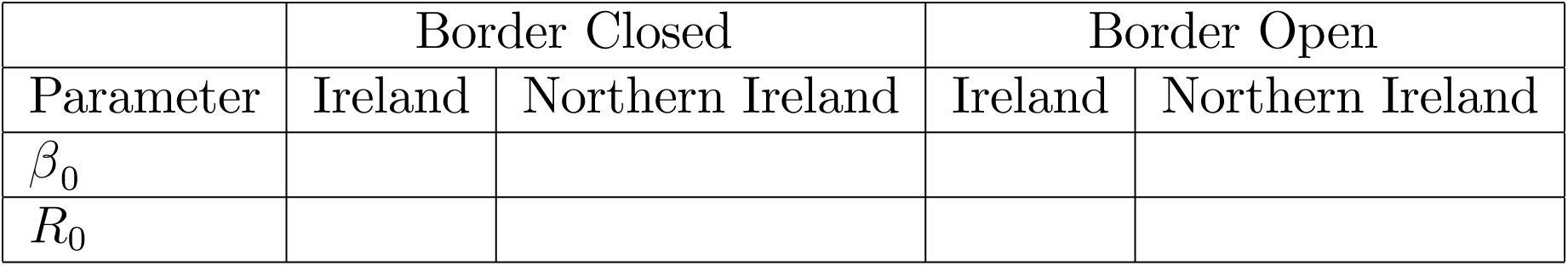

Most case studies are accompanied by two figures (one for each region), each containing two graphs showing the infective population versus time in the open and closed border cases and the effective reproduction ratio versus time in the open and closed border cases.

The values of *α*_*i*_, *i* = 1, 2 are as specified earlier:

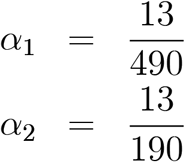

in all cases with the exception of two in which the sensitivity of the evolution to the values of *α*_*i*_, *i* = 1, 2 are studied.

We make a few abbreviations to fit the tables into the page:

1. Min - minimum
2. Diff - magnitude of difference

### THE COMPUTER CODE

The results presented in this report were computed using Wolfram Mathematica 12 [51]. The initial value problems for the open and closed border cases were solved using the standard NDSolve algorithm or NDSolve with the BDF solver option (which employs implicit backward differentiation formulae with orders 1 through 5).

#### CASE STUDY 1

The parameters for this study were as follows:

| Parameter | Ireland | Northern Ireland |
| --- | --- | --- |
| $\mu$ | 0.319512 | 0.227221 |
| $T_0$ | 2.16939 | 3.05054 |
| $1/\sigma$ | 3.58 | 3.58 |
| $1/a$ | 4 | 4.5 |
| $\gamma_d$ | 0.88 | 0.84 |
| $t_d$ | 23 | 30 |
| $t_r$ | - | - |

There was a single transmission/contact reduction intervention in each region. The derived parameters were as follows:

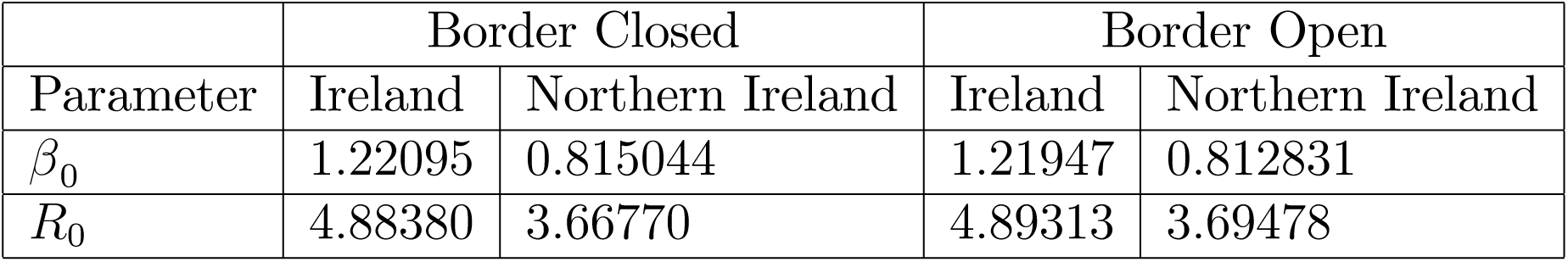

The infective population and the effective reproduction ratio versus time are shown for both the open and closed border cases for Ireland in Figure 3A. The corresponding results for Northern Ireland are shown in Figure 3B. The difference between the open and closed border infective peak population is substantially greater for Northern Ireland than for Ireland. In the context of the model aproximations, these are not large differences. The times of occurrence of the peaks are the same for both regions.

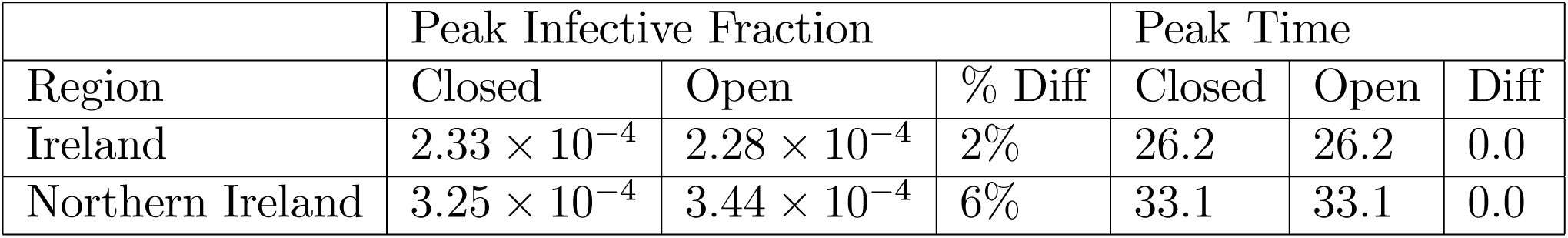

**FIGURE 3A.**
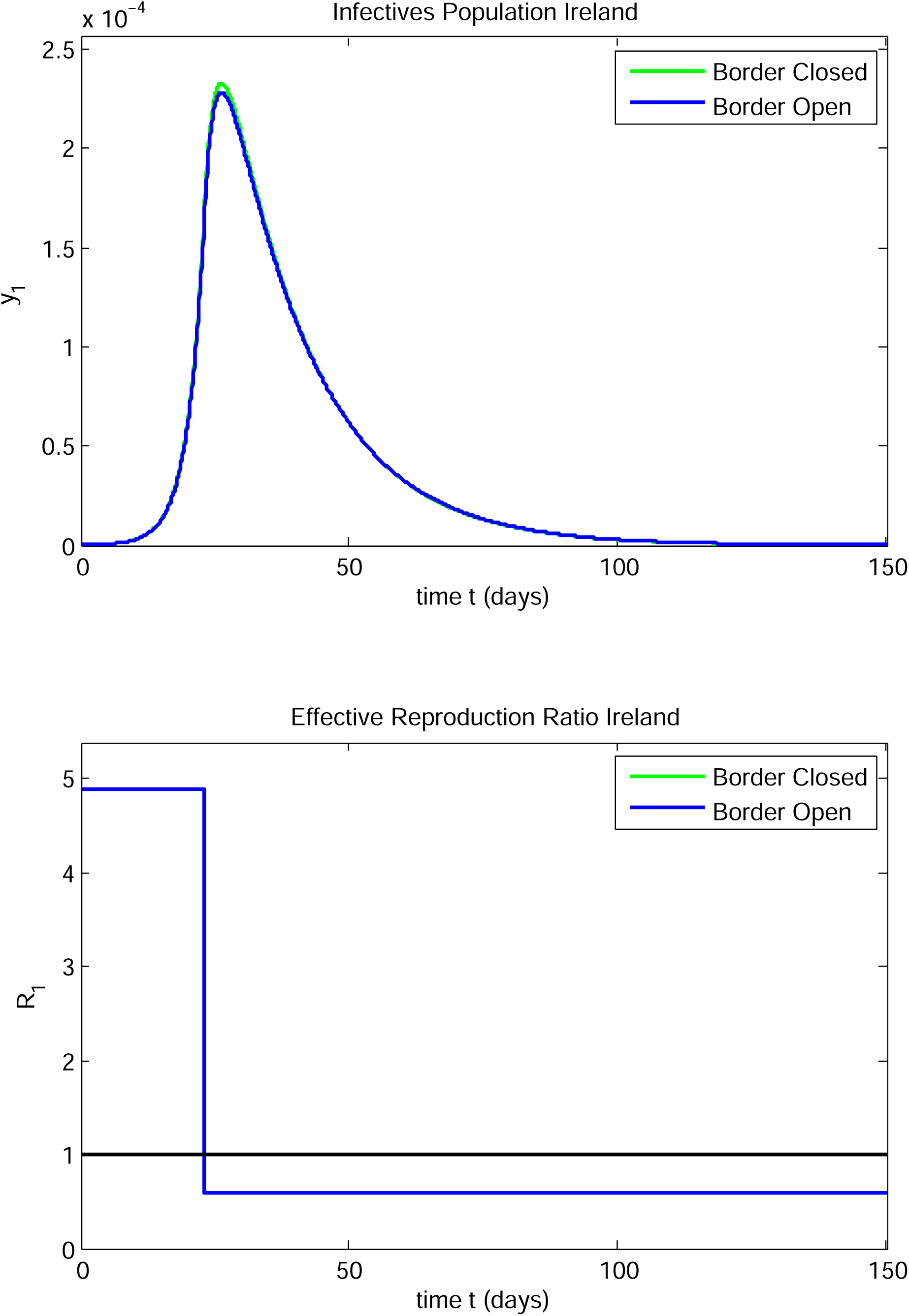

**FIGURE 3B.**
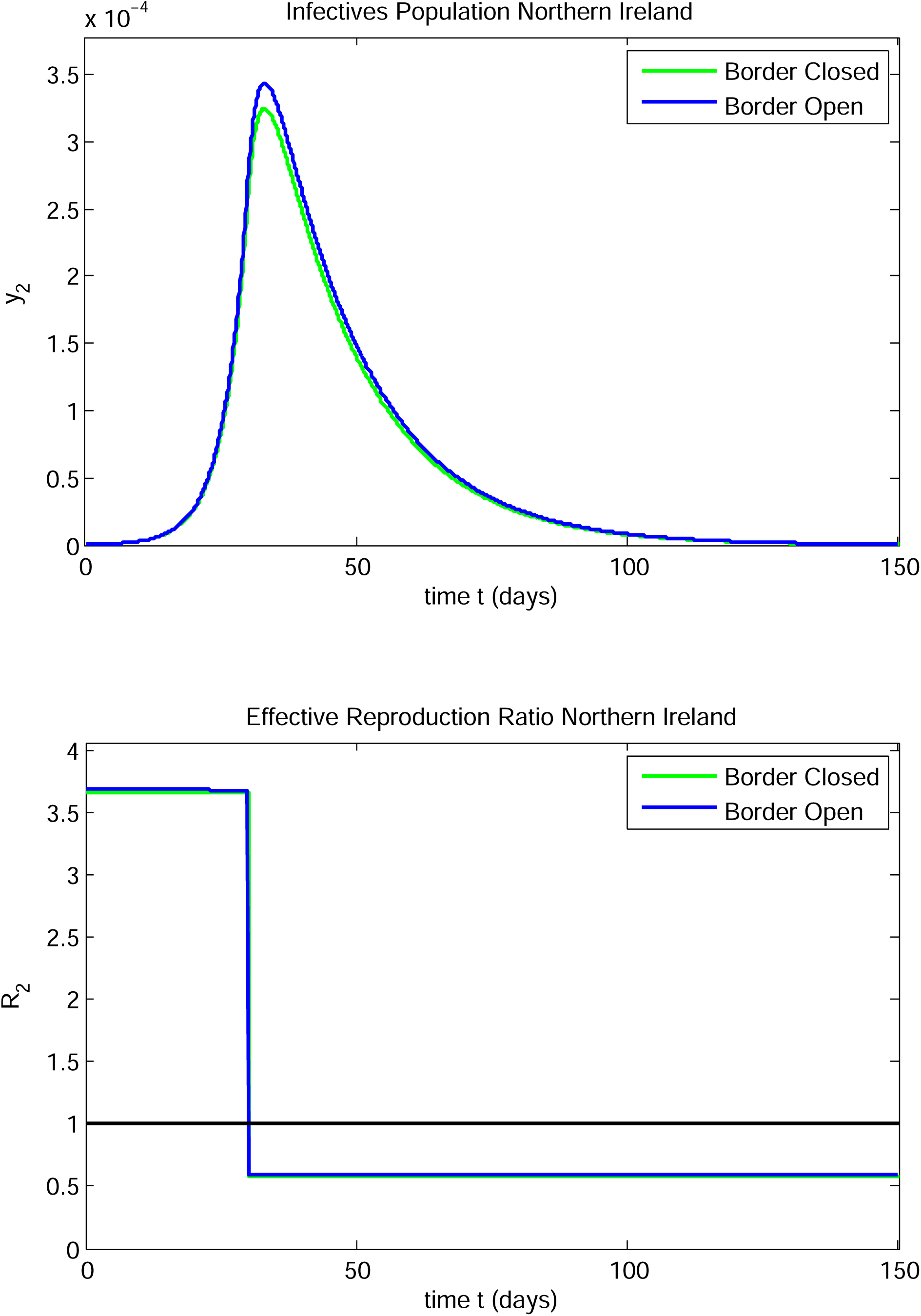

For both regions, the effective reproduction ratio falls to slightly under 0.6 post-intervention. The fraction of infectives rises rapidly and decays relatively slowly in both regions.

#### CASE STUDY 2

The parameters for this study were as follows:

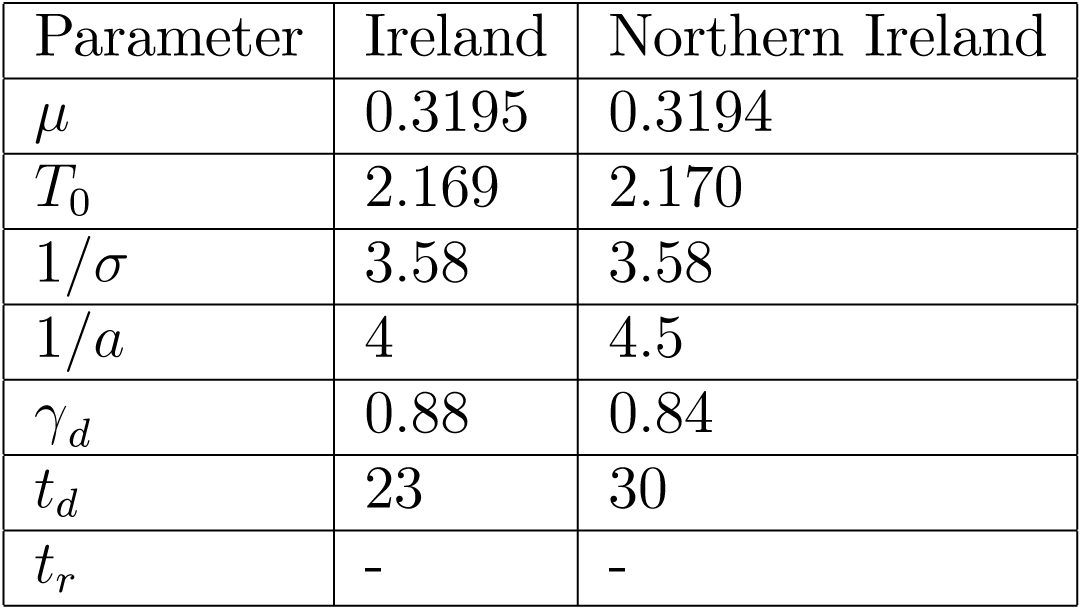

| Parameter | Ireland | Northern Ireland |
| --- | --- | --- |
| $\mu$ | 0.3195 | 0.3194 |
| $T_0$ | 2.169 | 2.170 |
| $1/\sigma$ | 3.58 | 3.58 |
| $1/a$ | 4 | 4.5 |
| $\gamma_d$ | 0.88 | 0.84 |
| $t_d$ | 23 | 30 |
| $t_r$ | - | - |

There was a single transmission/contact reduction intervention in each region. Note that the assumed initial doubling time for Northern Ireland is close to that for Ireland in this case. The derived parameters were as follows:

|  | Border Closed |  | Border Open |  |
| --- | --- | --- | --- | --- |
| Parameter | Ireland | Northern Ireland | Ireland | Northern Ireland |
| $\beta_0$ | 1.22095 | 1.16080 | 1.21885 | 1.15858 |
| $R_0$ | 4.88380 | 5.22359 | 4.89564 | 5.25795 |

The infective population and the effective reproduction ratio versus time are shown for both the open and closed border cases for Ireland in Figure 4A. The corresponding results for Northern Ireland are shown in Figure 4B. The differences between the open and closed border infective peak populations and their times of occurrence are small for both regions.

**FIGURE 4A.**
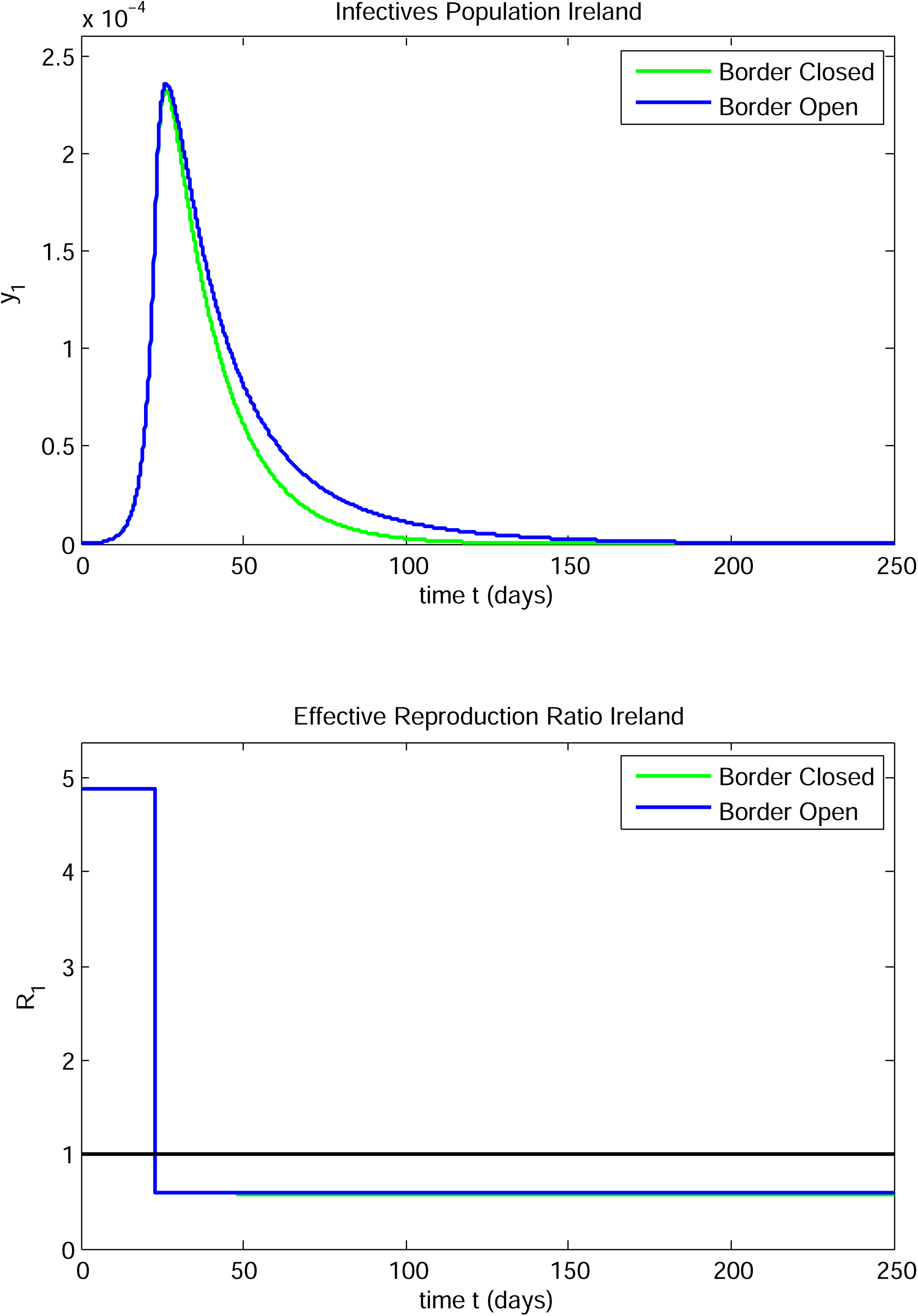

**FIGURE 4B.**
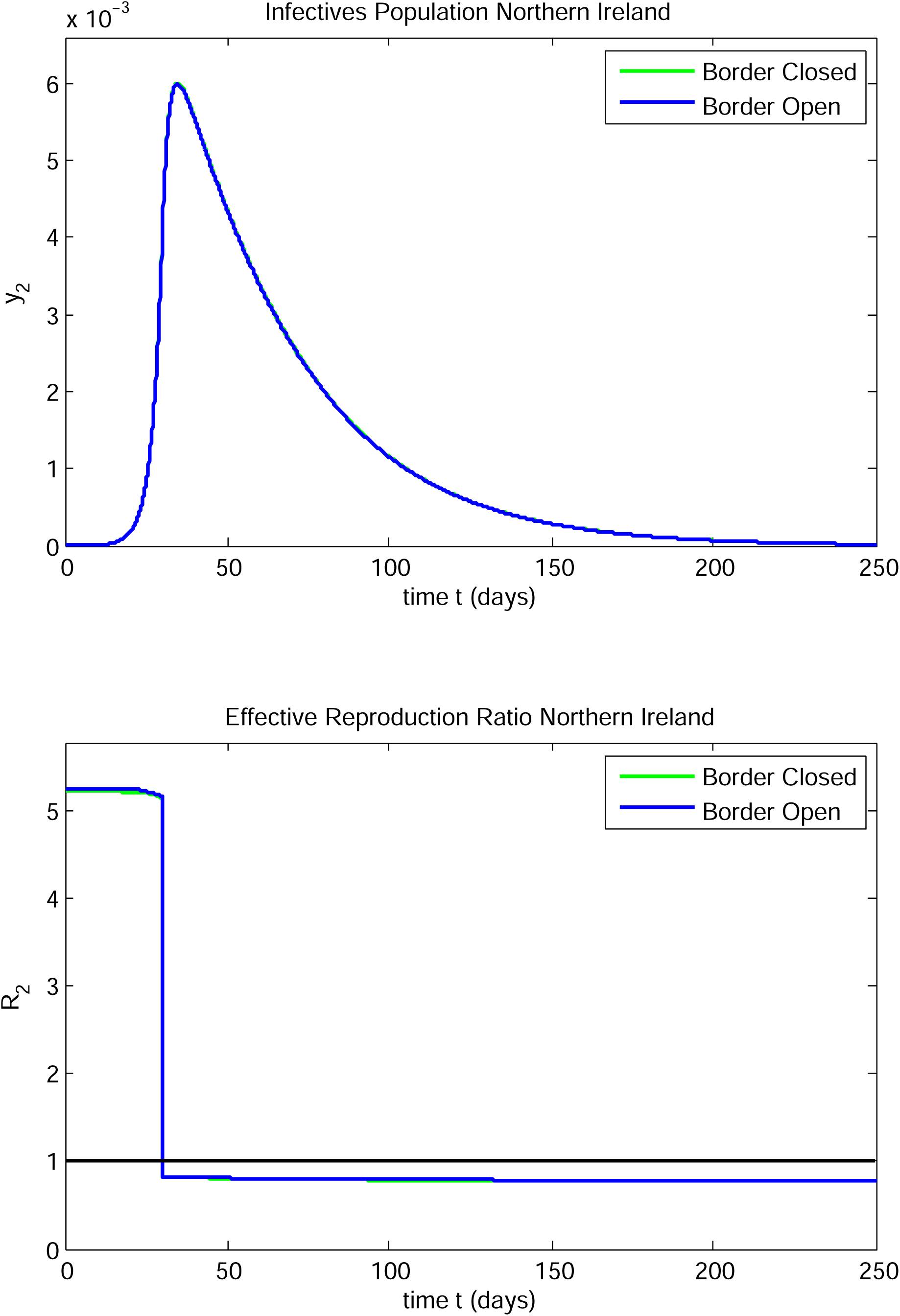

|  | Peak Infective Fraction |  |  | Peak Time |  |  |
| --- | --- | --- | --- | --- | --- | --- |
| Region | Closed | Open | % Diff | Closed | Open | Diff |
| Ireland | $2.33 \times 10^{-4}$ | $2.36 \times 10^{-4}$ | 1.3% | 26.2 | 26.4 | 0.2 |
| Northern Ireland | $6.00 \times 10^{-3}$ | $5.98 \times 10^{-3}$ | 0.3% | 34.8 | 34.8 | 0.0 |

The effective reproduction ratio for Ireland falls to about 0.6 and that for Northern Ireland falls to about 0.8, post-intervention. The fraction of infectives rises rapidly and decays relatively slowly in both regions. The main effect of the lower Northern Ireland initial infective doubling time is to dramatically increase the size of the peak in Northern Ireland relative to that in Case Study 1; it has a minor effect on the peak in Ireland.

#### CASE STUDY 3

The parameters for this study were as follows:

| Parameter | Ireland | Northern Ireland |
| --- | --- | --- |
| $\mu$ | 0.319512 | 0.319365 |
| $T_0$ | 2.16939 | 2.17039 |
| $1/\sigma$ | 3.58 | 3.58 |
| $1/a$ | 4 | 4.5 |
| $\gamma_d$ | 0.88 | 0.76 |
| $t_d$ | 23 | 30 |
| $t_r$ | - | - |

There was a single transmission/contact reduction intervention in each region. Note that the assumed initial doubling time for Northern Ireland is assumed to be close to that in Ireland in this case and that the value of *γ*_*d*2_ is smaller than that in case studies 1 and 2. The derived parameters were as follows:

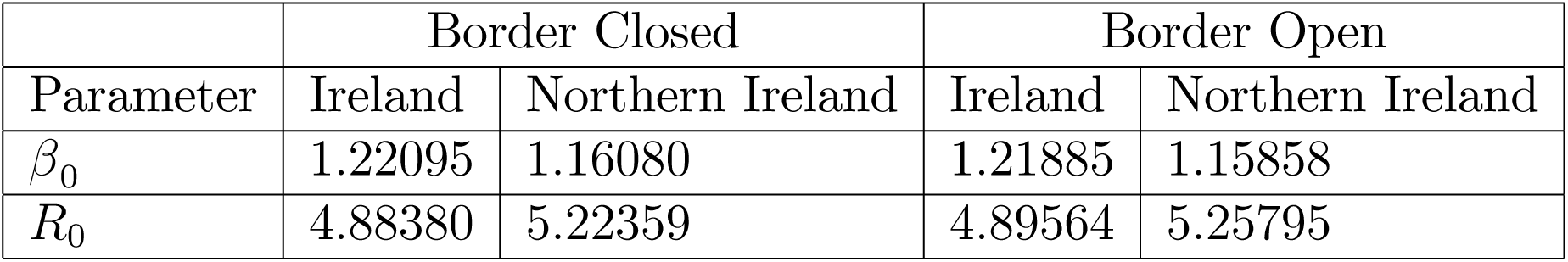

The infectives population and the effective reproduction ratio versus time are shown for both the open and closed border cases for Ireland in Figure 5A. The corresponding results for Northern Ireland are shown in Figure 5B. In this case, the presence of an open border causes a substantial effect in Ireland. In the closed border case, there is a single peak in Ireland and also in Northern Ireland. Leaving the border open causes a second peak in Ireland after the first peak. The data on the peaks, and the Ireland infectives local minimum in the case of the open border, are shown in the tables below.

|  | First Peak Infectives Fraction |  |  | Peak Time |  |  |
| --- | --- | --- | --- | --- | --- | --- |
| Region | Closed | Open | % Diff | Closed | Open | Diff |
| Ireland | $2.33 \times 10^{-4}$ | $2.36 \times 10^{-4}$ | 1% | 26.2 | 26.4 | 0.2 |
| Northern Ireland | $1.72 \times 10^{-2}$ | $1.71 \times 10^{-2}$ | 1% | 93.7 | 93.8 | 0.1 |

| Region | Minimum Infectives Fraction | Second Peak Infectives Fraction |
| --- | --- | --- |
| Ireland | $7.98 \times 10^{-5}$ | $9.58 \times 10^{-5}$ |
| Time | 67.6 | 108.1 |

**FIGURE 5A.**
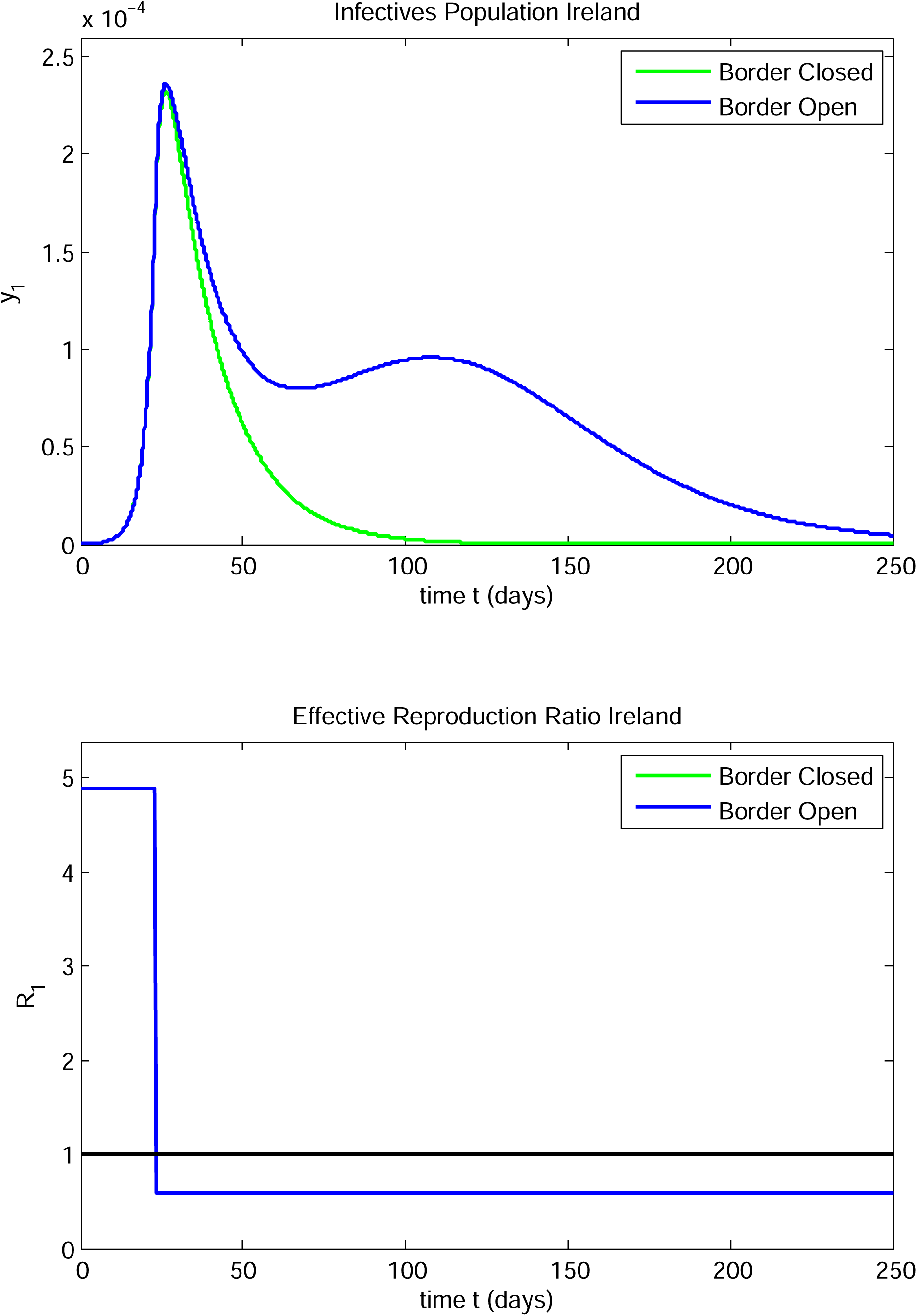

**FIGURE 5B.**
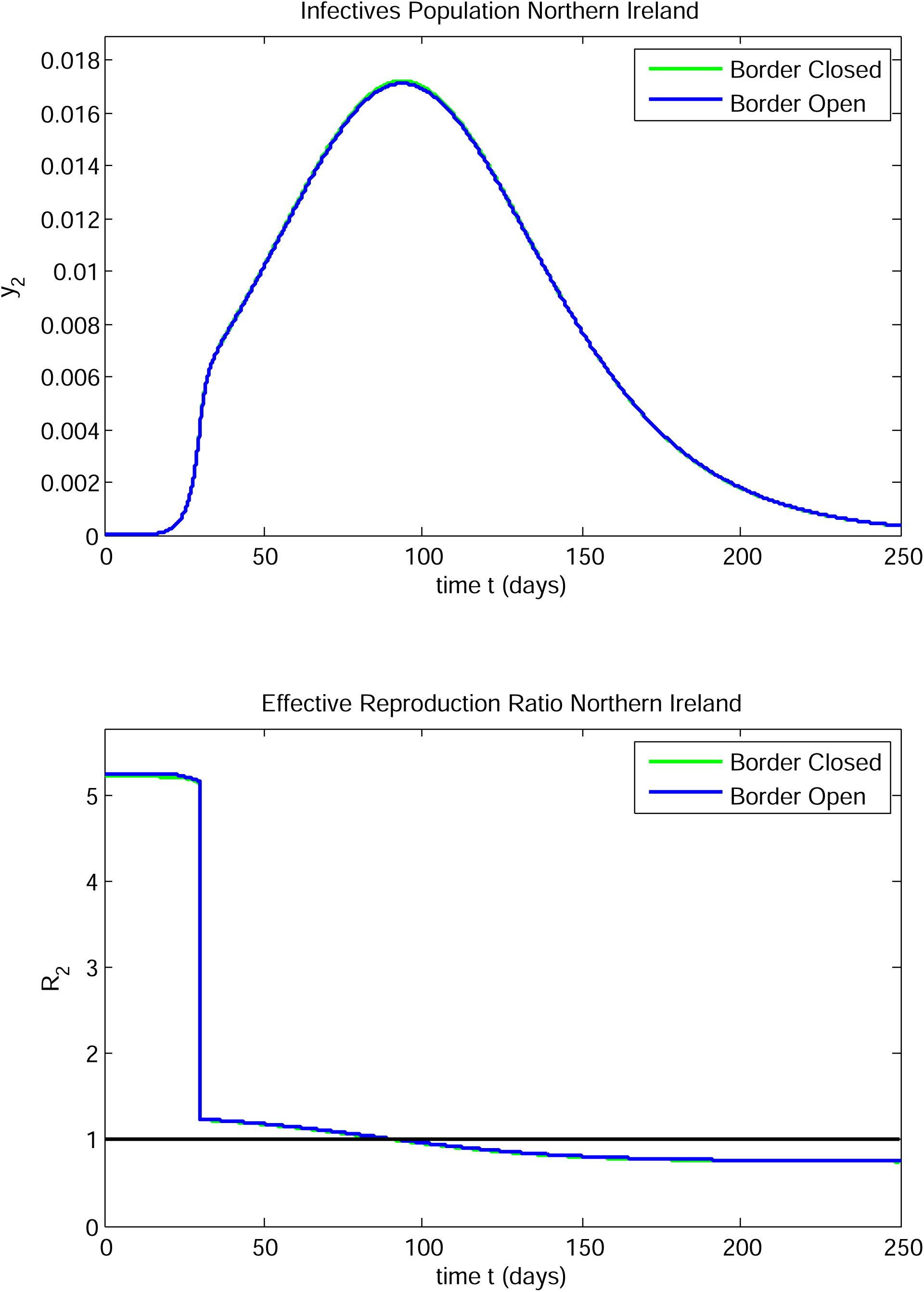

**FIGURE 5C.**
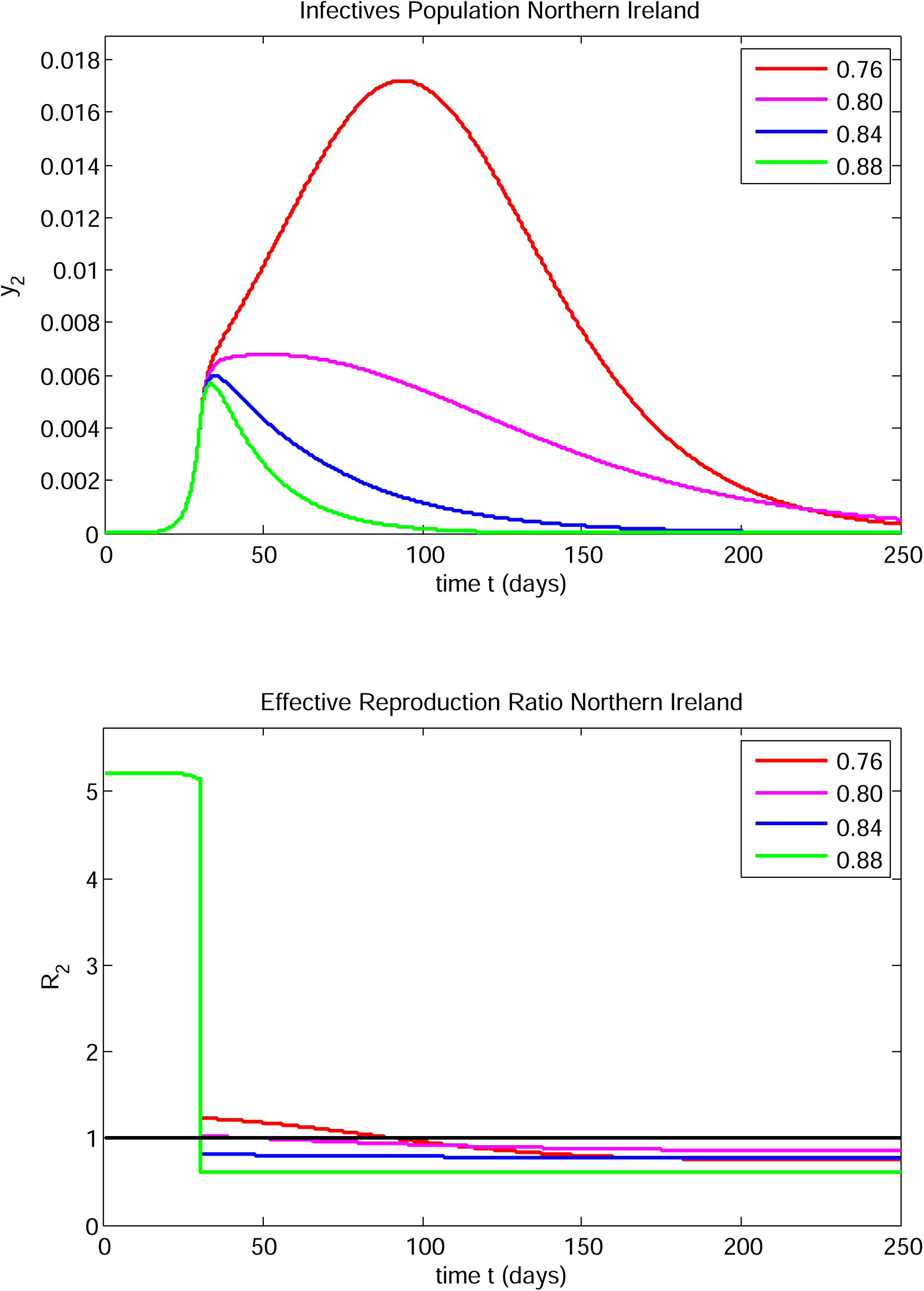

The infective fraction in Northern Ireland tends to a very high peak of about 1.71 10^−2^ at day 94 in both the open and closed border cases. A transmission/contact reduction would presumably be made by the Northern Ireland government before that peak was reached, but even with such mitigation, the delayed effect in Ireland could be substantial. *R*_2_ drops to a value of 1.24 immediately after the intervention on day 30 and then decays to unity at day 93 after which it decays further to 0.76 by day 250. In contrast, *R*_1_ drops to a value of 0.6 on day 23 and decays to 0.59 at day 250. The values of *R*_1_ and *R*_2_ are little affected by whether the border is open or closed. Note that *R*_*i*_*(t)* is the number of infectives created in both regions by a single infective in region *i* at time *t*. Even though *R*_1_ remains substantially below unity after day 23, nevertheless the infective fraction starts to increase after day 68. This might come as a surprise to Irish authorities if they were basing policy on a single region model for Ireland and if there was not an awareness of the interaction between the Ireland and the Northern Ireland epidemic progression. The second peak in Ireland is driven entirely by infectives in Northern Ireland. Note also that the progress of the epidemic in Northern Ireland, in this case, is essentially unaffected by whether the border is open or closed.

Another feature of this **SEIR** border interaction model arises in this case study. The very high infective peak in Northern Ireland is a consequence of one characteristic of the model which occurs when the effective reproduction ratio evolves such that its value is close to unity for an extended period, and in particular when it transitions from a period during which it is a little above unity to a period when it is a little below unity. This is illustrated in Figure 5C which shows four infectives curves corresponding to the data for this problem for Northern Ireland with the border closed, but with *γ*_*d2*_ varying from 0.88 down to 0.76 in steps of 0.4. Approximate sizes and locations of the peaks are shown in the table:

| $\gamma_{d2}$ | Peak Infective Fraction | Peak Time |
| --- | --- | --- |
| 0.92 | $5.5 \times 10^{-3}$ | 33 |
| 0.88 | $5.7 \times 10^{-3}$ | 34 |
| 0.84 | $6.0 \times 10^{-3}$ | 35 |
| 0.80 | $6.8 \times 10^{-3}$ | 51 |
| 0.76 | $1.7 \times 10^{-2}$ | 94 |
| 0.72 | $3.5 \times 10^{-2}$ | 87 |

If *R*_2_(t) gets closer to unity for an extended period, the infective peak first broadens dramatically and then increases in height substantially. This type of behaviour is not confined to variations in *γ*_*di*_, *i* = 1, 2 and can also occur consequent to even slight changes in 1/*a*_*i*_, *i* = 1, 2. In this study, a 11% decrease in *γ*_*d2*_ (from 0.84 to 0.76) leads to a 169% increase of peak time of occurrence from 35 days to 94 days. Larger delays consequent on much smaller changes in *γ*_*di*_ values can occur. These instabilities are discussed further in Case Study 10.

Note that this case study assumed *T*_02_ is close to 2. It is one of a number of cases included to enable the Irish government to err on the side of caution, as the initial doubling period in Northern Ireland was anomalous, as noted above.

This example is important because if the Irish Government was basing policy solely on an Ireland model not taking the border into account (effectively a closed border model), then the slower decrease from the peak might not be discerned and the upturn following day 68 would come as a surprise, and could not be explained by the model, since *R*_1_ would continue to remain stable at the value 0.6. If action were not taken, an extended period with the infective level at about 40% of the first peak level would ensue. This is an example where having an all-island model would be essential to understand what is happening. It would be important to investigate if this type of effect would occur in a model incorporating asymptomatic infection and testing/tracing.

#### CASE STUDY 4

The parameters for this study were as follows:

| Parameter | Ireland | Northern Ireland |
| --- | --- | --- |
| $\mu$ | 0.319512 | 0.227221 |
| $T_0$ | 2.16939 | 3.05054 |
| $1/\sigma$ | 3.58 | 3.58 |
| $1/a$ | 4 | 4.5 |
| $\gamma_d$ | 0.9 | 0.8 |
| $t_d$ | 26 | 30 |
| $t_r$ | - | - |

There was a single transmission/contact reduction intervention in each region. Note that the intervention on day 26 in Ireland is later than that in the previous cases. The time between interventions is 4 days in this case. The derived parameters are as follows:

|  | Border Closed |  | Border Open |  |
| --- | --- | --- | --- | --- |
| Parameter | Ireland | Northern Ireland | Ireland | Northern Ireland |
| $\beta_0$ | 1.22095 | 0.81504 | 1.21947 | 0.81283 |
| $R_0$ | 4.88380 | 3.66770 | 4.89313 | 3.69478 |

The infective population and the effective reproduction ratio versus time are shown for both the open and closed border cases for Ireland in Figure 6A.

**FIGURE 6A.**
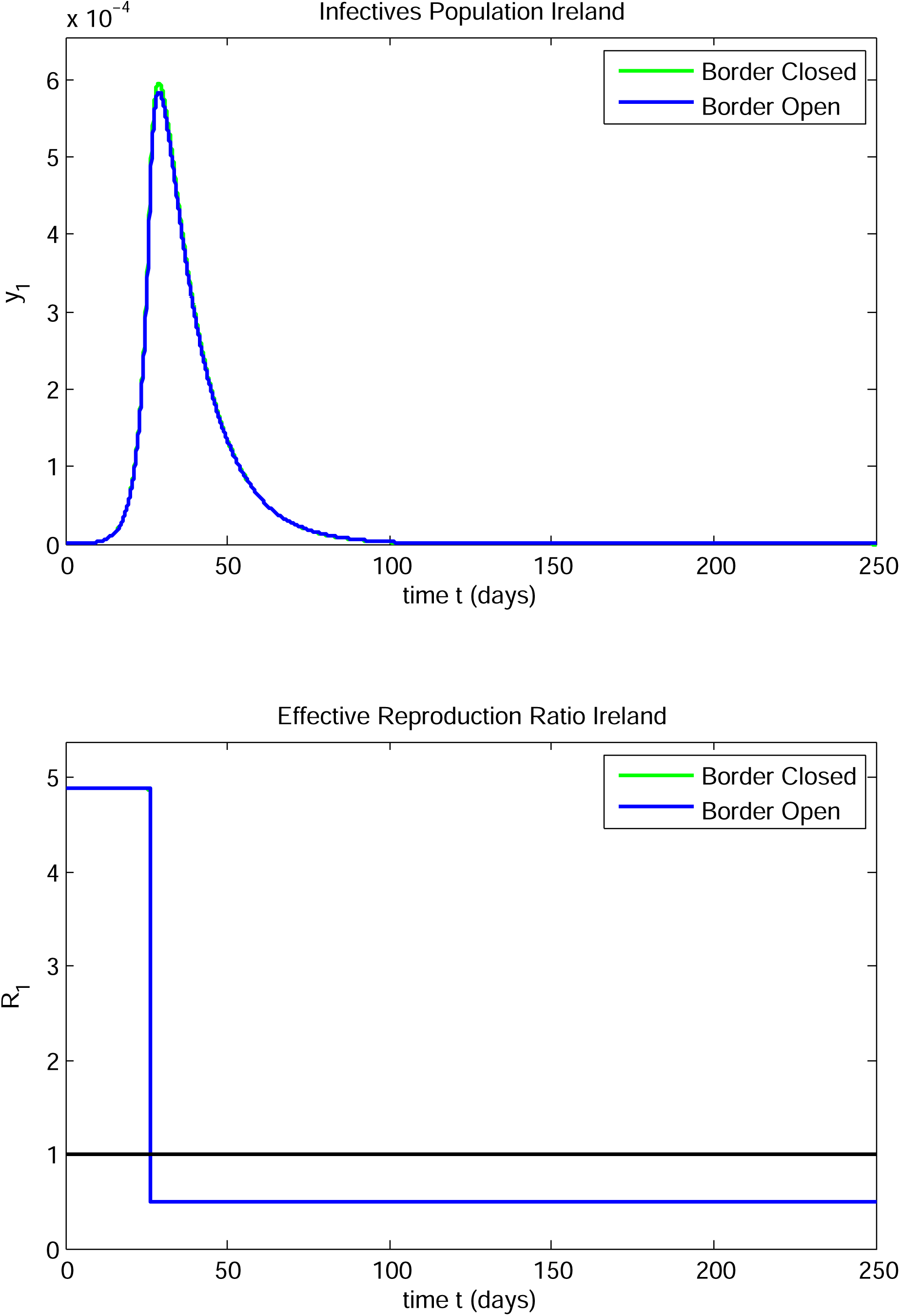

**FIGURE 6B.**
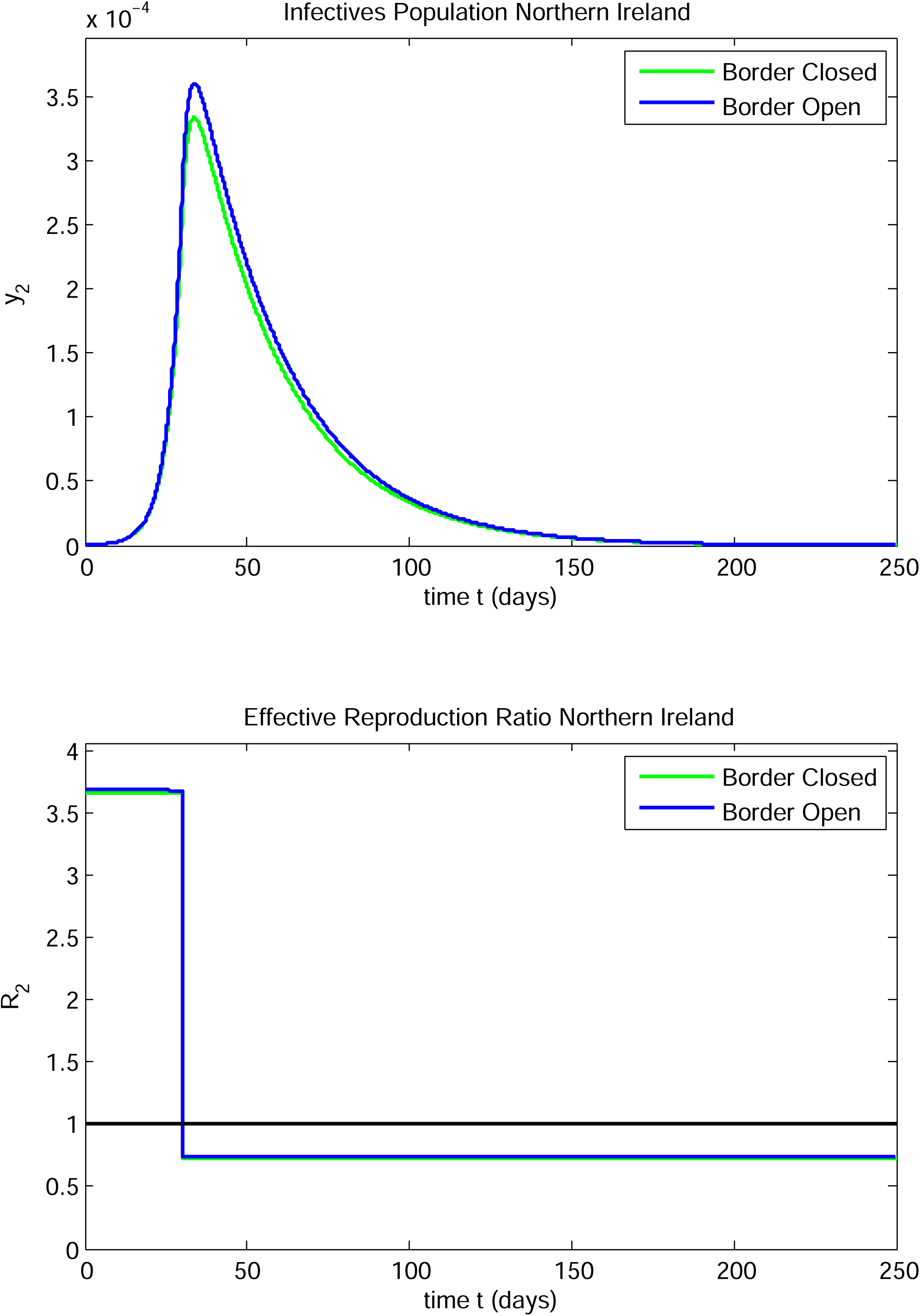

The corresponding results for Northern Ireland are shown in Figure 6B. The differences between the open and closed border infective peak populations and their times of occurrence are minor for Ireland, but not insignificant for Northern Ireland:

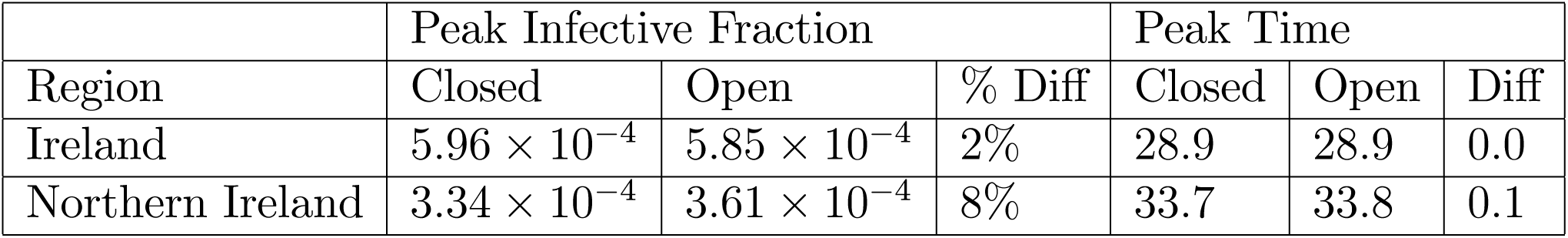

| Region | Peak Infective Fraction |  |  | Peak Time |  |  |
| --- | --- | --- | --- | --- | --- | --- |
|  | Closed | Open | % Diff | Closed | Open | Diff |
| Ireland | $5.96 \times 10^{-4}$ | $5.85 \times 10^{-4}$ | 2% | 28.9 | 28.9 | 0.0 |
| Northern Ireland | $3.34 \times 10^{-4}$ | $3.61 \times 10^{-4}$ | 8% | 33.7 | 33.8 | 0.1 |

#### CASE STUDY 5

The parameters for this study were as follows:

CASE STUDY 5 The parameters for this study were as follows:
| Parameter | Ireland | Northern Ireland |
| --- | --- | --- |
| $\mu$ | 0.319512 | 0.319365 |
| $T_0$ | 2.16939 | 2.17039 |
| $1/\sigma$ | 3.58 | 3.58 |
| $1/a$ | 4 | 4.5 |
| $\gamma_d$ | 0.88 | 0.84 |
| $\gamma_r$ | 0.88 | 0.7 |
| $t_d$ | 23 | 30 |
| $t_r$ | 90 | 90 |

There was a single transmission/contact reduction intervention in each region. There was also a transmission/contact increase corresponding to a lockdown release in Northern Ireland on day 90, whereas a low value of *β*_1_ was maintained in Ireland. The derived parameters were as follows:

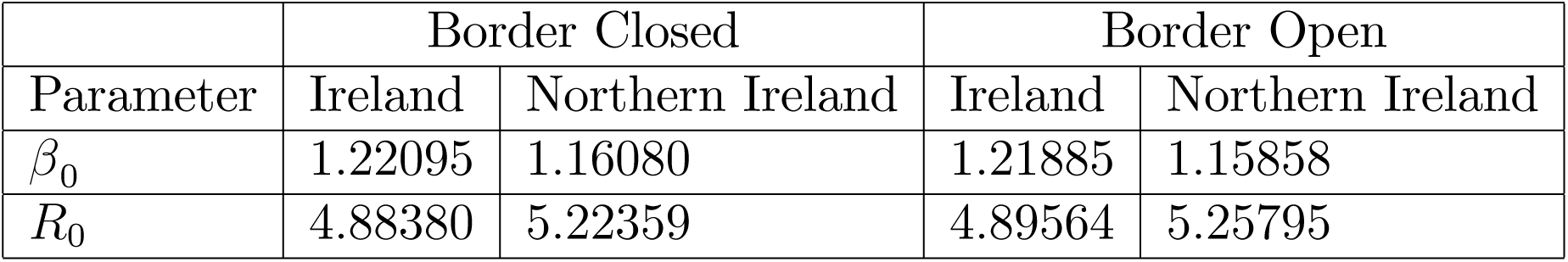

The infective fraction and the effective reproduction ratio versus time are shown for both the open and closed border cases for Ireland in Figure 7A. The corresponding results for Northern Ireland are shown in Figure 7B. In the closed border case, the infective fraction in Ireland rose to a maximum and decayed to zero. In the open border case, the infective fraction in Ireland rose to a peak, decayed to a local minimum and increased again to reach a second peak after which the fraction decayed to zero. In both the open and closed border cases, there were two infective peaks in Northern Ireland.

**FIGURE 7A.**
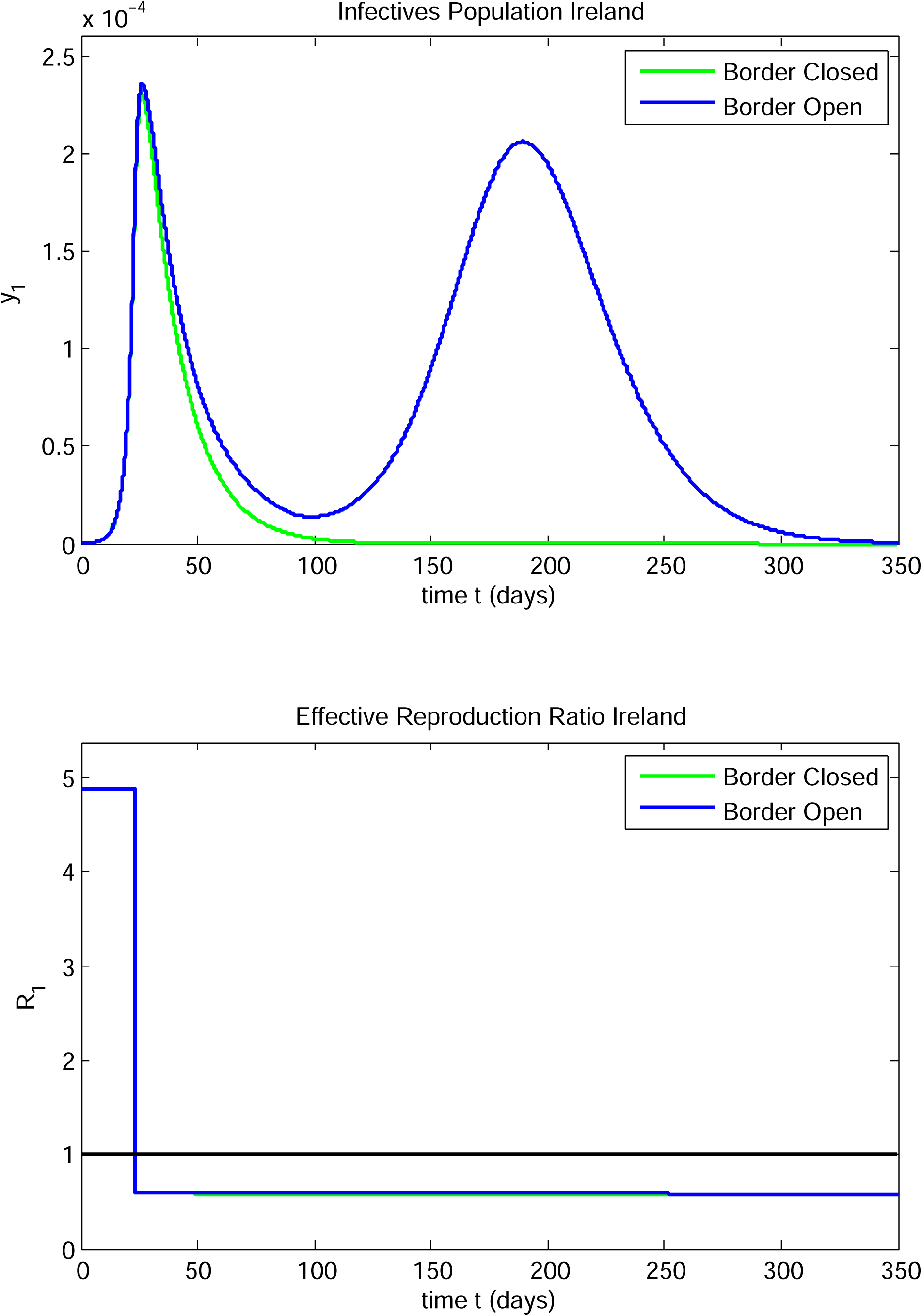

**FIGURE 7B.**
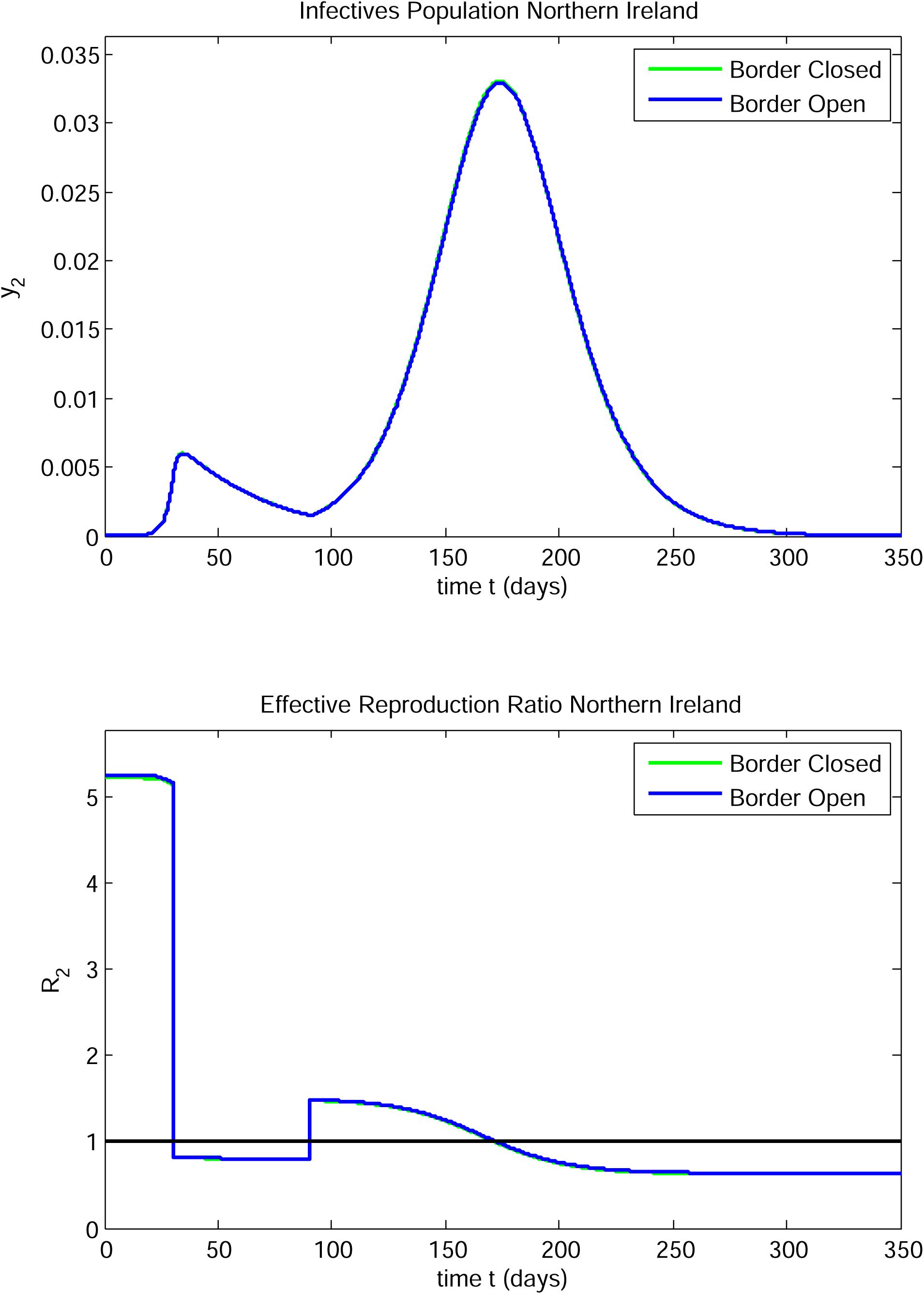

| Region | 1st Peak Infectives Fraction |  |  | 1st Peak Time |  |  |
| --- | --- | --- | --- | --- | --- | --- |
|  | Closed | Open | % Diff | Closed | Open | Diff |
| Ireland | $2.31 \times 10^{-4}$ | $2.36 \times 10^{-4}$ | 2% | 26.2 | 26.4 | 0.2 |
| Northern Ireland | $6.00 \times 10^{-3}$ | $5.98 \times 10^{-3}$ | 0.3% | 34.8 | 34.8 | 0.0 |

The sizes and times of occurrence of the first peak in both Northern Ireland and Ireland are both close. However, the border had a greater effect on the evolution in Ireland. Instead of a decay to zero after the infective peak at 26.4 days, the infective fraction appeared to be decaying to zero as in the closed border case, but on day 91 it began to increase again up to a new peak after which it decayed to zero, as shown in the tables below:

| Region | Minimum Infectives Fraction |  | Minimum Infectives time |  |
| --- | --- | --- | --- | --- |
|  | Closed | Open | Closed | Open |
| Ireland | - | $1.38 \times 10^{-5}$ | - | 99.3 |
| Northern Ireland | $1.51 \times 10^{-3}$ | $1.51 \times 10^{-3}$ | 90.8 | 90.8 |

| Region | 2nd Peak Infectives Fraction |  | 2nd Peak Time |  |
| --- | --- | --- | --- | --- |
|  | Closed | Open | Closed | Open |
| Ireland | - | $2.06 \times 10^{-4}$ | - | 189.5 |
| Northern Ireland | $3.30 \times 10^{-2}$ | $3.29 \times 10^{-2}$ | 174 | 174.2 |

During the period from day 30 to day 90, *R*_2_*(t)* remained close to 0.8. On day 90, *R*_2_*(t)* jumped to a value slightly under 1.5. It decreased thereafter, reaching the value 1 on day 173 and decreasing to slightly under 0.64 by day 350. During the entire period after day 23, the value of *R*_1_(t) remained very close to 0.6. If there was no awareness of the interaction taking place between Northern Ireland and Ireland, it could have reasonably been assumed that the epidemic was well-controlled in Ireland and the rise when the infective level had dropped to a low level at day 99 would not have been anticipated and would not be explainable using a single country model. So, it is important for planning for various contingencies in the post-peak period to be aware of possible impacts of Northern Ireland government interventions and societal responses on the progress of the epidemic in Ireland. This requires a two-region approach to modelling the epidemic.

#### CASE STUDY 6

The parameters for this study were as follows:

| Parameter | Ireland | Northern Ireland |
| --- | --- | --- |
| $\mu$ | 0.319512 | 0.227221 |
| $T_0$ | 2.16939 | 3.050540 |
| $1/\sigma$ | 3.58 | 3.58 |
| $1/a$ | 4 | 4.5 |
| $\gamma_d$ | 0.9 | 0.8 |
| $\gamma_r$ | 0.8 | 0.7 |
| $t_d$ | 26 | 30 |
| $t_r$ | 90 | 90 |

There was a single transmission/contact reduction in each region, followed by a single transmission/contact increase in each region corresponding to lockdown releases. The reductions were separated by 4 days, but the increases took place on the same day.

The derived parameters were as follows:

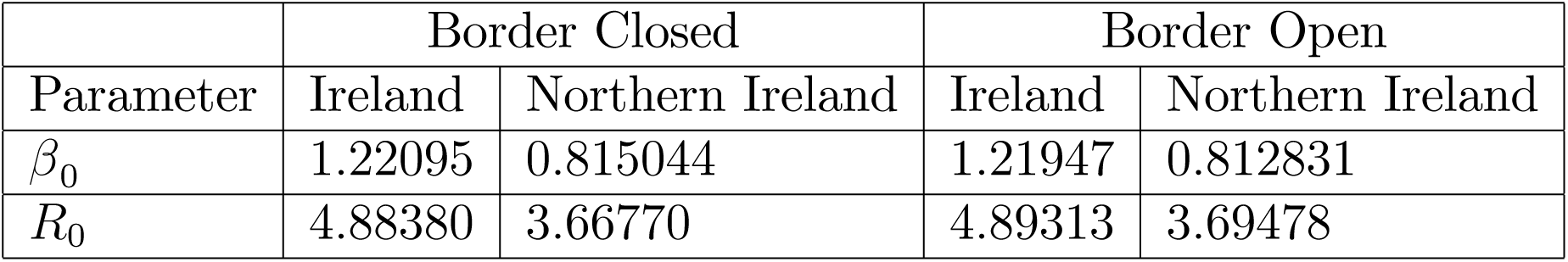

The infective population and the effective reproduction ratio versus time are shown for both the open and closed border cases for Ireland in Figure 8A. The corresponding results for Northern Ireland are shown in Figure 8B. In the closed border case, the infective fraction in Ireland rose to a maximum and decayed to zero. In the open border case, the infective fraction in Ireland rose to a peak, decayed to a local minimum and increased again to reach a second peak after which the fraction decayed to zero. The second peak was lower and very wide in comparison with the first. For both the open and closed border cases, the infective fraction in Northern Ireland rose to a peak, decayed to a local minimum and increased again to reach a second higher peak after which the fraction decayed to zero. The values and times of the extrema are as follows:

**FIGURE 8A.**
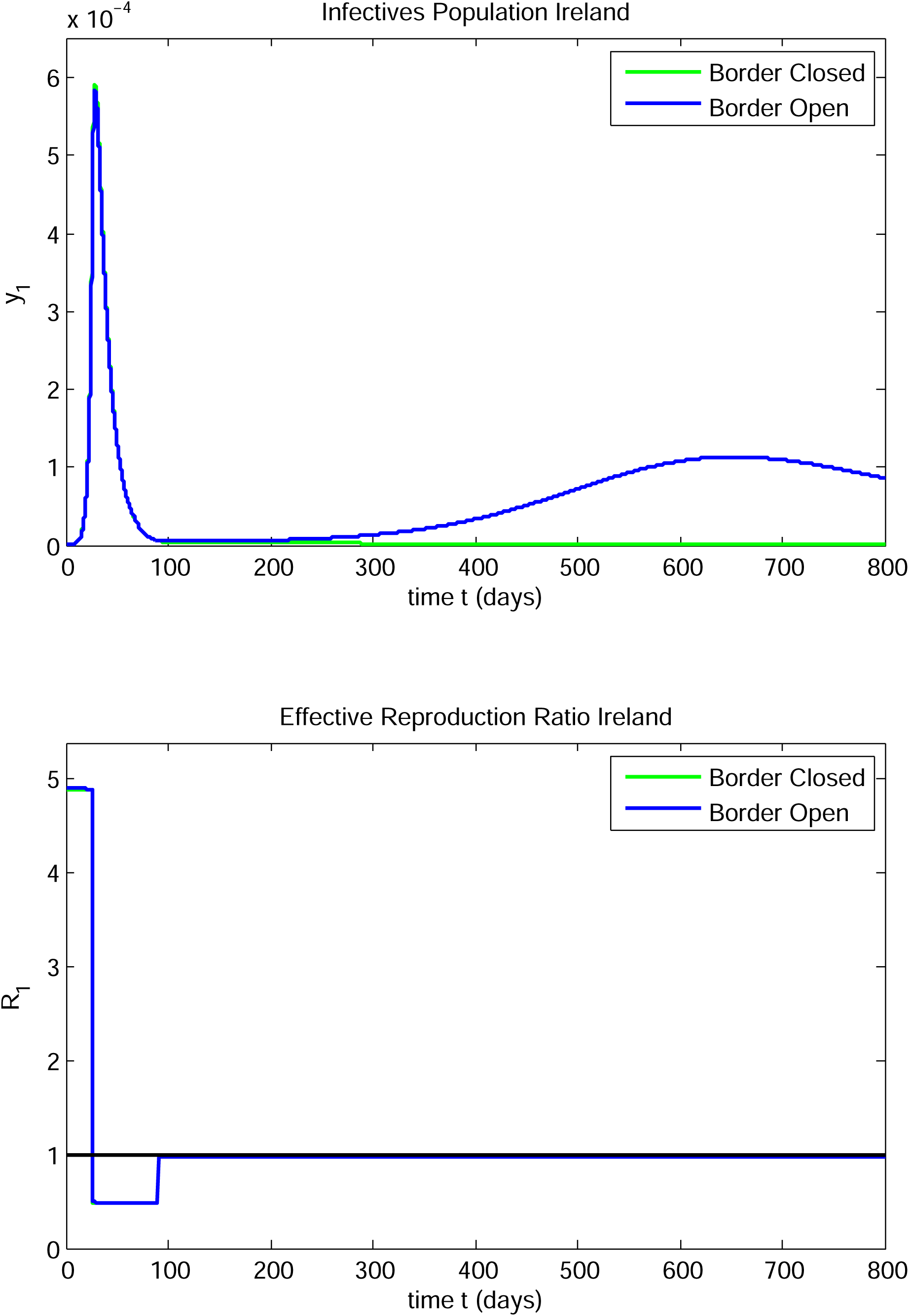

**FIGURE 8B.**
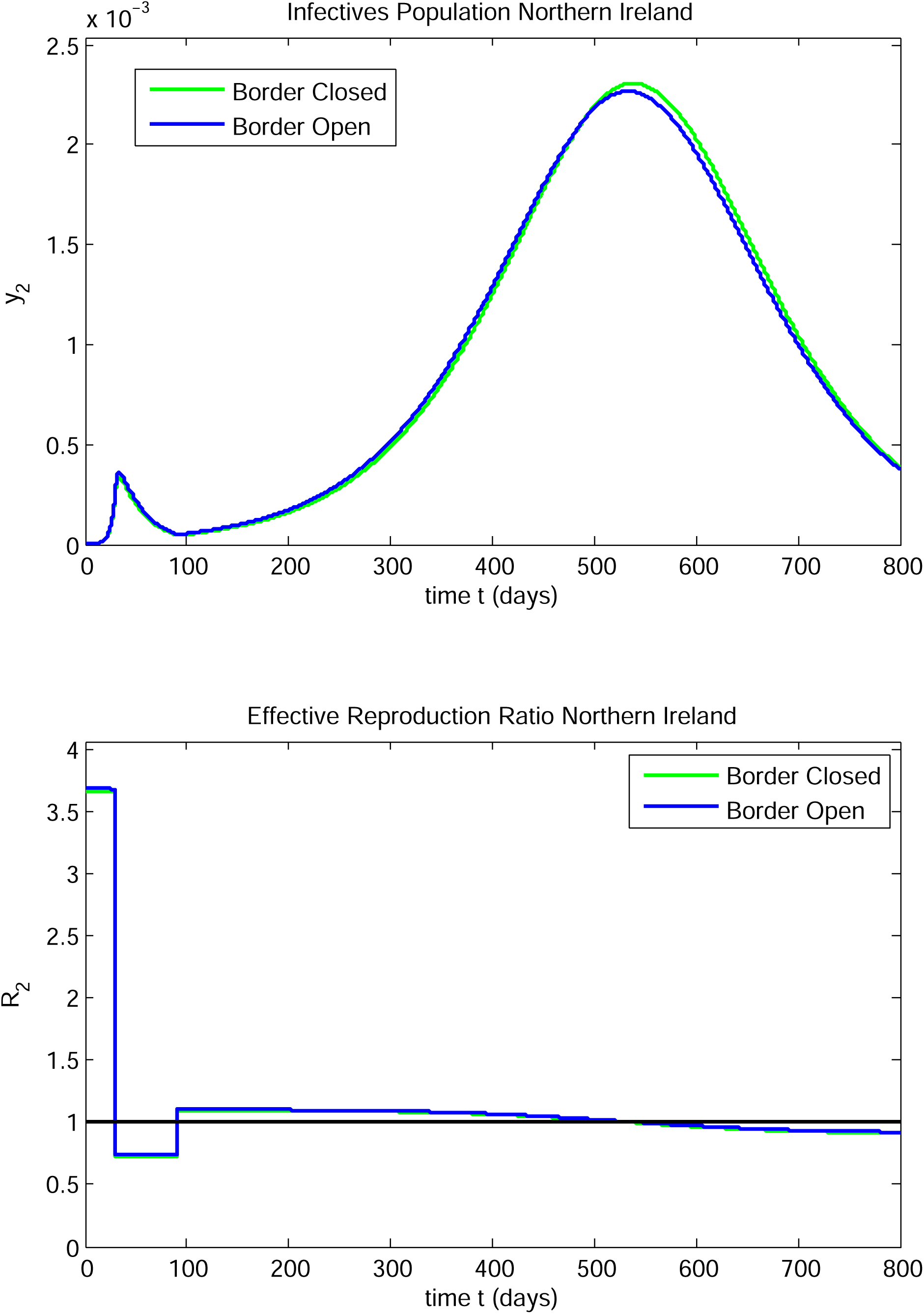

|  | First Peak Infective Fraction |  |  | First Peak Time |  |  |
| --- | --- | --- | --- | --- | --- | --- |
| Region | Closed | Open | % Diff | Closed | Open | Diff |
| Ireland | $5.92 \times 10^{-4}$ | $5.85 \times 10^{-4}$ | 1% | 28.9 | 28.9 | 0.0 |
| Northern Ireland | $3.34 \times 10^{-4}$ | $3.61 \times 10^{-4}$ | 8% | 33.7 | 33.8 | 0.1 |

| Region | Minimum Infective Fraction |  |  | Minimum Time |  |  |
| --- | --- | --- | --- | --- | --- | --- |
|  | Closed | Open | % Diff | Closed | Open | Diff |
| Ireland | - | $4.95 \times 10^{-6}$ | - | - | 101.9 | - |
| Northern Ireland | $4.67 \times 10^{-5}$ | $5.08 \times 10^{-5}$ | 9% | 92.8 | 92.8 | 0.0 |

| Region | 2nd Peak Infective Fraction |  |  | 2nd Peak Time |  |  |
| --- | --- | --- | --- | --- | --- | --- |
|  | Closed | Open | % Diff | Closed | Open | Diff |
| Ireland | - | $1.13 \times 10^{-4}$ | - | - | 653.3 | - |
| Northern Ireland | $2.31 \times 10^{-3}$ | $2.27 \times 10^{-3}$ | 2% | 538.7 | 533.8 | |

The early infective peak in Ireland was little affected by the border status, but the first peak in Northern Ireland was substantially increased (by 8%) in the case of the open border. After the infective fraction in Ireland decays to the low level of about 5 ×10^−6^, which is about 1% of the first peak level, it persists at this level for a long time. The second peak in Ireland is much lower and broader than the first.

Both *R*_1_*(t)* and *R*_2_*(t)* are little affected by the border status. The value of *R*_1_*(t)* drops to 0.5 on day 26, decreases to 0.49 on day 90 at which time it jumps to 0.98 and decays to 0.97 by day 800. There might be concern in Ireland about the value of *R*_1_ remaining slightly below 1 for an extended period, but the increase in the infective fraction while *R*_1_ remains below unity would be unexpected in the context of a one-region model for Ireland. The increase in infectives in Ireland is very gradual and is driven solely by the evolution in Northern Ireland. So, in this case, different policies and societal responses on both sides of the open border would have negative effects, of different natures, in both regions. If there was not awareness that Northern Ireland was affecting the evolution of the epidemic in Ireland, the occurrence of the second peak would be unexpected. Equally well, the open border causes an increase of 8% in the first peak in Northern Ireland.

The value of *R*_2_ drops to 0.74 on day 30 and remains very close to this level until day 90 at which time it increases to 1.1. It then decreases very slowly to 1 on day 544 and decreases further to 0.92 by day 800. The extended trough and subsequent peak in region 1 may be a feature associated with *R*_2_ staying close to unity for so long, as such a pronounced feature does not appear in Case Study 3 above.

#### CASE STUDY 7

This study differs from Case Study 6 only in the value of *T*_02_ which is close to *T*_01_ to examine the implications of the initial infective doubling times to be near 2 days - again, to err on the side of caution from the perspective of the Government in Ireland. The parameters for this study were as follows:

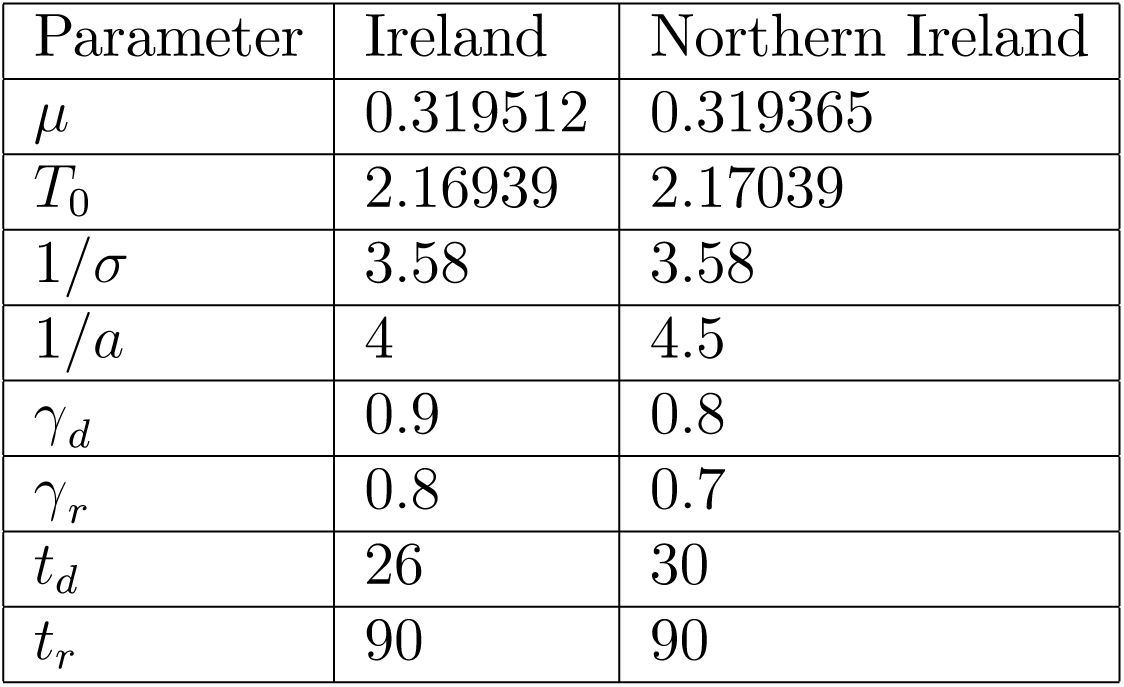

There was a single transmission/contact reduction in each region, followed by a single transmission/contact increase, corresponding to a lockdown release, in each region.

The derived parameters were as follows:

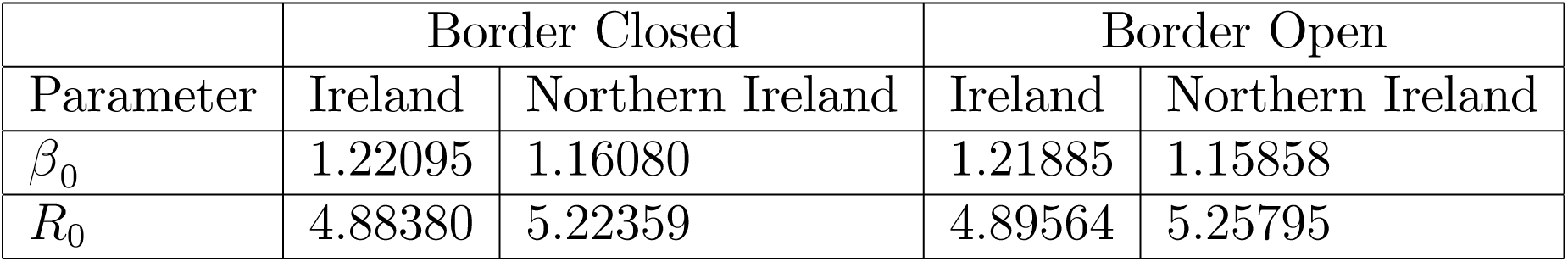

The infectives population and the effective reproduction ratio versus time are shown for both the open and closed border cases for Ireland in Figure 9A. The corresponding results for Northern Ireland are shown in Figure 9B. In the closed border case, the infective fraction in Ireland rose to a maximum and then decayed to zero. In the open border case, the infective fraction in Ireland rose to a peak, decayed to a local minimum, increased again to reach a second peak and then decayed to zero. The second Ireland peak was higher and dramatically wider than the first. Thus, the border status had a dramatic effect on the evolution of the infectives fraction in Ireland. Note, however, that the border status had little effect on the evolution of the effective reproduction ratio in Ireland. The border status had a negligible effect on the evolution of both the infective fraction and the effective reproduction ratio in Northern Ireland. The extrema in both regions are indicated in the following tables:

**FIGURE 9A.**
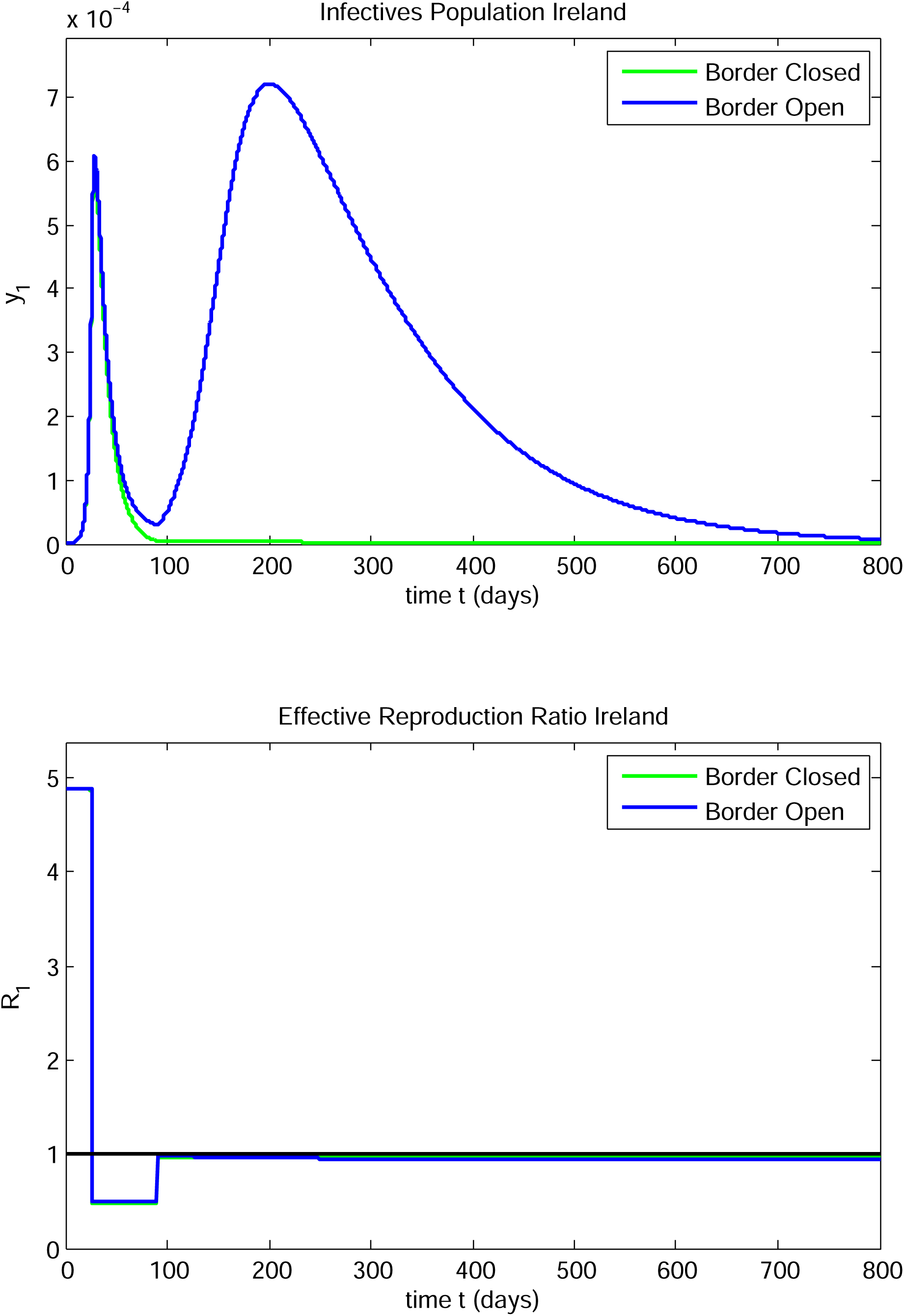

**FIGURE 9B.**
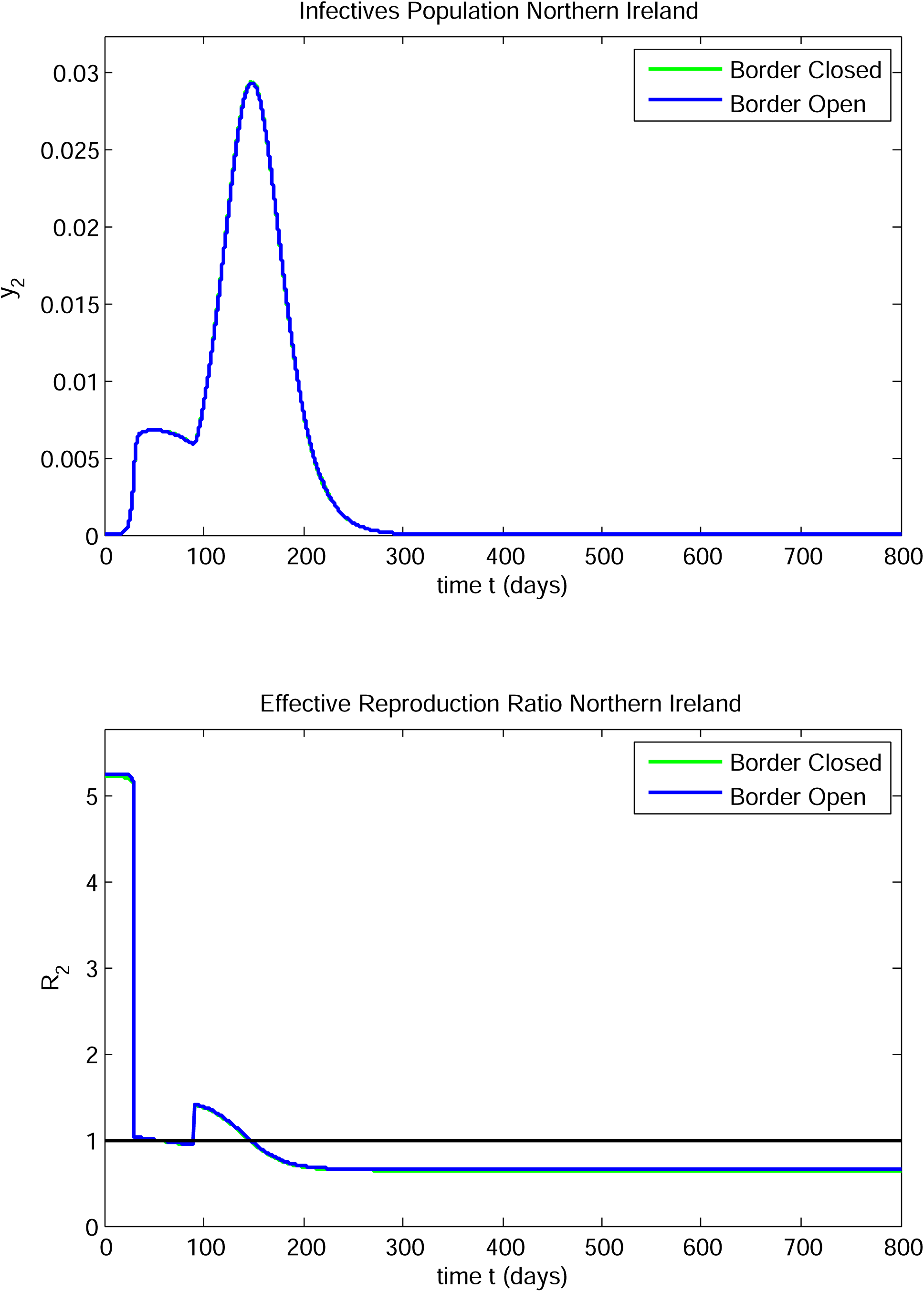

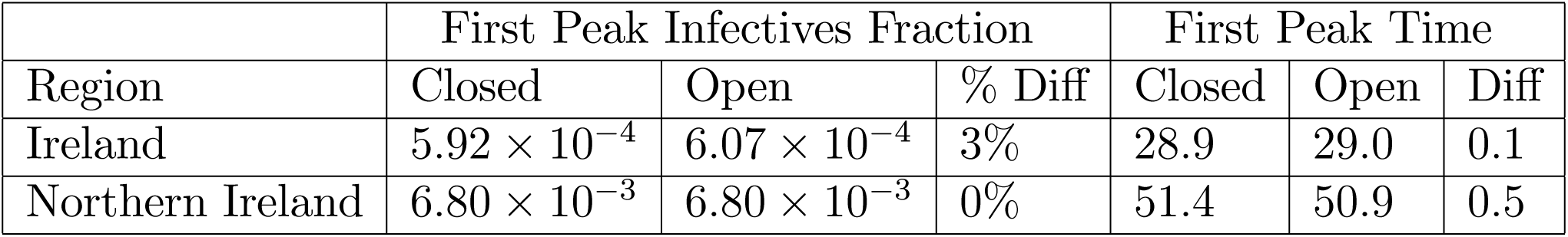

|  | Minimum Infectives Fraction |  |  | Minimum Time |  |  |
| --- | --- | --- | --- | --- | --- | --- |
| Region | Closed | Open | % Diff | Closed | Open | Diff |
| Ireland | - | $3.04 \times 10^{-5}$ | - | - | 90.4 | - |
| Northern Ireland | $5.90 \times 10^{-3}$ | $5.87 \times 10^{-3}$ | 0.5% | 90.3 | 90.3 | 0.0 |

|  | Second Peak Infectives Fraction |  |  | Second Peak Time |  |  |
| --- | --- | --- | --- | --- | --- | --- |
| Region | Closed | Open | % Diff | Closed | Open | Diff |
| Ireland | - | $7.21 \times 10^{-4}$ | - | - | 200.9 | - |
| Northern Ireland | $2.94 \times 10^{-2}$ | $2.94 \times 10^{-2}$ | 0% | 148.5 | 148.6 | 0.1 |

The early infective peak in Ireland was increased by 3% in the case of the open border. The second peak in Ireland, in the open border case is 22% higher than the single closed border first peak.

The value of *R*_*i*_ varied monotonically between interventions in region *i*, for *i* = 1, 2. The values just prior to and just after the interventions on days 26 and 90 in Ireland, for each border status, are as follows:

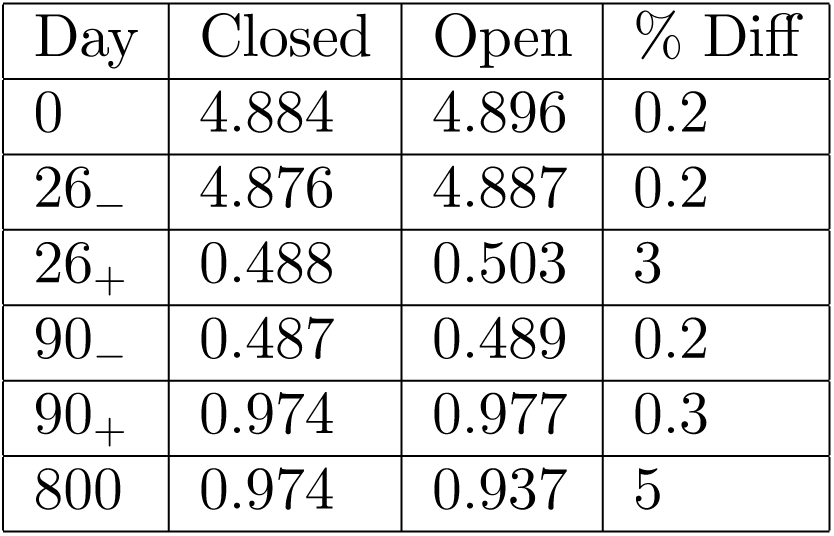

The corresponding data for Northern Ireland were as follows:

| Day | Closed | Open | % Diff |
| --- | --- | --- | --- |
| 0 | 5.224 | 5.257 | 0.6 |
| 30<sub>-</sub> | 5.147 | 5.164 | 0.3 |
| 30<sub>+</sub> | 1.029 | 1.034 | 0.5 |
| 90<sub>-</sub> | 0.941 | 0.946 | 0.5 |
| 90<sub>+</sub> | 1.411 | 1.419 | 0.6 |
| 800 | 0.646 | 0.655 | 1.4 |

After day 26, *R*_1_ remained below 1. So, on the basis of a single region model for Ireland, the infectives fraction should have continued to decrease, albeit slowly, after day 26. The subsequent rise to a broad and even higher peak would be unexpected and unexplained in the context of a one-region model.

The only difference in the data between this case and Case Study 6 is the lower value of *T*_02_, the initial doubling time for Northern Ireland (2.17 versus 3.05). This causes a substantial difference. The second infective peak in Ireland comes earlier and is much higher than in Case Study 6. It is also much broader than the first peak in this case. Again, the infectives level in Ireland has dropped to a relatively low value in Ireland when it starts to pick up to enter a new rapidly rising phase quite at odds with the stable *R*_1_ level, just below 1 in Ireland.

#### CASE STUDY 8

In this case, we look at the effect of increasing the border interaction parameters *α*_*i*_, *i* = 1, 2 on the evolution of the infectives and the effective reproduction ratios in both regions. Note that only the products *α*_*i*_*α*_*j*_, *i, j* = 1, 2 appear in the governing differential equations. We increase these products by an order of magnitude by increasing *α*_*i*_ by a factor of 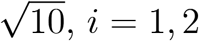.

We consider the problem described in Case Study 4, with the increased border interaction parameters. The data are as follows:

| Parameter | Ireland | Northern Ireland |
| --- | --- | --- |
| $\mu$ | 0.319512 | 0.227222 |
| $T_0$ | 2.16939 | 3.050534 |
| $1/\sigma$ | 3.58 | 3.58 |
| $1/a$ | 4 | 4.5 |
| $\gamma_d$ | 0.9 | 0.8 |
| $t_d$ | 26 | 30 |
| $t_r$ | - | - |
| $\alpha$ | $\frac{13}{490}\sqrt{10}$ | $\frac{13}{190}\sqrt{10}$ |

There was a single transmission/contact reduction intervention in each region. Note that the intervention was on day 26 in Ireland. The time between interventions is 4 days. The derived parameters for the two-region models for each set of (*α*_1_, *α*_2_) values were as follows:

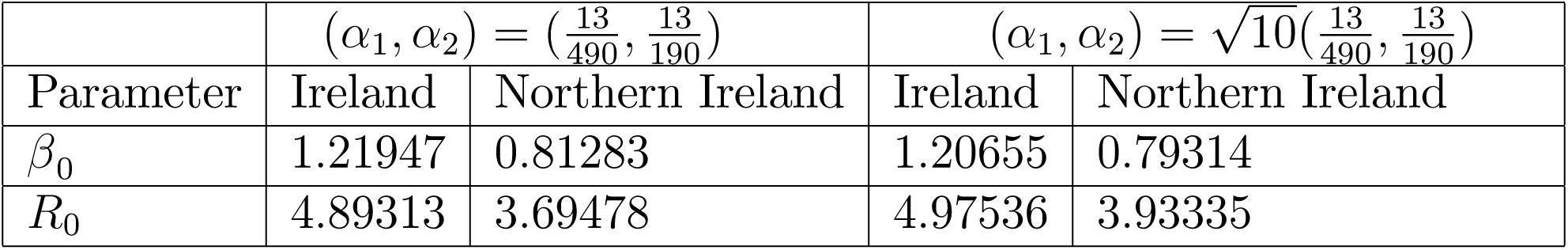

The infective population and the effective reproduction ratio versus time are shown for the low and high (*α*_1_, *α*_2_) cases for Ireland in Figure 10A. The corresponding results for Northern Ireland are shown in Figure 10B. The peak infective fractions for the two sets of (*α*_1_, *α*_2_) values for the two-region models are as follows:

**FIGURE 10A.**
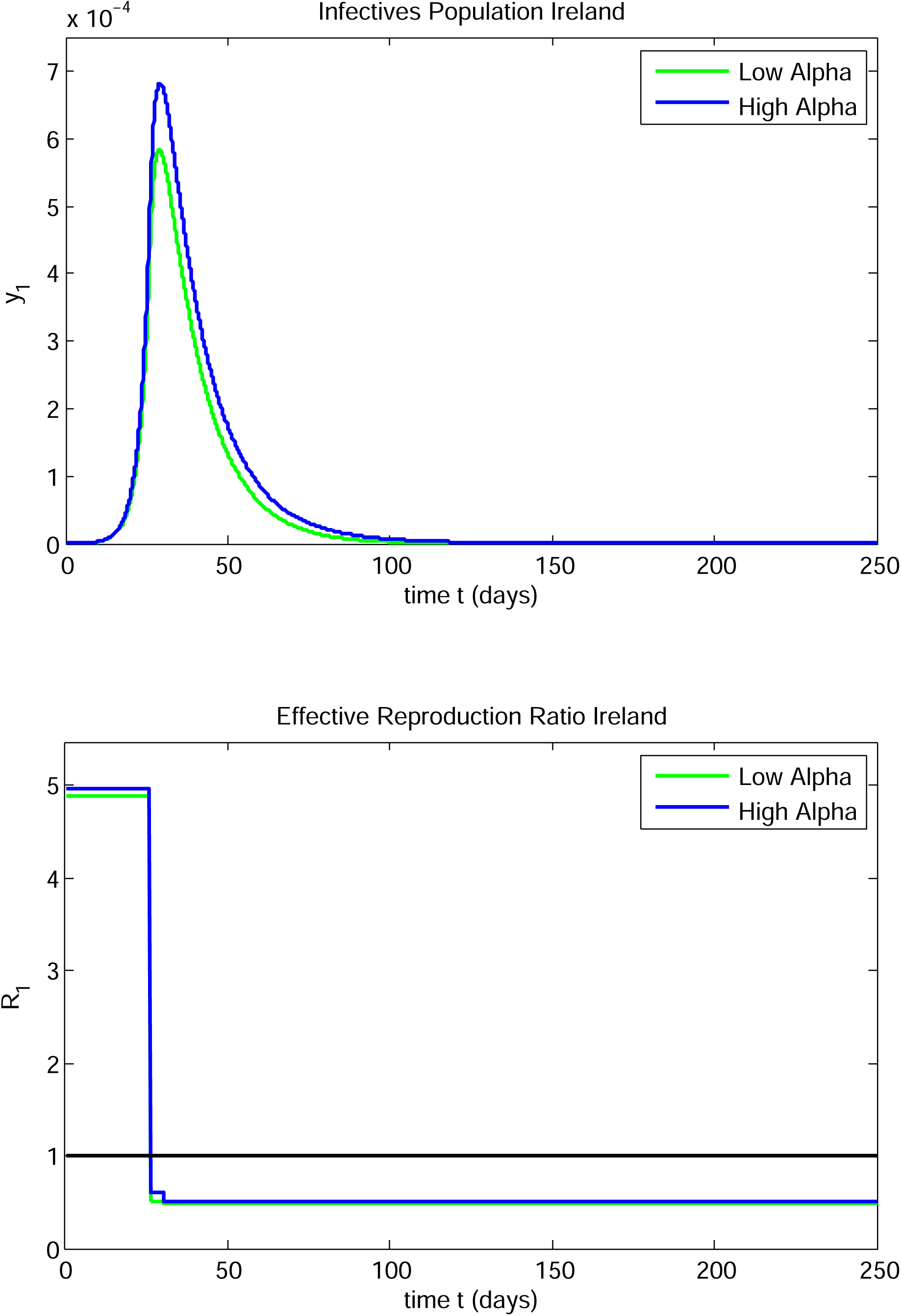

**FIGURE 10B.**
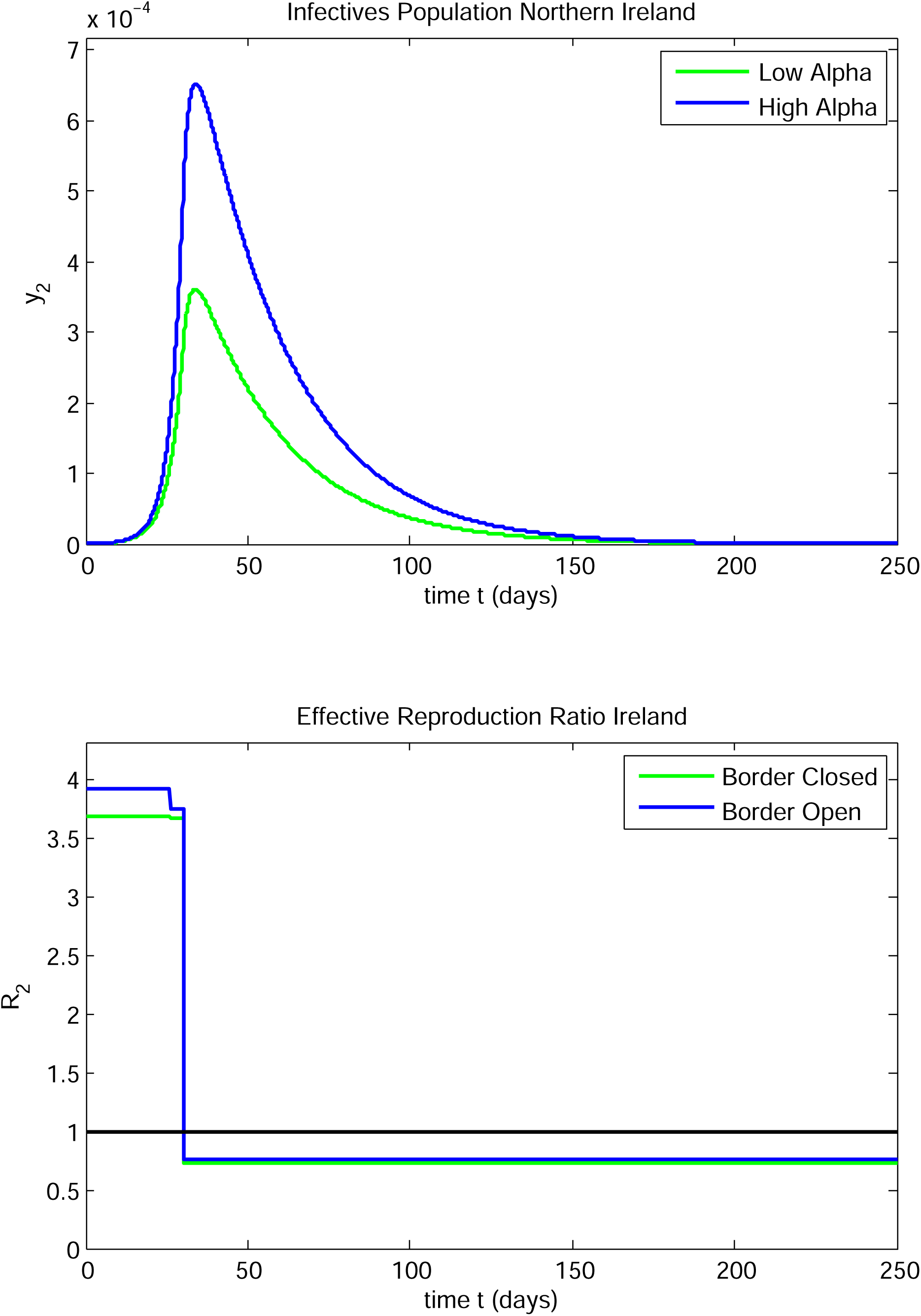

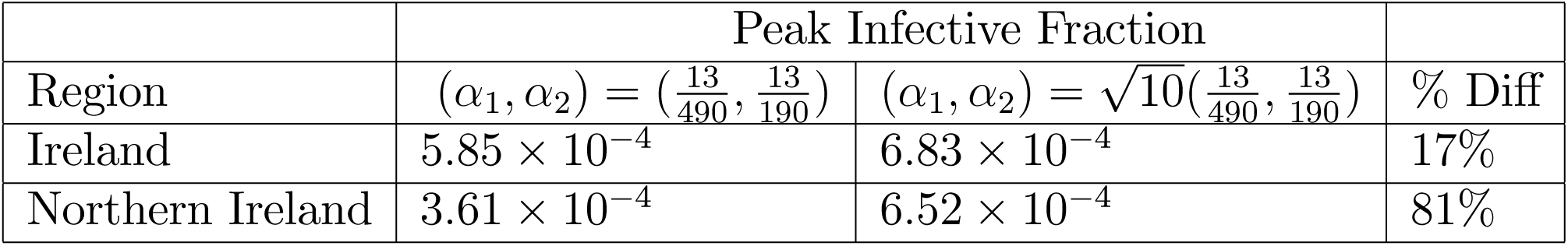

and the corresponding peak times are as follows:

|  | Peak Time |  |  |
| --- | --- | --- | --- |
| Region | $(\alpha_1, \alpha_2) = (\frac{13}{490}, \frac{13}{190})$ | $(\alpha_1, \alpha_2) = \sqrt{10}(\frac{13}{490}, \frac{13}{190})$ | Diff |
| Ireland | 28.9 | 29.1 | 0.3 |
| Northern Ireland | 33.8 | 34.0 | 0.1 |

For this problem, both peaks are sensitive to the increase in the border interaction parameters; the impact of this increased border interaction on Northern Ireland is very large. These results suggest a more thorough study of the three types of border interactions noted above to obtain more precise values for the interaction parameters would be worthwhile and of especial interest to the Northern Ireland Executive.

#### CASE STUDY 9

In this case, we look at the effect of increasing the border interaction parameters *α*_*i*_, *i* = 1, 2 on the evolution of the infectives and the effective reproduction ratios in both regions for the problem described in Case Study 6. We increase the products *α*_*i*_*α*_*j*_, *i, j* = 1, 2 by an order of magnitude by increasing *α*_*i*_ by a factor of 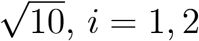.

The data are as follows:

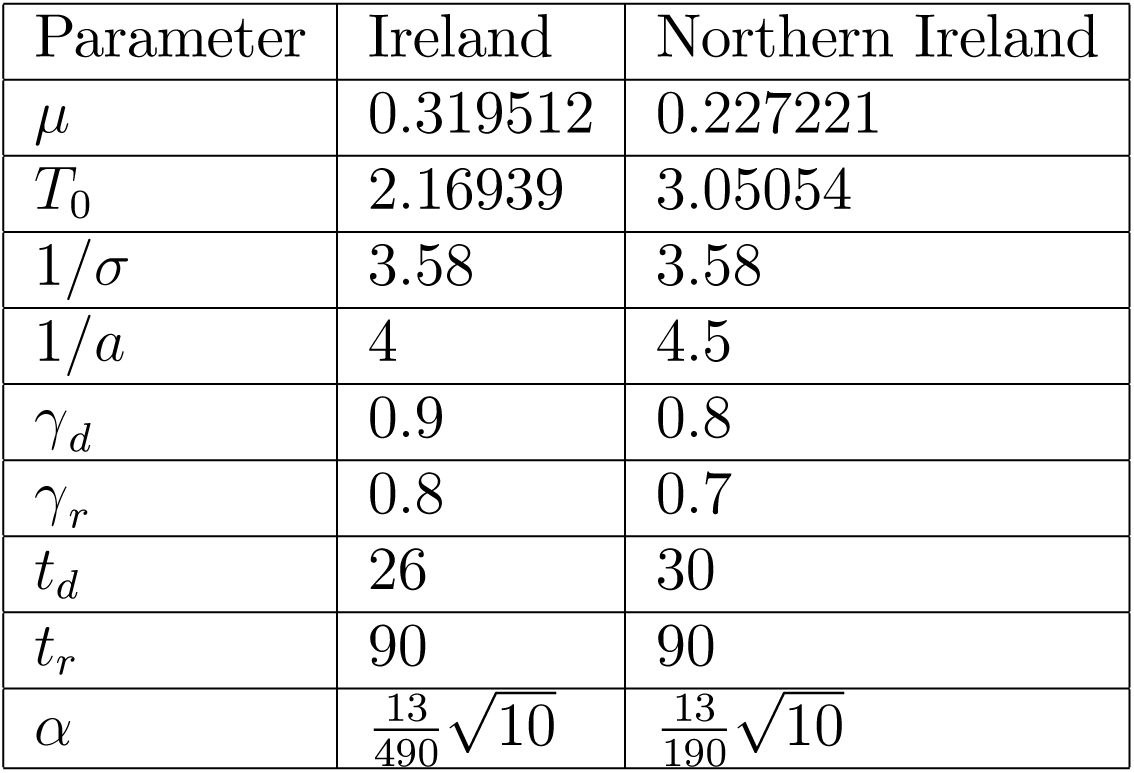

| Parameter | Ireland | Northern Ireland |
| --- | --- | --- |
| $\mu$ | 0.319512 | 0.227221 |
| $T_0$ | 2.16939 | 3.05054 |
| $1/\sigma$ | 3.58 | 3.58 |
| $1/a$ | 4 | 4.5 |
| $\gamma_d$ | 0.9 | 0.8 |
| $\gamma_r$ | 0.8 | 0.7 |
| $t_d$ | 26 | 30 |
| $t_r$ | 90 | 90 |
| $\alpha$ | $\frac{13}{490}\sqrt{10}$ | $\frac{13}{190}\sqrt{10}$ |

There was a single transmission/contact reduction in each region, followed by a single transmission/contact increase, corresponding to a lockdown release, in each region. The derived parameters for the two-region models in each case were as follows:

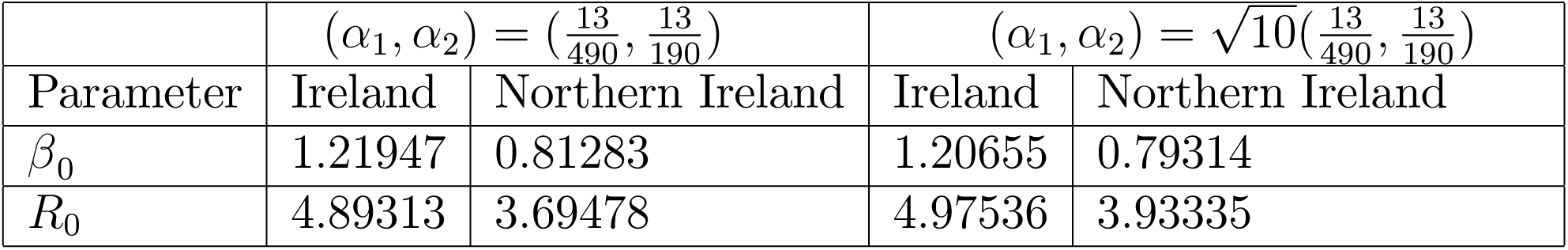

The infective population and the effective reproduction ratio versus time are shown for the low and high (*α*_1_, *α*_2_) cases for Ireland in Figure 11A. The corresponding results for Northern Ireland are shown in Figure 11B. The first peak infective fractions for the two sets of (*α*_1_, *α*_2_) values for the two-region models are as follows:

**FIGURE 11A.**
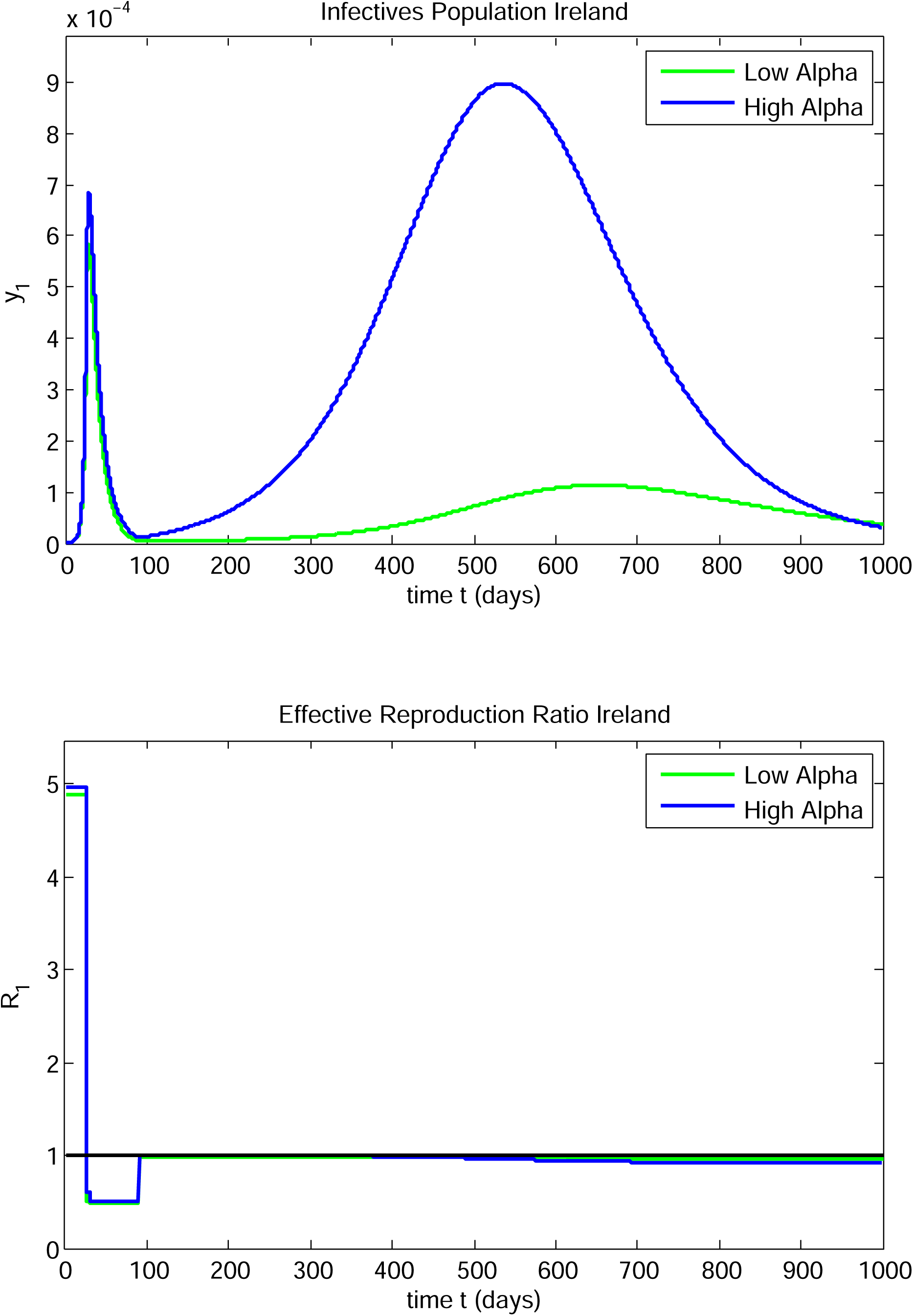

**FIGURE 11B.**
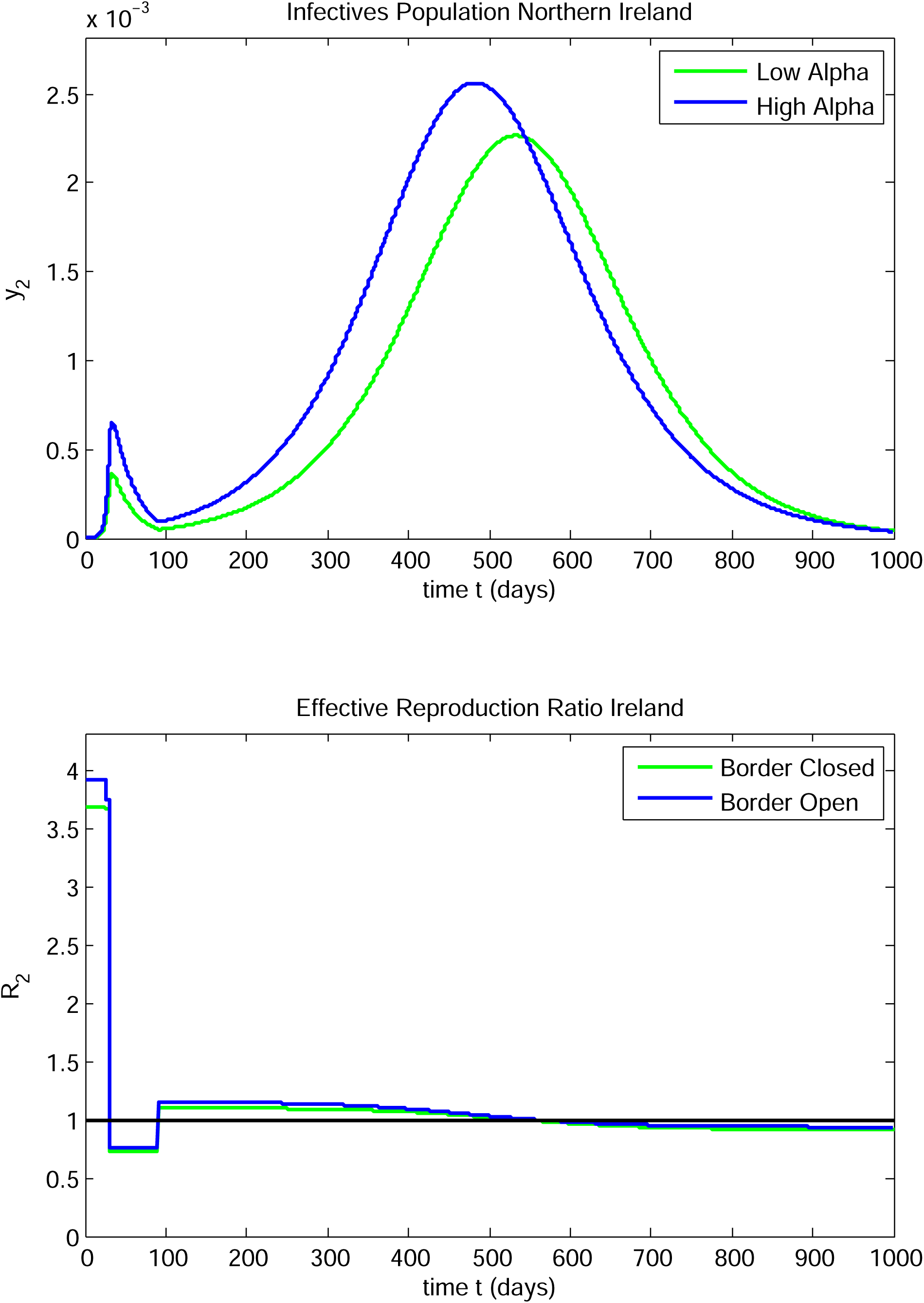

|  | First Peak Infective Fraction |  |  |
| --- | --- | --- | --- |
| Region | $(\alpha_1, \alpha_2) = (\frac{13}{490}, \frac{13}{190})$ | $(\alpha_1, \alpha_2) = \sqrt{10}(\frac{13}{490}, \frac{13}{190})$ | % Diff |
| Ireland | $5.85 \times 10^{-4}$ | $6.83 \times 10^{-4}$ | 18% |
| Northern Ireland | $3.61 \times 10^{-4}$ | $6.52 \times 10^{-4}$ | 78% |

and the corresponding peak times are as follows:

|  | First Peak Time |  |  |
| --- | --- | --- | --- |
| Region | $(\alpha_1, \alpha_2) = (\frac{13}{490}, \frac{13}{190})$ | $(\alpha_1, \alpha_2) = \sqrt{10}(\frac{13}{490}, \frac{13}{190})$ | Diff |
| Ireland | 28.9 | 29.1 | 0.2 |
| Northern Ireland | 33.8 | 34.0 | 0.2 |

The local minimum infective fractions for the two sets of (*α*_1_, *α*_2_) values for the two-region models are as follows:

|  | Local Minimum Infective Fraction |  |  |
| --- | --- | --- | --- |
| Region | $(\alpha_1, \alpha_2) = (\frac{13}{490}, \frac{13}{190})$ | $(\alpha_1, \alpha_2) = \sqrt{10}(\frac{13}{490}, \frac{13}{190})$ | % Diff |
| Ireland | $4.95 \times 10^{-6}$ | $1.16 \times 10^{-5}$ | 134% |
| Northern Ireland | $5.08 \times 10^{-5}$ | $9.43 \times 10^{-5}$ | 86% |

and the corresponding peak times are as follows:

|  | Local Minimum Peak Time |  |  |
| --- | --- | --- | --- |
| Region | $(\alpha_1, \alpha_2) = (\frac{13}{490}, \frac{13}{190})$ | $(\alpha_1, \alpha_2) = \sqrt{10}(\frac{13}{490}, \frac{13}{190})$ | Diff |
| Ireland | 101.9 | 92.5 | 9.4 |
| Northern Ireland | 92.8 | 92.9 | 0.1 |

The second peak infective fractions for the two sets of (*α*_1_, *α*_2_) values for the two-region models are as follows:

|  | Second Peak Infective Fraction |  |  |
| --- | --- | --- | --- |
| Region | $(\alpha_1, \alpha_2) = (\frac{13}{490}, \frac{13}{190})$ | $(\alpha_1, \alpha_2) = \sqrt{10}(\frac{13}{490}, \frac{13}{190})$ | % Diff |
| Ireland | $1.13 \times 10^{-4}$ | $8.98 \times 10^{-4}$ | 695% |
| Northern Ireland | $2.27 \times 10^{-3}$ | $2.56 \times 10^{-3}$ | 13% |

and the corresponding peak times are as follows:

|  | Second Peak Time |  |  |
| --- | --- | --- | --- |
| Region | $(\alpha_1, \alpha_2) = (\frac{13}{490}, \frac{13}{190})$ | $(\alpha_1, \alpha_2) = \sqrt{10}(\frac{13}{490}, \frac{13}{190})$ | Diff |
| Ireland | 653.3 | 537.3 | 116.3 |
| Northern Ireland | 533.8 | 483.2 | 50.6 |

Comparing with Case Study 8, substantial negative effects are experienced in both Ireland and Northern Ireland in this case. The first infectives peaks for both regions occur at about the same time as for the low *α*_*i*_, *i* = 1, 2 case, but with an increased peak level (up 18%) in Ireland and a substantially higher peak level (up 78%) in Northern Ireland. The long low trough and the subsequent long low infectives peak in Ireland are replaced by a short trough followed by a dramatically higher second peak (up 695%) in Ireland. The second Ireland infectives peak is also of considerably longer duration than the first Ireland infectives peak. There is an increase of 13% in the second peak in Northern Ireland and it comes over 50 days earlier, which is also quite significant.

Case Studies 8 and 9, taken together, imply there is a need to obtain more data and seek a more precise values for the border interaction parameters. It should be noted that the model can describe interactions other than those in the immediate neighbourhood of the border. It simply requires interaction between two cohorts of people on either side of the border. So, family, business or holiday travel between the two regions could also be modelled by adjusting the values of *α*_*i*_, *i* = 1, 2.

#### CASE STUDY 10

##### Case A - Sensitive dependence on the size of a transmission/contact reduction intervention

The parameter values for this case are as follows:

| Parameter | Ireland | Northern Ireland |
| --- | --- | --- |
| $\mu$ | 0.319512 | 0.227222 |
| $T_0$ | 2.16939 | 3.050534 |
| $1/\sigma$ | 3.58 | 3.58 |
| $1/a$ | 4 | 4 |
| $t_d$ | 23 | 23 |

Note that the durations of infectiousness are taken to be equal in both regions in this study. There is a single transmission/contact reduction, on the same day in region, with no release intervention. The strength of the transmission/contact reduction was taken to be the same in both regions:

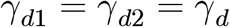

and was varied to study the sensitivity of the evolution with respect to *γ*_*d*_. It was found that the nature, especially location and width, of the infectives peak in Ireland was highly sensitive to the value of *γ*_*d*_ for *γ*_*d*_ near 0.8. The values and times of occurrence of the peak infective fraction in Ireland for seven values of *γ*_*d*_ in the range [0.77, 0.8] are as follows:

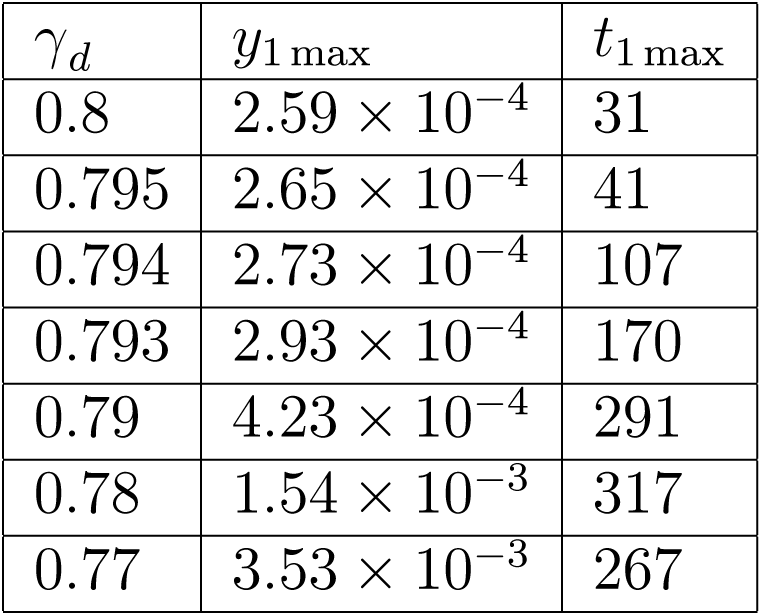

| $\gamma_d$ | $y_{1\max}$ | $t_{1\max}$ |
| --- | --- | --- |
| 0.8 | $2.59 \times 10^{-4}$ | 31 |
| 0.795 | $2.65 \times 10^{-4}$ | 41 |
| 0.794 | $2.73 \times 10^{-4}$ | 107 |
| 0.793 | $2.93 \times 10^{-4}$ | 170 |
| 0.79 | $4.23 \times 10^{-4}$ | 291 |
| 0.78 | $1.54 \times 10^{-3}$ | 317 |
| 0.77 | $3.53 \times 10^{-3}$ | 267 |

where *y*_1 max_ is the peak infectives level in Ireland and *t*_1 max_ is the time (in days) at which it occurs.

Graphs of the Ireland infectives fraction and the effective reproductive ratio versus time for the highest five out of the seven chosen values of *γ*_*d*_ are shown in Figure 12A. Graphs of the Ireland infectives fraction and the effective reproductive ratio versus time for the lowest four values of *γ*_*d*_ are shown in Figure 12B. The width of the peak broadens dramatically under a small decrease in *γ*_*d*_ (e.g., from 0.8 down to 0.78). The height of the Ireland infectives peak increases rapidly also as *γ* _*d*_ is decreased from 0.8 to 0.77 - see Figures 12A and 12B. Note that *R*_1_*(t)* remains close to unity after the intervention on day 23, in each of these cases. This is similar to, but more dramatic than, the sensitivity shown in Case Study 3.

**FIGURE 12A.**
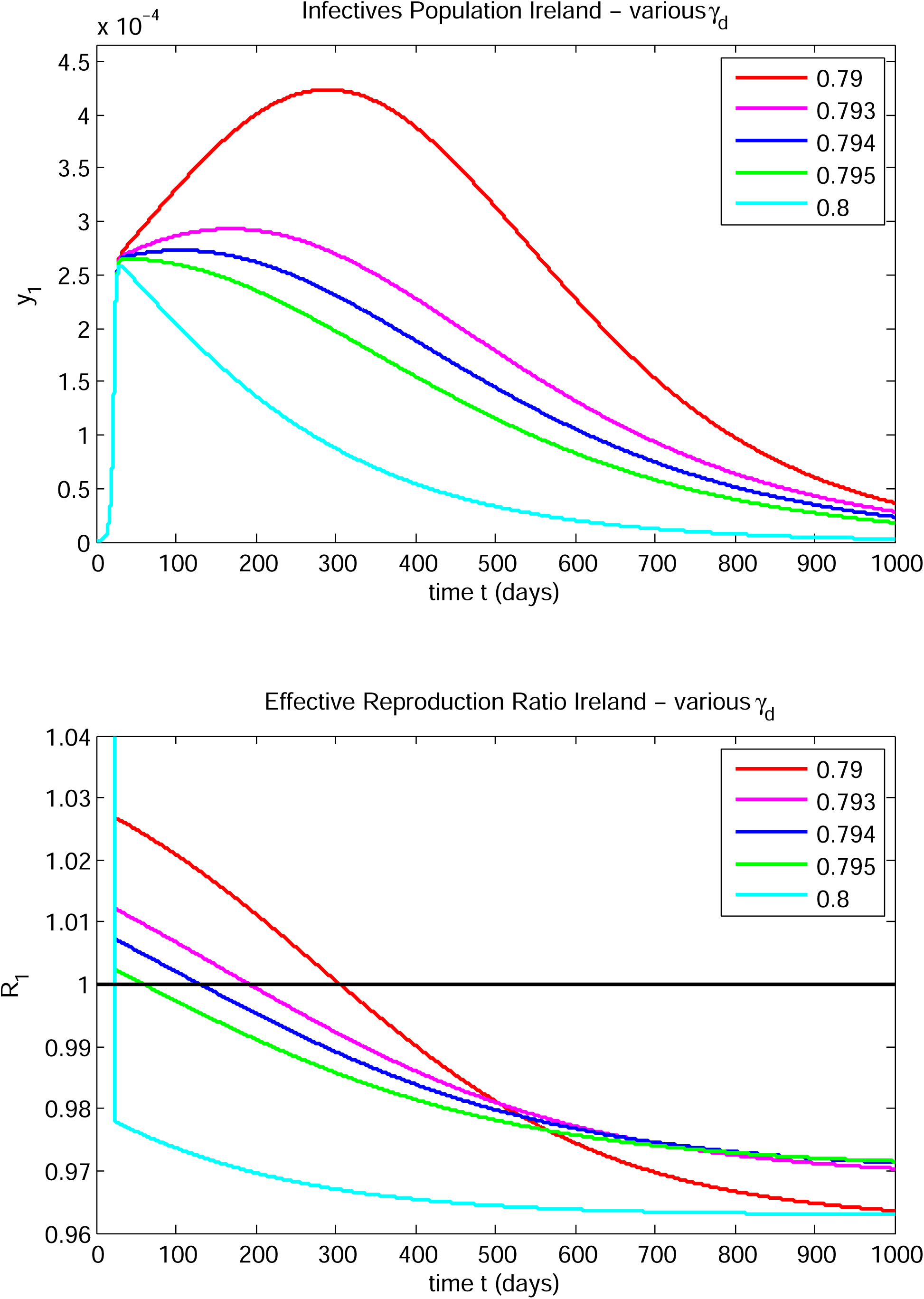

**FIGURE 12B.**
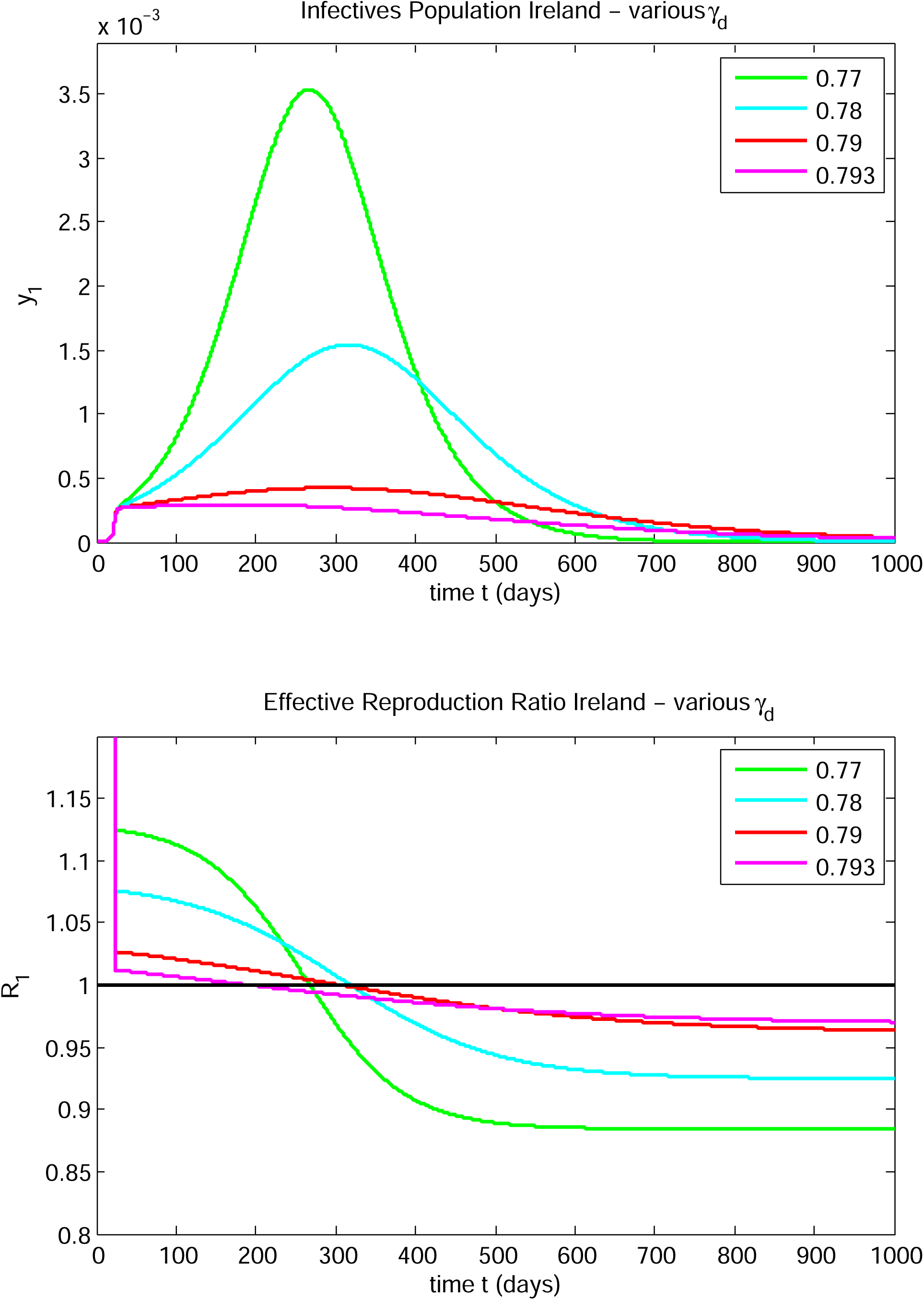

**FIGURE 12C.**
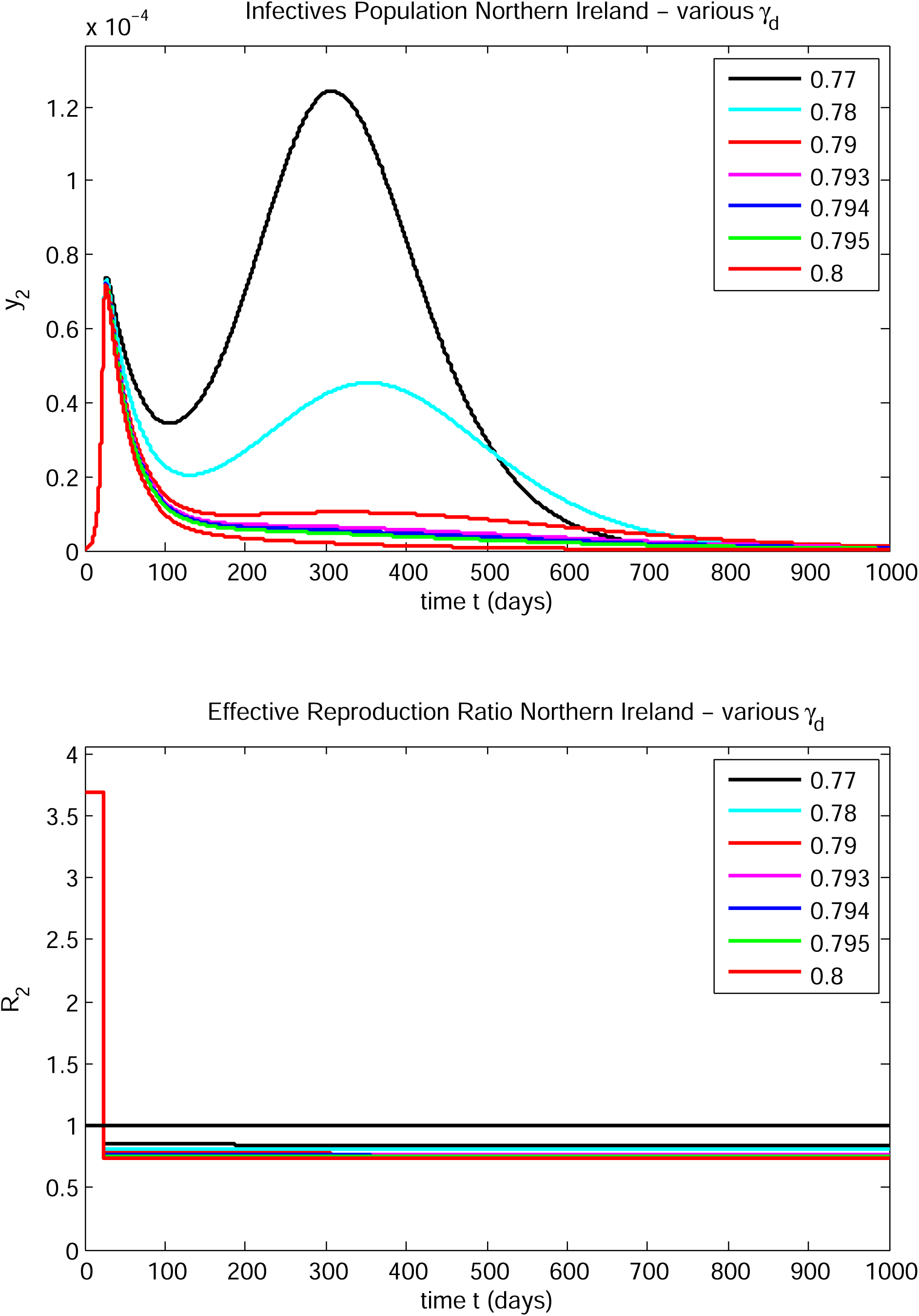

**FIGURE 12D.**
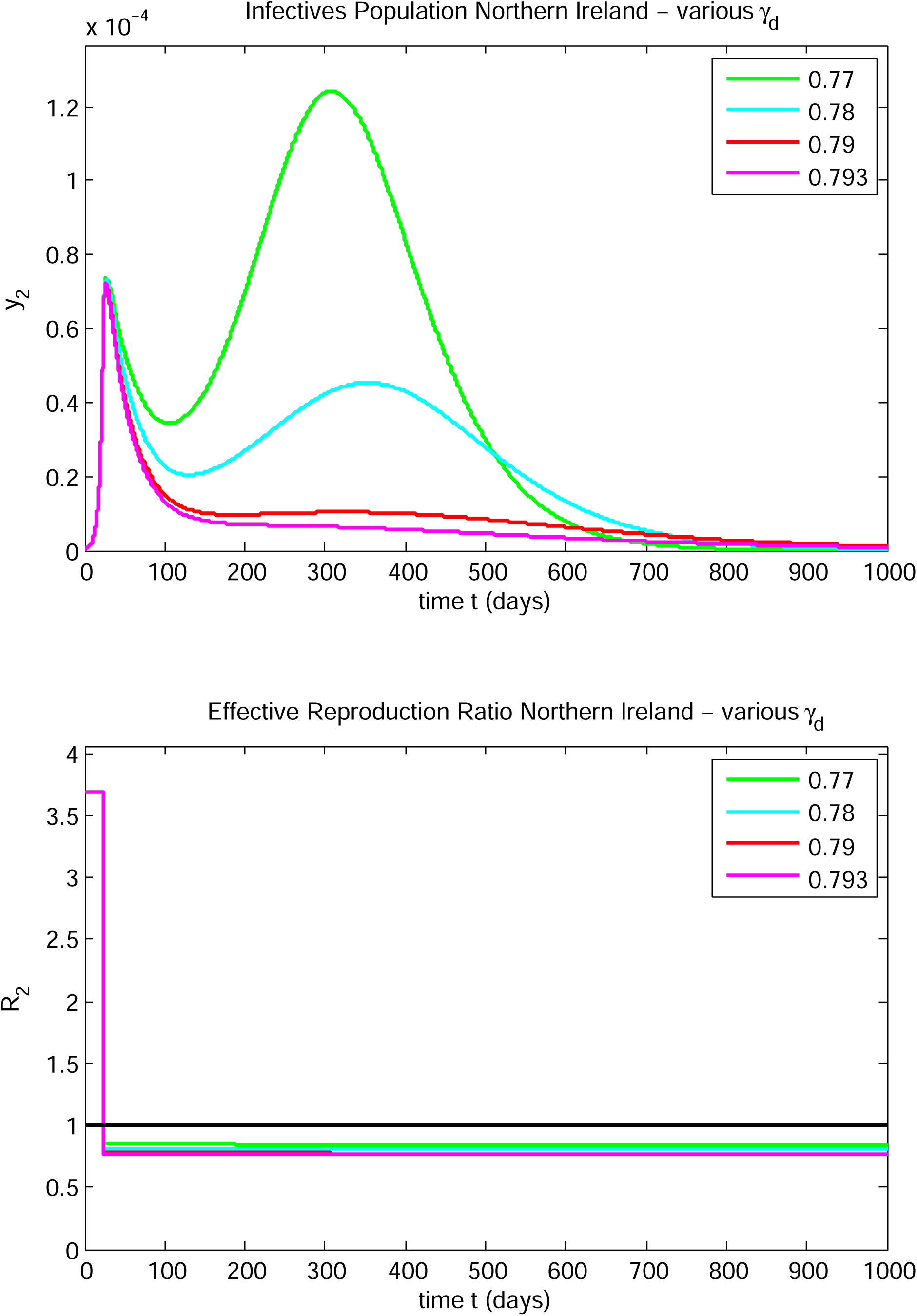

**FIGURE 12E.**
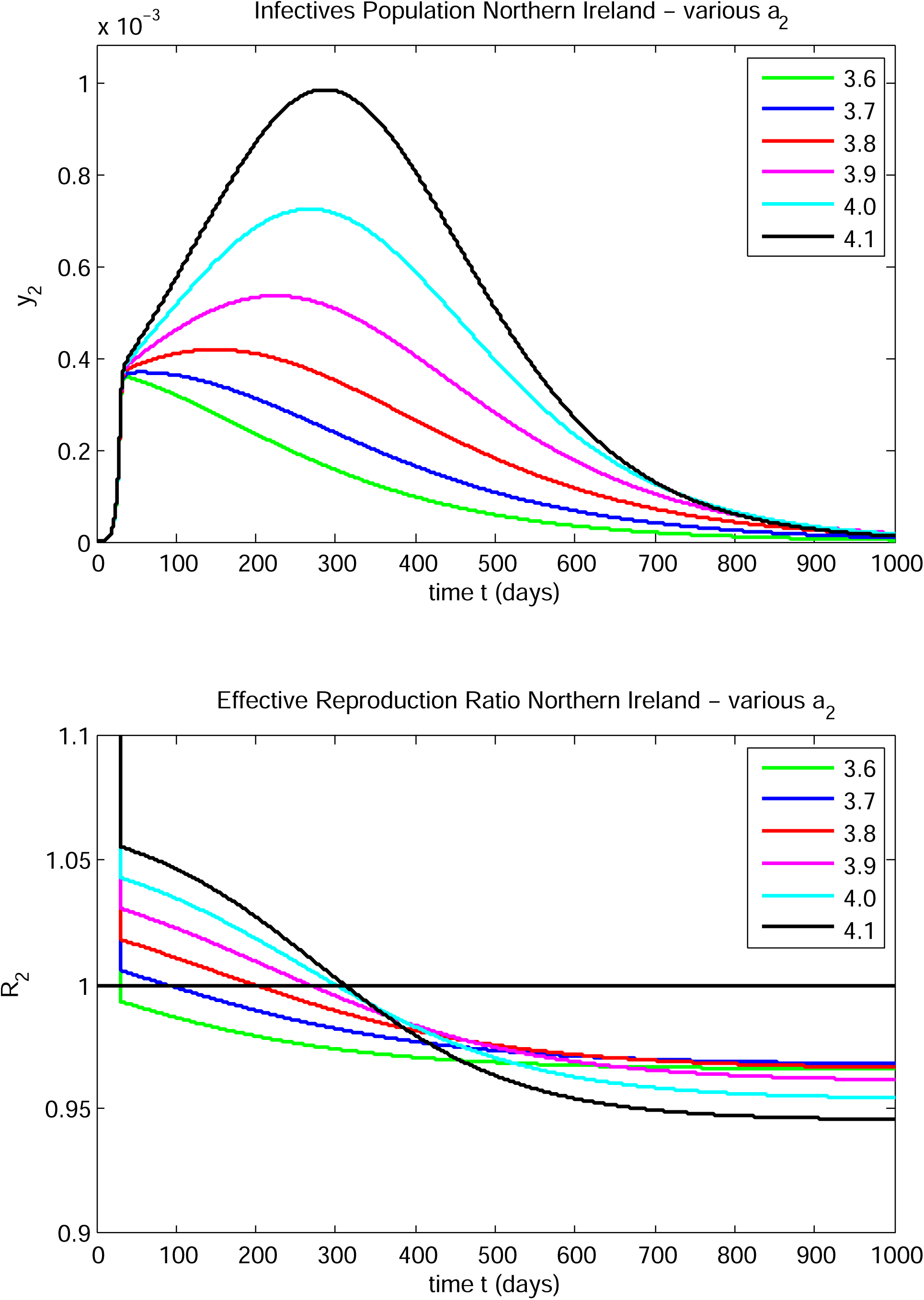

**FIGURE 12F.**
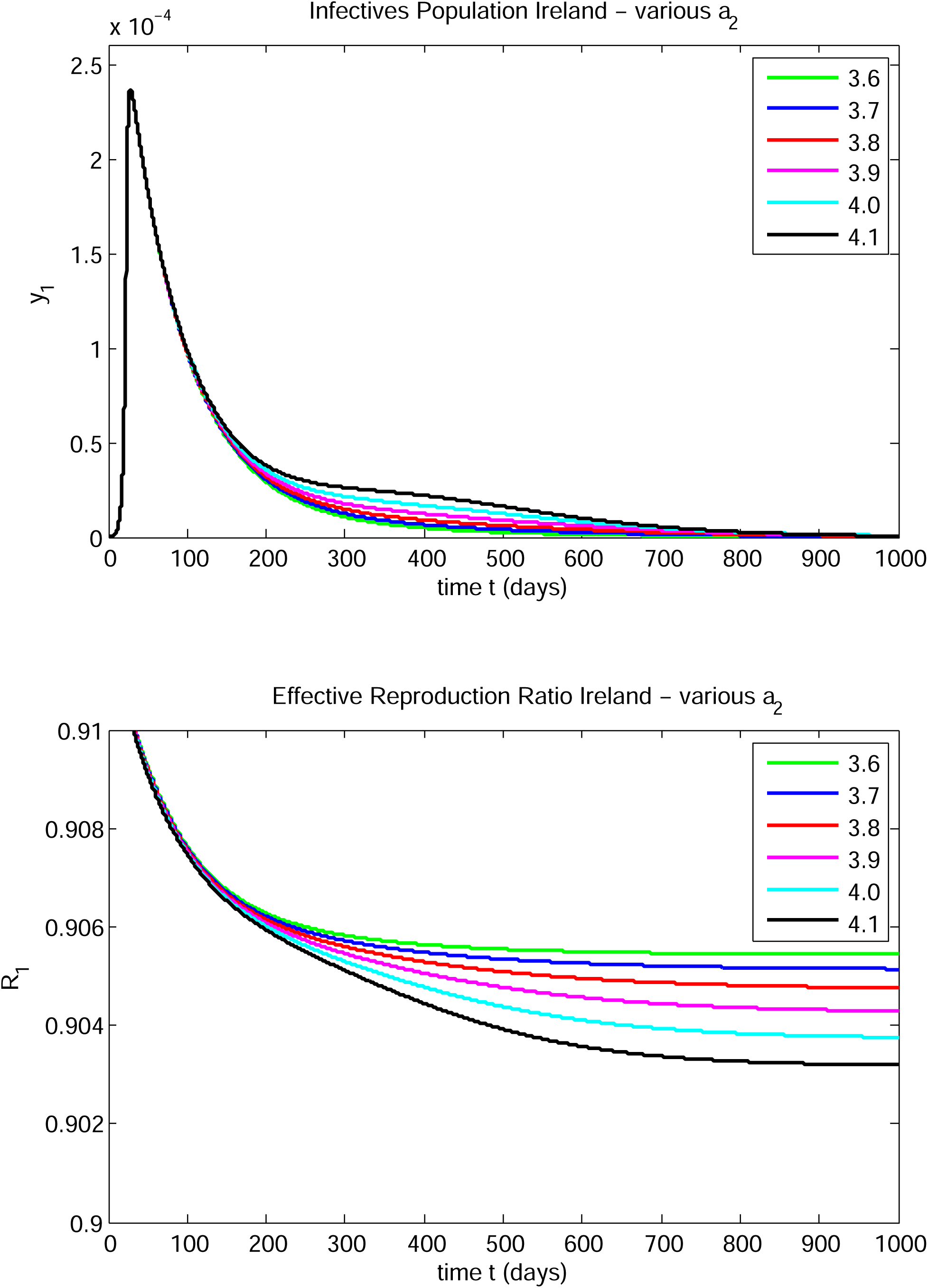

Graphs of the Northern Ireland infectives fraction and effective reproduction ratios are shown in Figures 12C and 12D. As *γ* _*d*_ decreases from 0.8 down to 0.793 there is a single infectives peak in Northern Ireland (see Figure 12C). However, the post-peak tail decays more slowly as *γ* _*d*_ decreases and at *γ* _*d*_ = 0.79 there is clearly a second peak. As *γ* _*d*_ decreases further, the second peak increases in size and becomes much higher than the first peak when 1_*d*_ has decreased to 0.77. As *γ* _*d*_ decreases from 0.79 to 0.77, the second peak in Northern Ireland increases by a factor of over 10. So, there is strong sensitivity in the behaviour of the epidemic in both Ireland and Northern Ireland to a small change in *γ* _*d*_ in this case study.

Note that the effective reproductive ratio *R*_1_ remains in the range (0.85, 1.15) after the intervention on day 23 for all vallues of *γ* _*d*_ considered. It decreases monotonically in all cases from a value slightly above 1 to a value slightly below. After day 23, *R*_2_ decreases monotonically and remains in the range (0.74, 0.85) in all cases considered. So, similar to what was found for Ireland in Case Study 3, a peak emerges in Northern Ireland even though *R*_2_ remains below 1 after day 23. Viewed from the perspective of a single region model for Northern Ireland alone, this would be unexpected and unexplainable.

Note that a reduction in *γ* _*d*_ corresponds to a transmission/contact relaxation. The possibility of a large increase following a very small increase in transmission/contact behaviour for certain model parameter value sets implies a small policy change could, under certain circumstances, cause a large negative development in the epidemic, both north and south of the border.

It would be worth exploring parameter space more thoroughly to see if similar effects could occur for other parameter combinations.

##### Case B - Sensitive dependence on specific infective removal rates

In this case, we find that a slight change in one of the specific infective removal rates can cause a substantial change in the evolution of the epidemic in one of the regions. The parameter values for this case are as follows:

| Parameter | Ireland | Northern Ireland |
| --- | --- | --- |
| $\mu$ | 0.319512 | 0.227222 |
| $T_0$ | 2.16939 | 3.050534 |
| $1/\sigma$ | 3.58 | 3.58 |
| $1/a$ | 3.5 | 3.6 – 4.1 |
| $\gamma_d$ | 0.8 | 0.7 |
| $t_d$ | 23 | 30 |

In this case 1/*a*_1_ has the value 3.5 and 1/*a*_2_ is varied from 3.6 to 4.1. In the table below, (*y*_*i* max_, *t*_*i* max_) denotes the peak infective population and time of occurrence of the peak in region i, i = 1, 2.

| $1/a_2$ | $y_{1\max}$ | $t_{1\max}$ | $y_{2\max}$ | $t_{2\max}$ |
| --- | --- | --- | --- | --- |
| 3.6 | $2.37 \times 10^{-4}$ | 28.3 | $3.60 \times 10^{-4}$ | 38.7 |
| 3.7 | $2.37 \times 10^{-4}$ | 28.3 | $3.70 \times 10^{-4}$ | 57.3 |
| 3.8 | $2.37 \times 10^{-4}$ | 28.3 | $4.19 \times 10^{-4}$ | 150 |
| 3.9 | $2.37 \times 10^{-4}$ | 28.3 | $5.36 \times 10^{-4}$ | 225 |
| 4.0 | $2.37 \times 10^{-4}$ | 28.3 | $7.26 \times 10^{-4}$ | 268 |
| 4.1 | $2.37 \times 10^{-4}$ | 28.3 | $9.86 \times 10^{-4}$ | 288 |

Graphs of the Northern Ireland infective fraction and the effective reproductive ratio versus time for the six values of 1/*a*_2_ are shown in Figure 12E. After the transmission/contact reduction intervention, *R*_2_ decreases from a little above unity to a little below unity in each case. *R*_l_ decreases from about 0.92 to 0.90 over the time period graphed. Again a small change in the parameter 1/*a*_2_ can cause a large change in the height and location of the peak in Northern Ireland; e.g., a change in *a*_2_ from 3.7 to 3.8 causes a 13% increase in the peak level together with a 162% increase (from 57.3 to 150) in the time of occurrence of the infective peak in Northern Ireland. This type of sensitive dependence does not induce a second peak in Ireland, for *a*_2_ in the range (3.6, 4.1) as indicated in Figure 12F, nor does it alter the Ireland peak height or location.

Note that *R*_2_ lies in the range (0.92, 1.06) for values of *a*_2_ in the range (3.6, 4.1). So, a large peak can occur in Northern Ireland even though *R*_2_ remains very close to 1. Note that *R*_1_ < 1 and remains close to 0.9 after day 30 in all cases considered. So, in this case the high peaks in Northern Ireland are not driven by infectives in Ireland, but, rather, in Northern Ireland (cf. Case Studies 6 and 7).

#### CASE STUDY 11

In this study, we set as many parameters in both regions equal to one another, in an attempt to model an all-island coordination of policy and societal response, starting at the beginning of the epidemic, and including a transmission/contact reduction, but not including a transmission/contact relaxation. We chose parameters as follows:

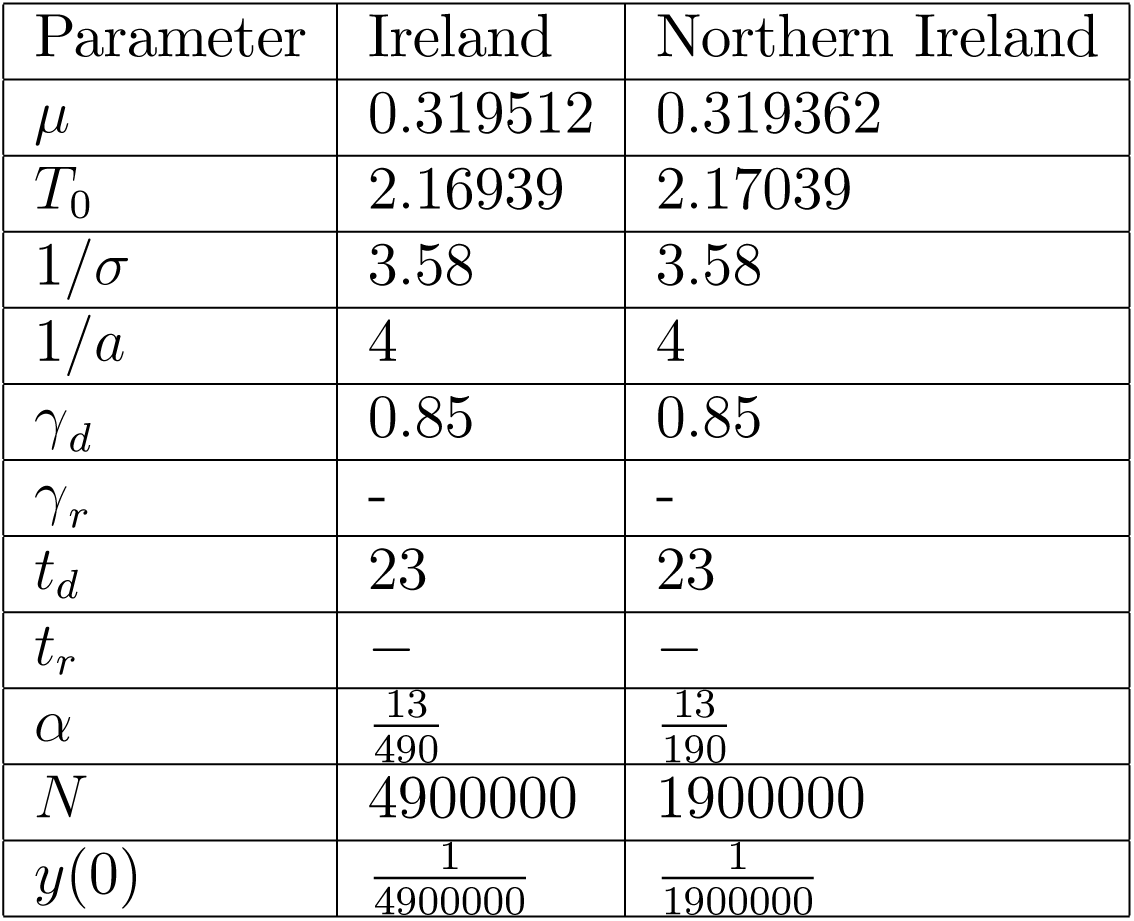

| Parameter | Ireland | Northern Ireland |
| --- | --- | --- |
| $\mu$ | 0.319512 | 0.319362 |
| $T_0$ | 2.16939 | 2.17039 |
| $1/\sigma$ | 3.58 | 3.58 |
| $1/a$ | 4 | 4 |
| $\gamma_d$ | 0.85 | 0.85 |
| $\gamma_r$ | - | - |
| $t_d$ | 23 | 23 |
| $t_r$ | — | — |
| $\alpha$ | $\frac{13}{490}$ | $\frac{13}{190}$ |
| $N$ | 4900000 | 1900000 |
| $y(0)$ | $\frac{1}{4900000}$ | $\frac{1}{1900000}$ |

We also set the initial doubling times almost equal. The model could be re-run with the other values indicated by the data for Northern Ireland. The only parameters that are different are the populations (north and south), the border interaction coefficients *α*_*i*_, *i* = 1, 2, and the initial infectives fractions (which depend on the populations). The derived parameters were as follows:

|  | Border Closed |  | Border Open |  |
| --- | --- | --- | --- | --- |
| Parameter | Ireland | Northern Ireland | Ireland | Northern Ireland |
| $\beta_0$ | 1.22095 | 1.22033 | 1.21874 | 1.21812 |
| $R_0$ | 4.88380 | 4.88134 | 4.89607 | 4.91300 |

The sizes and locations of the peak infectives fractions are as follows:

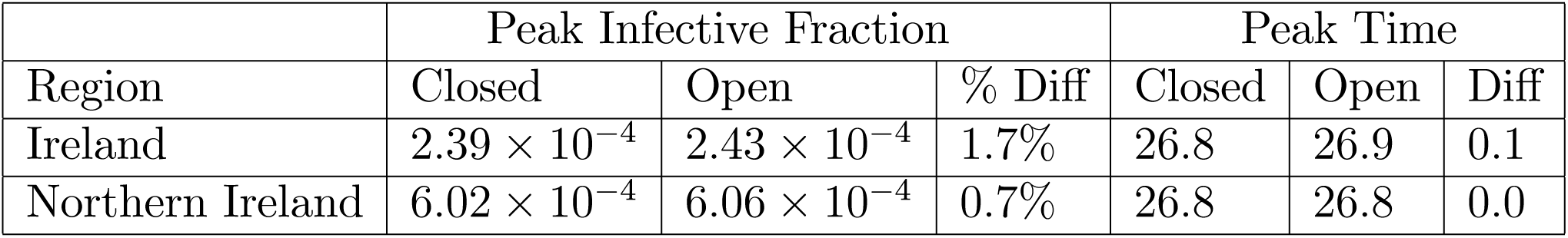

Plots of the infective fraction and effective reproduction ratio for each region in both the open and closed border cases are shown in Figures 13A and 13B. Plots of the Ireland and Northern Ireland infective fractions and of the corresponding effective reproductive ratios versus time are shown together in Figure 13C. In this case, the border status has little effect on the evolution of the epidemic in each region. However, from the tables and Figure 13C, we see that the infective fraction peak in Northern Ireland is substantially higher than that in Ireland. Is there an inherent difference between the two regions causing this? The main differences are the population sizes, the border interaction parameter values and the initial values of the infective fractions. The values of all these parameters are outside the control of the governments, north and south. We re-ran the model with one change: make *γ* _2_ = *γ* _1_. This amounts to reducing the fraction of the Northern Ireland population interacting with the Ireland population. We found that this did not reduce the difference in peak sizes - rather, it increased the ratio of the Northern Ireland infectives peak to the Ireland infectives peak slightly.

**FIGURE 13A.**
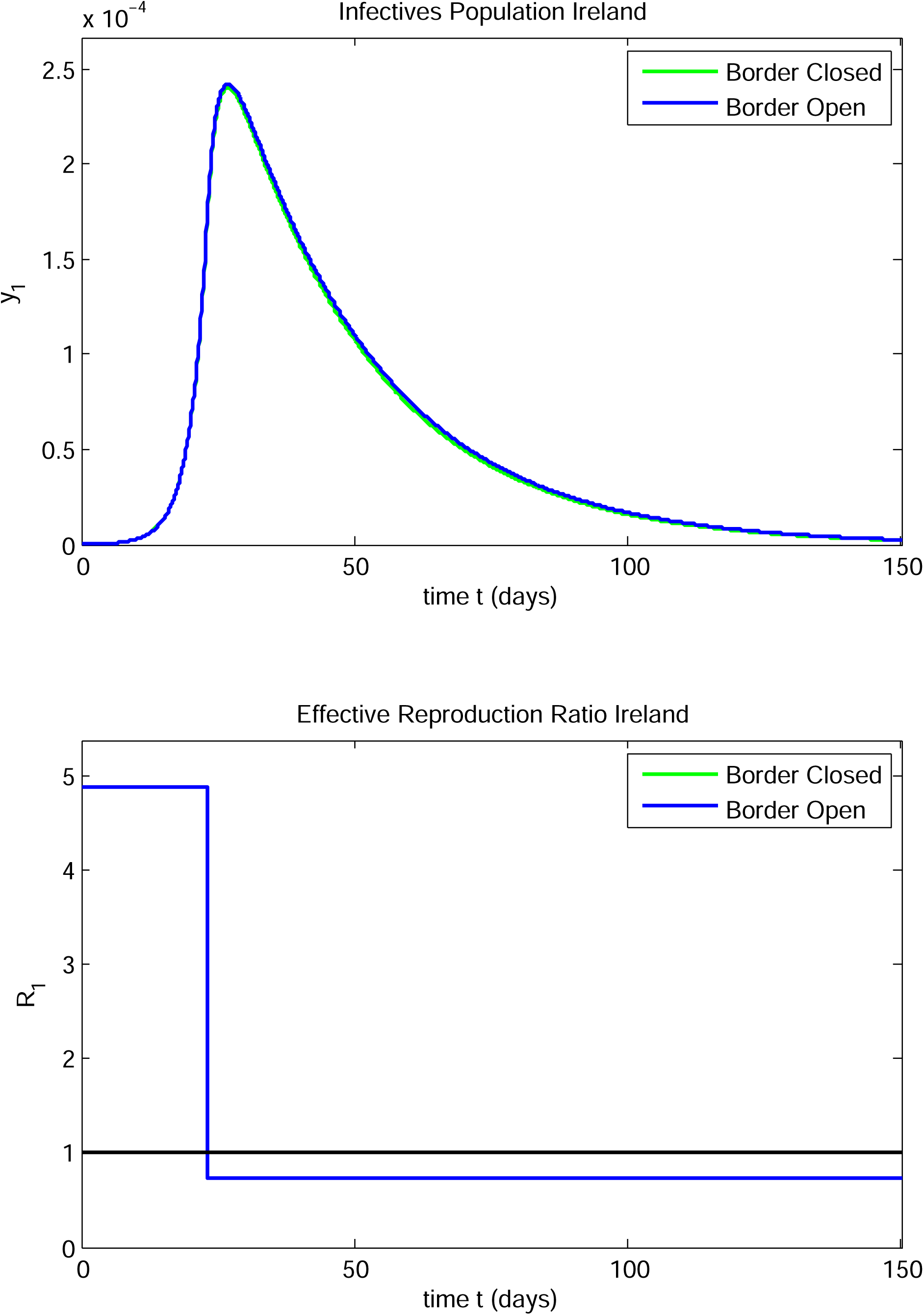

**FIGURE 13B.**
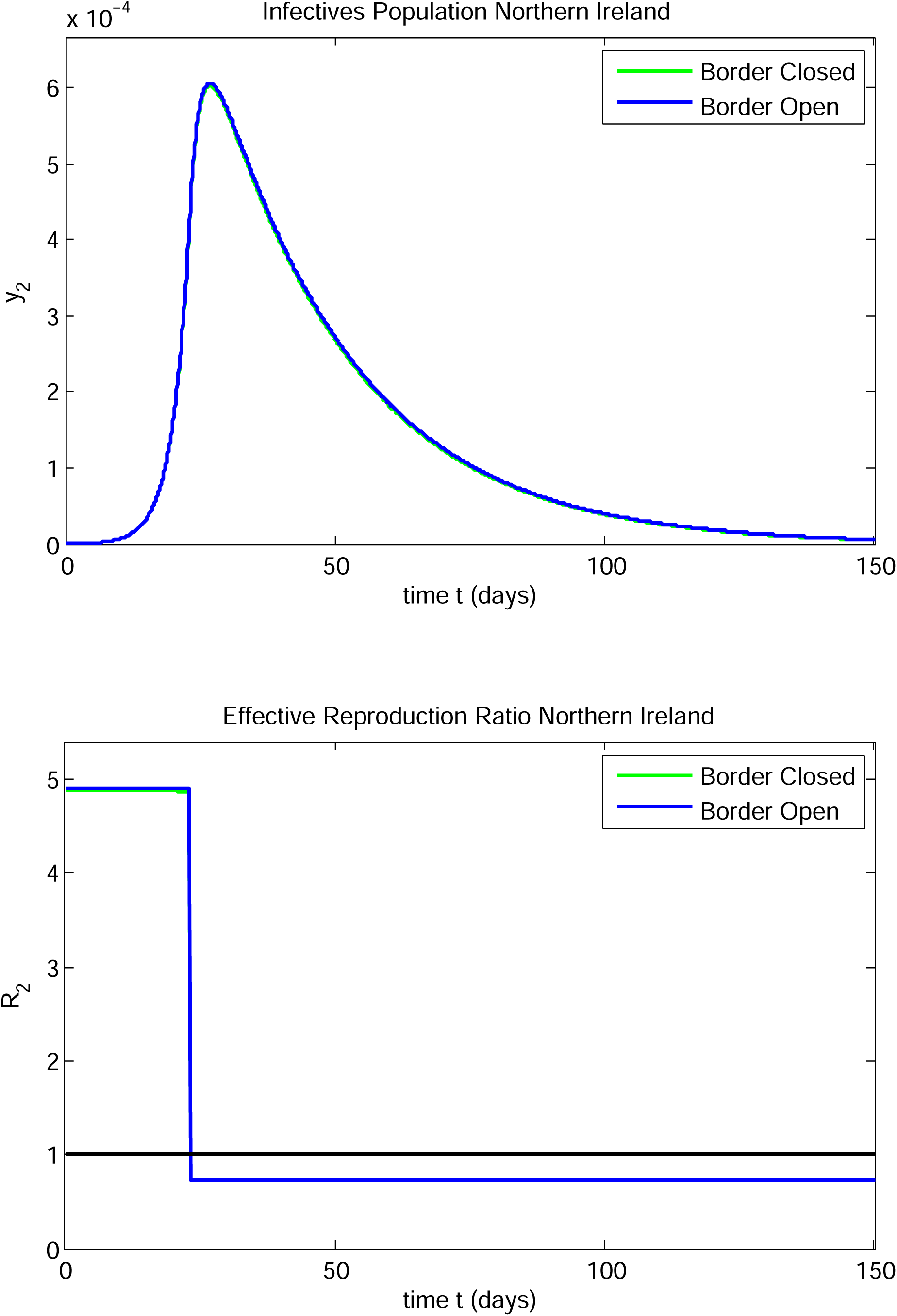

**FIGURE 13C.**
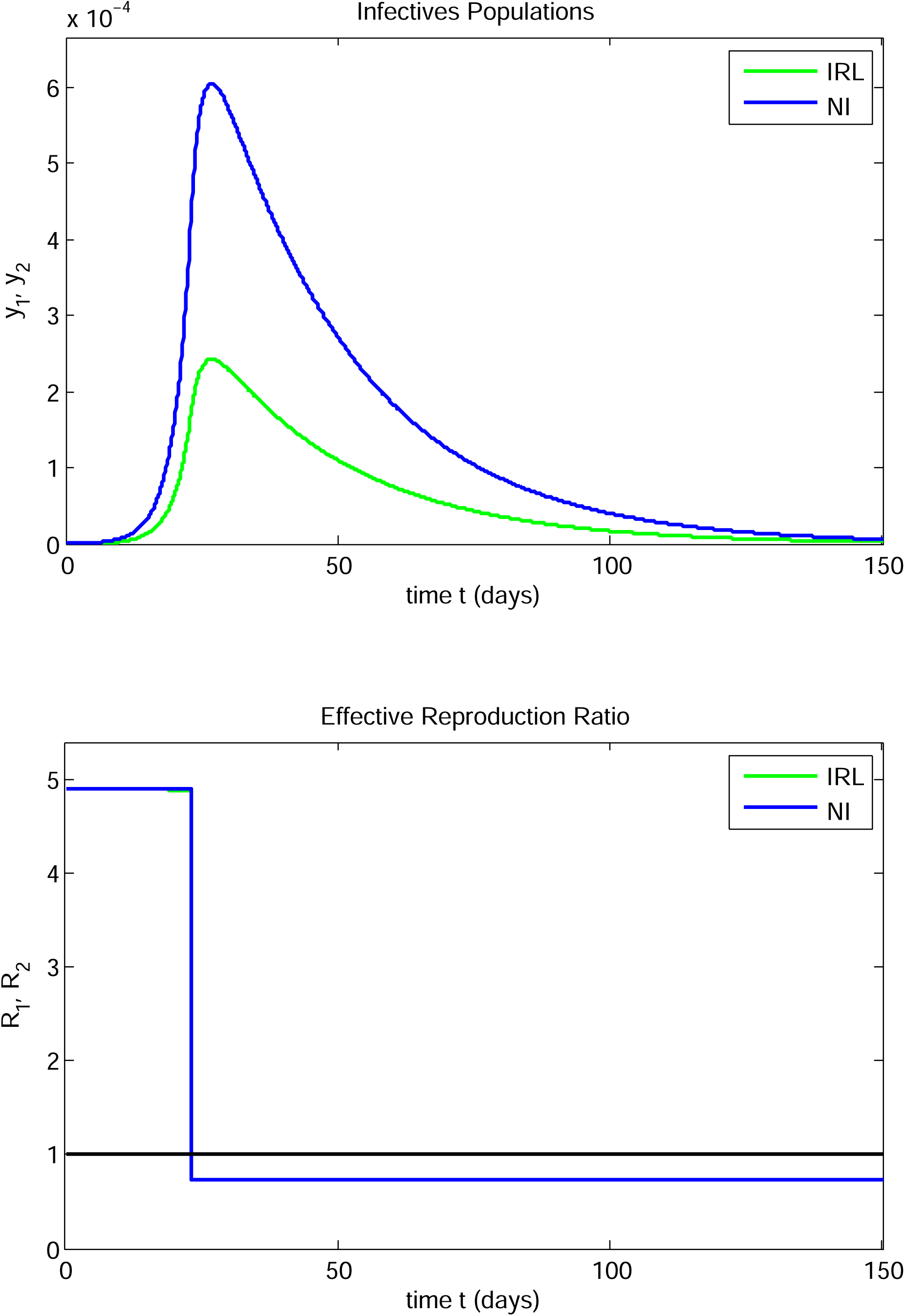

The populations of the two regions cannot be altered. This leaves the initial conditions as the source of the difference between the two countries. We investigate this in Case Study 12.

#### CASE STUDY 12

In this Case Study we investigate the dependence of the evolution in the two regions on the initial conditions by fixing the initial Northern Ireland number of infectives to be 1 and varying the initial number of infectives in Ireland.

Except for the initial conditions, the parameters the parameters were:

| Parameter | Ireland | Northern Ireland |
| --- | --- | --- |
| $\mu$ | 0.319512 | 0.319362 |
| $T_0$ | 2.16939 | 2.17039 |
| $1/\sigma$ | 3.58 | 3.58 |
| $1/a$ | 4 | 4 |
| $\gamma_d$ | 0.85 | 0.85 |
| $\gamma_r$ | - | - |
| $t_d$ | 23 | 23 |
| $t_r$ | — | — |
| $\alpha$ | $\frac{13}{490}$ | $\frac{13}{190}$ |
| $N$ | 4900000 | 1900000 |

The derived parameters are the same as in Case Study 11 and are as follows:

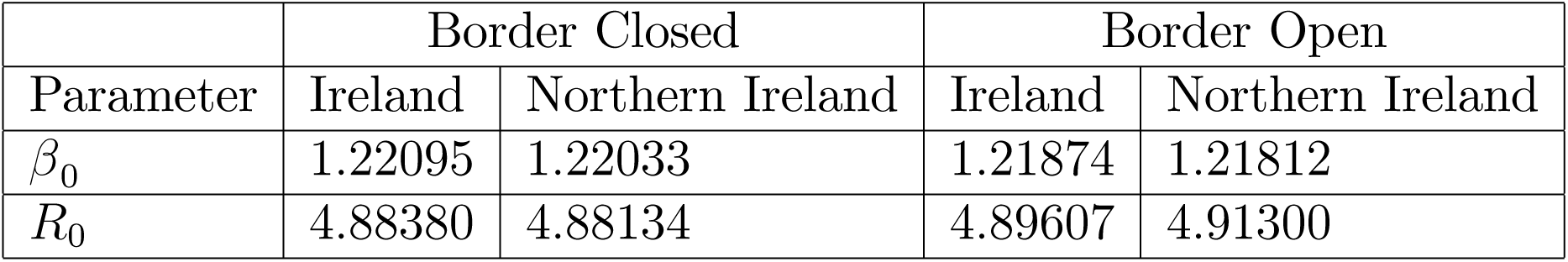

The initial number of infectives in region *i* is denoted by *I*_*i*0_, *i* = 1, 2. The initial fraction of infectives in region *i* is denoted by *y*_*i*0_, i = 1, 2 and

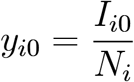

In the cases considered there is a single peak in each region. The peak infectives fraction in region i is denoted by *y*_*i max*_, *i* = 1, 2 and the time at which the peak in region i occurs is denoted by *t*_*i max*_, *i* = 1, 2. We considered several initial conditions to explore how the peaks depend on them:

| $I_{10}$ | $I_{20}$ | $y_{1\max} \times 10^4$ | $t_{1\max}$ | $y_{2\max} \times 10^4$ | $t_{2\max}$ | $y_{2\max}/y_{1\max}$ |
| --- | --- | --- | --- | --- | --- | --- |
| 1 | 1 | 2.426 | 26.9 | 6.057 | 26.8 | 2.497 |
| 2 | 1 | 4.740 | 26.8 | 6.165 | 26.9 | 1.301 |
| 3 | 1 | 7.052 | 26.8 | 6.273 | 26.9 | 0.890 |
| 1 | 2 | 2.534 | 26.9 | 11.984 | 26.8 | 4.729 |
| 1 | 3 | 2.642 | 26.9 | 17.889 | 26.8 | 6.771 |
| 3 | 2 | 7.159 | 26.9 | 12.199 | 26.8 | 1.704 |

The initial infectives populations must be integers. If *I*_20_ is held fixed at 1 and *I*_10_ is increased from 1 to 3, the ratio *y*_2 max_/*y*_1 max_ decreases from roughly 2.5 to 0.9. If *I*_l0_ is held fixed at 1 and *I*_*20*_ is increased from 1 to 3, the ratio *y*_2 max_*/y*_1 max_ increases from approximately 2.5 to 6.8. A plot of the Ireland and Northern Ireland infective fractions and of the corresponding effective reproduction ratios versus time for the open border case in which *I*_10_ = 3, *I*_20_ = 1 are shown in Figure 14. The evolution of the infective fraction in both countries is fairly close in contradistinction to the *I*_l0_ *= I*_20_ = 1 case considered in Case Study 11. The above choices are somewhat artificial in the sense that the SEIR model is not considered to be accurate for very small infective population sizes [14]. Another approach would be to consider evolution starting from some time after the epidemic has started. We investigated this and found a similar effect to that observed here.

**FIGURE 14.**
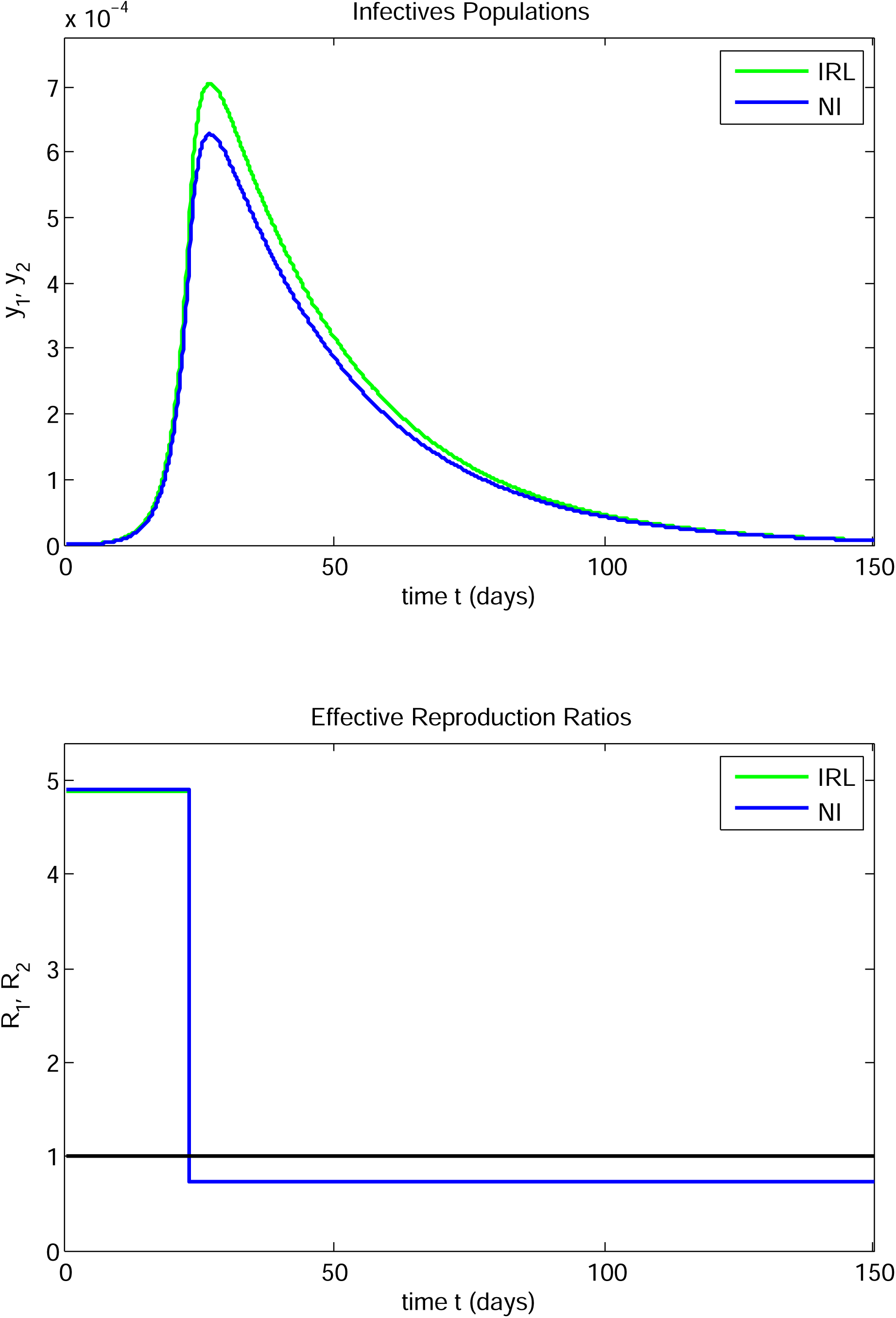

As noted, the initial conditions are not usually under the control of governments, neither are large differences in populations between two regions. Collectively, they determine the initial fractions of infectives. So, this difference between the evolution of the epidemic in the two regions is essentially due to the differences in size of the populations, as well as the initial numbers of infectives. If the epidemic was not to be worse in one region than the other, coordination of policies north and south would need to be considered, recognizing such inherent differences between the regions. This would involve developing policies to vary the parameters that can be affected by policy, such as γ _*di*_, *γ* _*ri*_, *γ* _*di*_, *γ* _*ri*_ and *a*_*i*_, *i* = 1, 2, recognising that some differences are caused by parameters that cannot be controlled by governments. How this might be done needs further investigation.

This case also illustrates a third type of sensitivity in the model - sensitivity to small changes in an initial condition, the others being sensitivity to small changes in *γ* _*di*_ or *a*_*i*_ in certain situations.

We also present another comparison. This involves replacing the Ireland population taken to be 4.9 × 10^6^ in the model, by a more recent estimate of the Ireland population, viz: 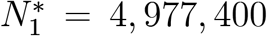, and considering the case in which *I*_l0_ = *I*_20_ = 1. This is equivalent to using the population value *N*_l_ = 4.9 × 10^6^ with 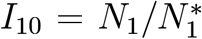 in the above model. The resulting peak values and locations for these two cases are as follows:

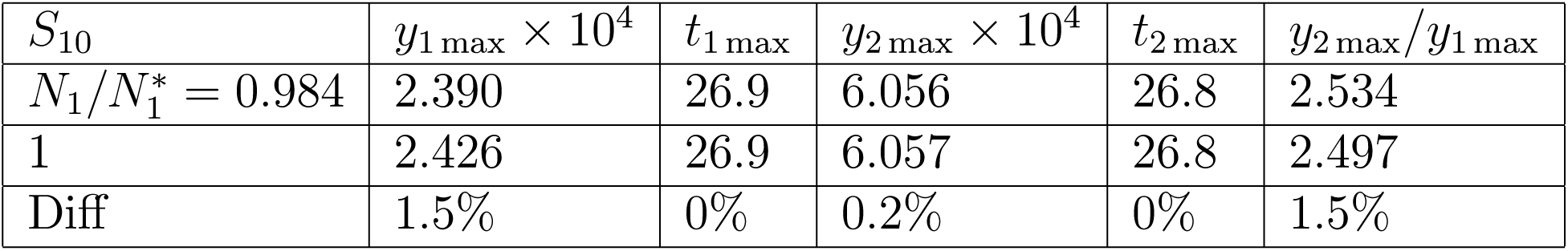

So, use of the value 4.9 × 10^6^ for the population of Ireland, rather than the more recent accurate value, produces a small error in the peak sizes, and is equivalent to using the more accurate population value with a slightly lower initial infective population fraction.

#### CASE STUDY 13

This case is similar to Case Study 5 with the parameters for Ireland and Northern Ireland interchanged. The parameters are as follows:

| Parameter | Ireland | Northern Ireland |
| --- | --- | --- |
| $\mu$ | 0.319512 | 0.319365 |
| $T_0$ | 2.16939 | 2.17039 |
| $1/\sigma$ | 3.58 | 3.58 |
| $1/a$ | 4.5 | 4 |
| $\gamma_d$ | 0.84 | 0.88 |
| $\gamma_r$ | 0.7 | 0.88 |
| $t_d$ | 30 | 23 |
| $t_r$ | 90 | 90 |

There was a single transmission/contact reduction intervention in each region. There was also a transmission/contact increase corresponding to a lockdown release in Ireland on day 90, whereas a low value of *R*_2_ was maintained in Northern Ireland. The derived parameters were as follows:

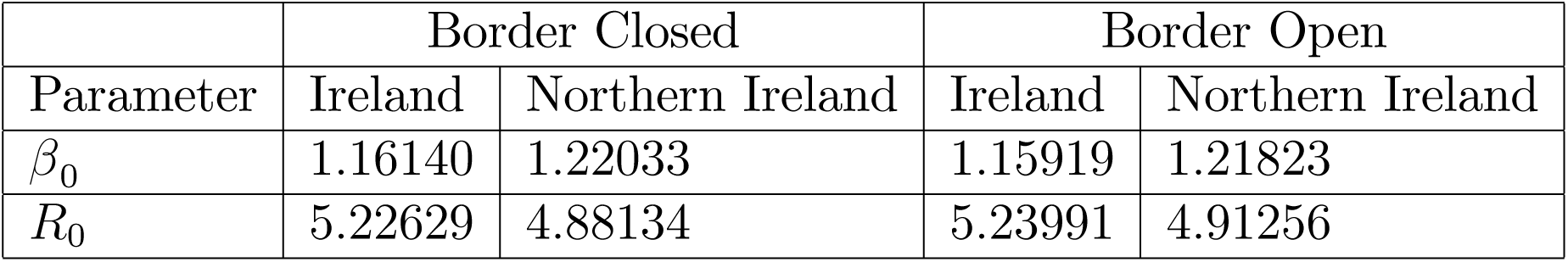

|  | Border Closed |  | Border Open |  |
| --- | --- | --- | --- | --- |
| Parameter | Ireland | Northern Ireland | Ireland | Northern Ireland |
| $\beta_0$ | 1.16140 | 1.22033 | 1.15919 | 1.21823 |
| $R_0$ | 5.22629 | 4.88134 | 5.23991 | 4.91256 |

The infective fraction and the effective reproduction ratio versus time are shown for both the open and closed border cases for Ireland in Figure 15A. The corresponding results for Northern Ireland are shown in Figure 15B. In the closed border case, the infectives fraction rose to a maximum and decayed to zero in Northern Ireland. In the open border case, the infectives fraction in Northern Ireland rose to a peak, decayed to a local minimum and increased again to reach a broader second peak after which the fraction decayed to zero. In both the open and closed border cases, there were two infective peaks in Ireland.

|  | 1st Peak Infectives Fraction |  |  | 1st Peak Time |  |  |
| --- | --- | --- | --- | --- | --- | --- |
| Region | Closed | Open | % Diff | Closed | Open | Diff |
| Ireland | $2.39 \times 10^{-3}$ | $2.42 \times 10^{-3}$ | 1.3% | 34.9 | 34.8 | 0.1 |
| Northern Ireland | $5.84 \times 10^{-4}$ | $5.88 \times 10^{-4}$ | 0.7% | 26.2 | 26.1 | 0.1 |

**FIGURE 15A.**
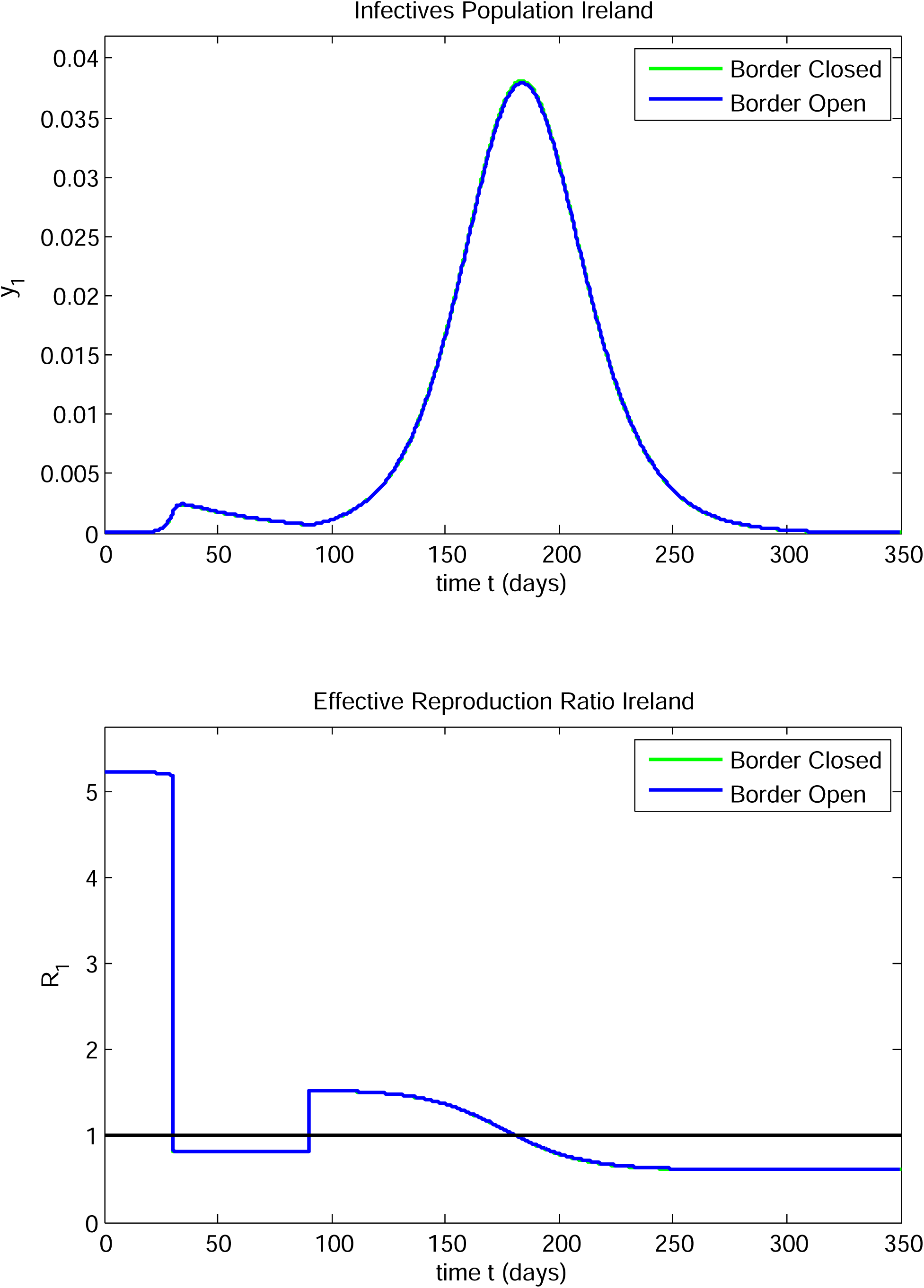

**FIGURE 15B.**
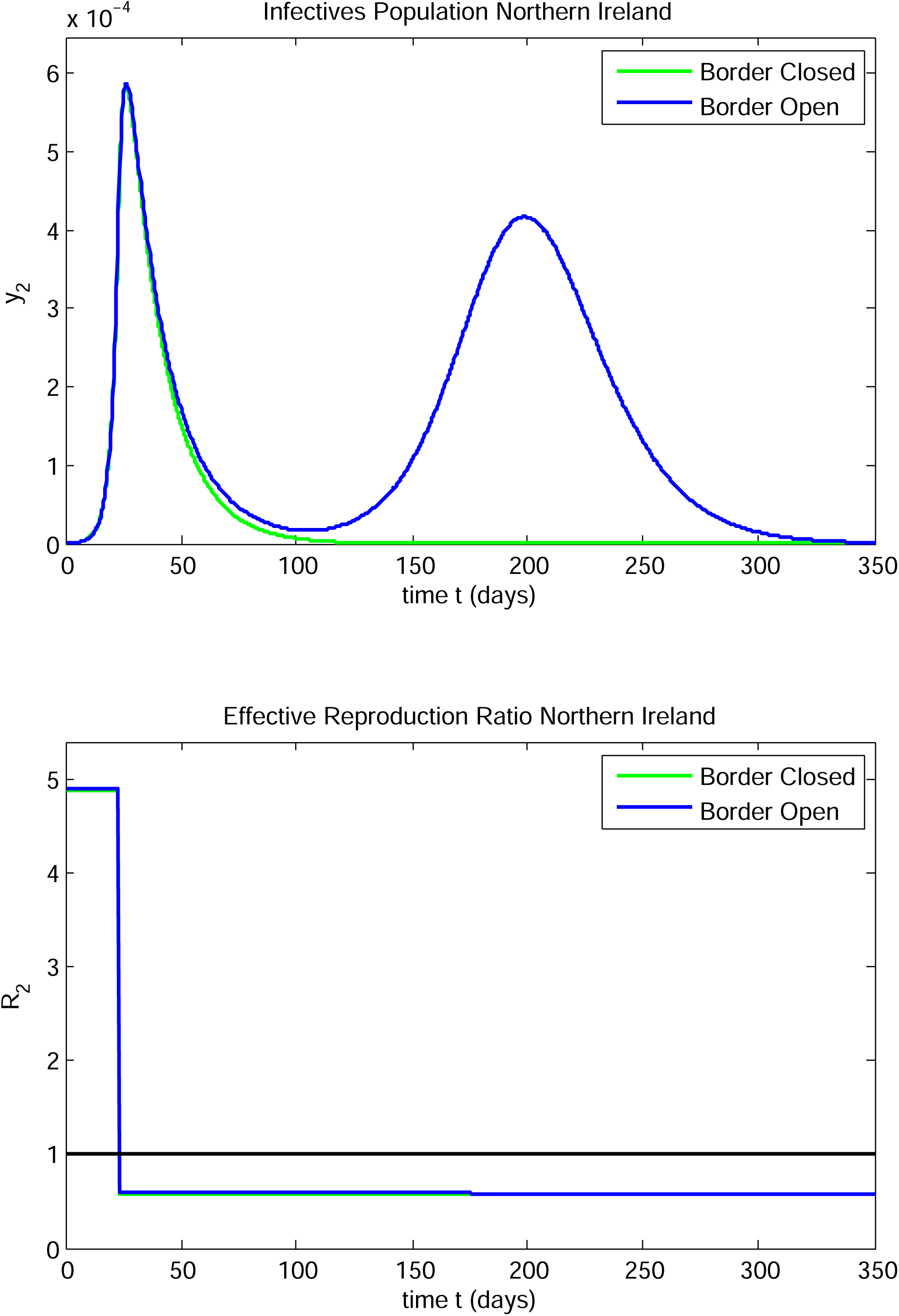

The border had a much greater effect on the evolution in Northern Ireland than in Ireland. Instead of a decay to zero after the infective peak at 26.1 days, the infective fraction appeared to be decaying to zero as in the closed border case, but on day 90 it began to increase again up to a new peak on day 184 after which it decayed to zero. The heights and locations of the local minimum and second peak are shown in the tables:

| Region | Minimum Infectives Fraction |  | Minimum Infectives time |  |
| --- | --- | --- | --- | --- |
|  | Closed | Open | Closed | Open |
| Ireland | $7.02 \times 10^{-4}$ | $7.09 \times 10^{-4}$ | 90.6 | 90.4 |
| Northern Ireland | - | $1.66 \times 10^{-5}$ | - | 104.9 |

| Region | 2nd Peak Infectives Fraction |  | 2nd Peak Time |  |
| --- | --- | --- | --- | --- |
|  | Closed | Open | Closed | Open |
| Ireland | $3.81 \times 10^{-2}$ | $3.79 \times 10^{-2}$ | 183.6 | 183.5 |
| Northern Ireland | - | $4.17 \times 10^{-4}$ | - | 198.8 |

During the period from day 30 to day 90, *R*_1_*(t)* remained slightly over 0.8. On day 90, *R*_1_*(t)* jumped to slightly over 1.5. It decreased thereafter, reaching the value 1 on day 182 and decreasing to slightly over 0.6 by day 350. During the entire period after day 23, the value of *R*_2_*(t)* remained very close to 0.6. If there was no awareness of the interaction taking place between Northern Ireland and Ireland, it could have reasonably been assumed that the epidemic was well-controlled in Northern Ireland and the subsequent rise to a second peak after the infective level had dropped to a low level at day 105 would not have been anticipated. It would not be explainable using the one country model. So,contingencies in the post-peak period to be aware of possible impacts of Ireland government interventions and societal responses on the progress of the epidemic in Northern Ireland.

